# The epidemiology of herpes simplex virus type 2 in sub-Saharan Africa: systematic review, meta-analyses, and meta-regressions

**DOI:** 10.1101/2021.01.25.21250443

**Authors:** Manale Harfouche, Farah M. Abu-Hijleh, Charlotte James, Katharine J. Looker, Laith J. Abu-Raddad

## Abstract

**Background:** Herpes simplex virus type 2 (HSV-2) infection is a prevalent sexually transmitted infection with a sizable disease burden that is highest in sub-Saharan Africa. This study aimed to characterize HSV-2 epidemiology in this region.

**Methods:** Cochrane and PRISMA guidelines were followed to systematically review, synthesize, and report HSV-2 related findings. Meta-analyses and meta-regressions were conducted.

**Findings:** From 218 relevant publications, 451 overall outcome measures and 869 stratified measures were extracted. Pooled incidence rates ranged between 2.4-19.4 per 100 person-years across populations. Pooled seroprevalence was lowest at 37.3% (95% confidence interval (CI): 34.9-39.7%) in general populations and high in female sex workers and HIV positive individuals at 62.5% (95% CI: 54.8-70.0%) and 71.3% (95% CI: 66.5-75.9%), respectively. In general populations, pooled seroprevalence increased steadily with age. Compared to women, men had a lower seroprevalence with an adjusted risk ratio (ARR) of 0.61 (95% CI: 0.56-0.67).

Seroprevalence decreased in recent decades with an ARR of 0.98 (95% CI: 0.97-0.99) per year. Seroprevalence was highest in Eastern and Southern Africa. Pooled HSV-2 proportion in genital ulcer disease was 50.7% (95% CI: 44.7-56.8%) and in genital herpes it was 97.3% (95% CI: 84.4-100%).

**Interpretation:** Seroprevalence is declining by 2% per year, but a third of the population is infected. Age and geography play profound roles in HSV-2 epidemiology. Temporal declines and geographic distribution of HSV-2 seroprevalence mirror that of HIV prevalence, suggesting sexual risk behavior has been declining for three decades. HSV-2 is the etiological cause of half of GUD and nearly all genital herpes cases.

**Funding:** This work was supported by pilot funding from the Biomedical Research Program at Weill Cornell Medicine in Qatar and by the Qatar National Research Fund [NPRP 9-040-3-008].

## Introduction

Herpes simplex virus type 2 (HSV-2) infection is a highly prevalent sexually transmitted infection (STI) worldwide.(1) It is a leading cause of genital ulcer disease (GUD) and genital herpes, manifesting in the form of painful, recurrent, and frequent genital lesions.(2–8) Its vertical transmission from mother-to-child can lead to neonatal herpes, a severe and sometimes fatal outcome in newborns.(3, 9) HSV-2 is linked to a 3-fold increase in sexual transmission and acquisition of HIV,(10–12) implying a potential epidemiological synergy between the two viruses.(11, 13, 14)

HSV-2 is typically asymptomatic in most of those who acquire it.(3) This trait coupled with HSV-2’s chronic and reactivating nature, as well as its subclinical shedding,(15, 16) increases its rate of transmission and leads to high antibody prevalence (seroprevalence) among general and higher-risk populations alike.(11, 17, 18) Since HSV-2 is more transmissible than HIV and produces long-lasting antibodies, it has been used as an objective biological marker of sexual risk behavior and risk of HIV infection.(19–23) Analyses using empirical data and mathematical modeling supported the utility of using HSV-2 seroprevalence to predict HIV epidemic potential.(19, 20, 24)

Inadequate understating of HSV-2 epidemiology and its considerable consequences on sexual and reproductive health and the HIV epidemic,(11, 13) calls for urgent preventive and control measures to tackle it. The World Health Organization (WHO) outlined a “Global Health Sector Strategy on STIs”(25) that sets goals to eliminate STIs as a main public health concern by 2030 through integration of preventive and control measures. Consequently, WHO, along with their global partners, are spearheading efforts to develop an HSV vaccine as an urgent priority.(26, 27)

To inform these efforts, we conducted a systematic review to characterize HSV-2 infection levels and trends in sub-Saharan Africa (SSA), the hub of the HIV epidemic.(28, 29) We estimated pooled means for each outcome measure (incidence rate, seroprevalence, proportion of HSV-2 in GUD, and proportion of HSV-2 in genital herpes) across populations and subpopulations. We also conducted meta-regression analyses to assess temporal trends and identify predictors of high seroprevalence and between-study heterogeneity.

## Methodology

### Data sources and search strategy

This systematic review was guided by the Cochrane Collaboration Handbook(30) and was reported according to the Preferred Reporting Items for Systematic Reviews and Meta-analyses (PRISMA) guidelines(31) which can be found in Table S1. The review was informed by the methodology applied recently in a series of systematic reviews for HSV-1 infection.(32–36)

All available publications were systematically reviewed up to August 23, 2020. The search was conducted in PubMed and Embase databases, using search strategies with exploded MeSH/Emtree terms, broad search criteria, and no language or year restrictions to widen the scope and include all subheadings (Table S2). The definition of Africa included 45 countries, as defined by the WHO for the African Region,(37) covering countries of sub-Saharan Africa. The list of countries and their subregional stratification is in Box S1.

### Study selection and eligibility criteria

Search results were imported into the reference manager Endnote (Thomson Reuters, USA), whereby duplicate publications were identified and removed. The remaining records were screened for relevance based on titles and abstracts, followed by full text screening of relevant and potentially relevant records. Additional bibliography screening was performed on both reviews and the relevant articles to identify any missing publications.

Inclusion criteria were met if publications reported primary data on any of the following four outcomes: 1) HSV-2 incidence rate, 2) HSV-2 seroprevalence, 3) proportion of HSV-2 detection in GUD, and 4) proportion of HSV-2 detection in genital herpes. A sample size of ≥10 was required for inclusion for all outcome measures. Exclusion criteria encompassed case reports, series, commentaries, reviews, and qualitative studies. In this review, “publication” refers to a document reporting any outcome measure, whilst a “study” refers to details of a specific outcome measure. Special care was given to ensure that overlapping studies were only included once and not in duplicate.

### Data extraction and synthesis

Data extraction and double extraction were performed by MH and FA independently. A list of extracted variables is in Box S2.

Overall outcome measures and their stratified measures were extracted, provided the stratification agreed with a pre-set stratification hierarchy and the subsample in each stratum was ≥10. The pre-set stratification hierarchy sequence for incidence and seroprevalence measures was as follows: population type (see Box S3 for definition), sex, and age. As for proportion of HSV-2 detection in GUD and genital herpes, the sequence was: genital herpes episode status (primary *versus* recurrent episode), sex, age, and study site (hospital *versus* outpatient clinic).

Measures reporting any HSV-2 outcome among children <15 years old were only reported, and not included in the analyses.

### Quality assessment

Relevant studies were subjected to a pre-quality assessment to evaluate the validity of the assays used, given their limitations.(38, 39) This assessment was done with the help of an expert from the University of Washington, Professor Rhoda Ashley-Morrow. Only studies with valid, sensitive, and specific assays were included in the review. These studies were then evaluated using the Cochrane approach for risk of bias (ROB) assessment.(30) Study’s precision was classified into low *versus* high based on the study sample size (<100 *versus* ≥100).(40) Studies were classified into low *versus* high ROB using two quality domains: sampling method (probability *versus* non-probability based sampling) and response rate (≥80% *versus* <80% or unclear). Effect of ROB on study outcome was investigated through meta-regression as noted below.

### Meta-analyses

Pooled mean estimates for all four outcomes were calculated across all strata using meta-analyses, provided each stratum had ≥3 measures.

Only studies reporting incidence rate by person-time were included in the meta-analyses. Log transformed incidence rates were used to calculate pooled estimates using the inverse variance method in the metarate function.(41, 42)

HSV-2 seroprevalence measures and proportion of HSV-2 detection in GUD and in genital herpes were each pooled using the DerSimonian-Laird random-effects model,(41) applying the Freeman-Tukey double arcsine transformation to stabilize the variance,(43) and factoring knowledge of the applicability of this transformation.(44)

Existence of heterogeneity in effect size was assessed using Cochran’s Q statistic.(41, 45) Magnitude of between-study variation attributed to *true* difference in effect size, as opposed to *chance*, was measured using I^2^.(41, 45) Distribution of *true* measures around the pooled mean was described using the prediction interval.

Meta-analyses were conducted in R version 3.4.1(46) using the meta package.(42)

### Meta-regressions

Univariable and multivariable random-effects meta-regression analyses were conducted using log transformed seroprevalence measures to examine factors and predictors potentially associated with increased HSV-2 seroprevalence, as well as sources of between-study heterogeneity. Pre-set variables included in these analyses are listed in Box S4.

Factors included in the multivariable models had to have a p-value<0.1 in the univariable model. Strength of evidence for an association was deemed significant for factors with a p-value<0.05 in the multivariable models.

Meta-regressions were conducted in Stata/SE version 13(47) using the metareg package.(48)

## Results

### Search results and scope of evidence

Figure 1 depicts the PRISMA flow chart describing the study selection process.(31) A total of 14,830 citations were captured in the initial search (2,039 from PubMed and 12,791 from Embase). After deduplication, title and abstract screening, and full text screening, relevant publications were identified. Thirteen additional publications, including country level reports, were identified through bibliography screening.(49–61)

**Figure 1.**
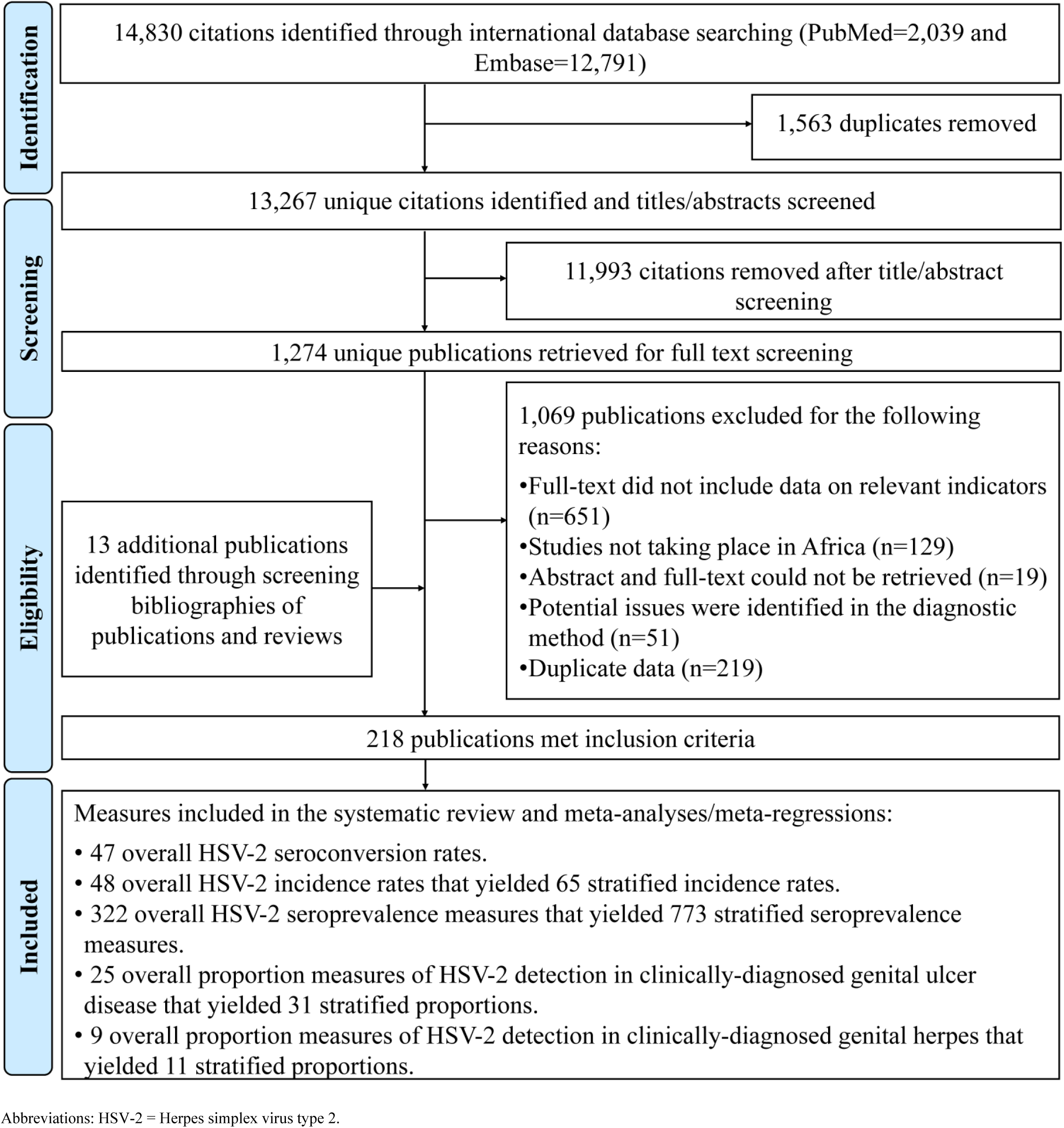
Flow chart of article selection for the systematic review of HSV-2 infection in sub-Saharan Africa, per the PRISMA guidelines.(31)

In total, 218 publications met the inclusion criteria. Extracted measures encompassed 47 seroconversion rates, 48 overall (65 stratified) incidence rates, 322 overall (773 stratified) seroprevalence measures, 25 overall (31 stratified) proportions of HSV-2 detection in GUD, and 9 overall (11 stratified) proportions of HSV-2 detection in genital herpes.

### Incidence overview and pooled mean estimates for HSV-2 incidence rate

Table S3 summarizes extracted seroconversion and incidence rates. Studies were either longitudinal cohorts (number of measures (n)=35; 49.2%) or randomized controlled trials (n=36; 50.7%), with a follow-up duration ranging between 6 weeks to 6 years. HSV-2 seroconversion rates (n=47) ranged between 0.0-58.0% across studies, reflecting in part the widely variable duration of follow-up.

In general populations, HSV-2 incidence rates among women (n=20) ranged between 3.6-21.7 per 100 person-years with a median of 7.5 and a pooled mean of 7.2 (95% confidence interval (CI): 5.5-9.4) per 100 person-years (Table 1). Among men (n=20), incidence rate ranged between 1.5-10.5 per 100 person-years with a median of 5.5 and a pooled mean of 4.1 (95% CI: 3.1-5.3) per 100 person-years. Higher incidence rates were found in higher risk populations. A summary of pooled mean incidence rate by population type and associated forest plots are in Table 1 and Figure S1, respectively.

**Table 1.** Pooled mean estimates for herpes simplex virus type 2 incidence rate among different populations in sub-Saharan Africa.

| Population type | Outcome measures | Samples | HSV-2 incidence rate (per 100 person-years) |  | Pooled mean HSV-2 incidence rate | Heterogeneity measures |  |
| --- | --- | --- | --- | --- | --- | --- | --- |
|  | Total N | Total person-years | Range | Median | Mean (per 100 person-years) (95% CI) | Q* (p-value) | I<sup>2</sup> (%) (95% CI) |
| General populations |  |  |  |  |  |  |  |
| Women | 20 | 21,607.4 | 3.6-21.7 | 7.5 | 7.2 (5.5-9.4) | 321.1 (p<0.001) | 94.1 (92.1-95.6) |
| <25 years old | 6 | 6,914.5 | 4.0-21.7 | 9.1 | 6.7 (4.6-9.7) | 51.5 (p<0.001) | 90.3 (81.6-94.9) |
| ≥25 years old | 3 | 154.0 | 9.0-15.4 | 9.3 | 11.4 (7.1-18.3) | 1.1 (p=0.582) | 0.0 (0.0-80.8) |
| Mixed ages | 11 | 14,538.9 | 3.6-21.0 | 7.0 | 6.9 (4.7-10.1) | 257.5 (p<0.001) | 96.1 (94.5-97.3) |
| Men | 20 | 29,287.3 | 1.5-10.5 | 5.5 | 4.1 (3.1-5.3) | 250.9 (p<0.001) | 92.4 (89.7-94.4) |
| <25 years old | 6 | 3,841.1 | 1.5-10.4 | 6.5 | 4.0 (2.3-7.0) | 49.9 (p<0.001) | 90.0 (80.9-94.7) |
| ≥25 years old | 8 | 1,835.2 | 4.0-10.5 | 6.5 | 6.5 (5.3-7.9) | 7.7 (p=0.364) | 8.6 (0.0-70.4) |
| Mixed age | 6 | 23,611.1 | 1.4-4.9 | 5.0 | 2.4 (1.6-3.6) | 120.0 (p<0.001) | 95.8 (93.1-97.5) |
| Mixed sexes | 5 | 15,603.7 | 2.3-3.6 | 3.5 | 3.2 (2.5-3.7) | 17.9 (p=0.001) | 77.7 (46.2-90.7) |
| Intermediate-risk populations |  |  |  |  |  |  |  |
| Women | 3 | 774.8 | 14.2-28.6 | 17.3 | 19.4 (11.8-32.1) | 16.8 (p<0.001) | 88.1 (66.9-95.7) |
| Higher-risk populations |  |  |  |  |  |  |  |
| Female sex workers | 3 | 1,458.0 | 8.7-23.0 | 21.0 | 18.0 (13.1-24.9) | 11.2 (p=0.004) | 82.2 (45.2-94.2) |
| HIV negative populations |  |  |  |  |  |  |  |
| Mixed sexes | 5 | 5,203.0 | 5.2-11.0 | 6.1 | 7.9 (6.0-10.5) | 31.7 (p<0.001) | 87.4 (73.0-94.1) |
| HIV positive individuals and in individuals in HIV discordant couples |  |  |  |  |  |  |  |
| Mixed sexes | 3 | 2,606.8 | 5.5-14.8 | 7.7 | 8.6 (4.7-15.7) | 38.2 (p<0.001) | 94.8 (88.1-97.7) |
| Other populations |  |  |  |  |  |  |  |
| Mixed sexes | 6 | 767.3 | 3.4-35.1 | 17.1 | 14.2 (8.6-23.6) | 27.7 (p<0.001) | 82.0 (61.6-91.5) |

\*Q: the Cochran's Q statistic is a measure assessing the existence of heterogeneity in incidence rates.
†I<sup>2</sup>: a measure that assesses the magnitude of between-study variation that is due to actual differences in incidence rates across studies rather than chance.
Abbreviations: CI = Confidence interval, HIV = human immunodeficiency virus, HSV-2 = Herpes simplex virus type 2.

### Prevalence overview

Tables S4, S5, S6, S7, and S8 summarize (the overall) seroprevalence measures (n=322) across the subregions of SSA. Half the studies were published after the year 2010 (n=176; 55.5%), and most were based on cross-sectional study design (n=202; 62.7%) and convenience sampling (n=233; 72.4%).

For the stratified seroprevalence measures, HSV-2 seroprevalence among general population women (n=290) ranged between 0.1-97.4% with a median of 43.1%, and among general population men (n=186) between 0.0-84.2% with a median of 27.5% (Table 2).

**Table 2.** Pooled mean estimates for herpes simplex virus type 2 seroprevalence among the different at risk populations in sub-Saharan Africa by sex.

| Population type | Outcome measures | Sample size | HSV-2 seroprevalence (%) |  | Pooled mean HSV-2 seroprevalence | Heterogeneity measures |  |  |
| --- | --- | --- | --- | --- | --- | --- | --- | --- |
|  | Total N | Total n | Range | Median | Mean (%) (95% CI) | Q* (p-value) | I<sup>2</sup> (%) (95% CI) | Prediction interval<sup>‡</sup> (%) |
| General populations | 507 | 230,541 | 0.0-97.4 | 39.5 | 37.3 (34.9-39.7) | 73,247.9 (p<0.001) | 99.3 (99.3-99.3) | 0.7-88.0 |
| Women | 290 | 134,034 | 0.1-97.4 | 43.1 | 43.1 (39.8-46.5) | 44,291.3 (p<0.001) | 99.3 (99.3-99.4) | 1.8-92.4 |
| Men | 186 | 83,898 | 0.0-84.2 | 27.5 | 29.1 (25.7-32.6) | 21,577.1 (p<0.001) | 99.1 (99.1-99.2) | 0.1-77.7 |
| Mixed | 31 | 12,609 | 1.3-90.9 | 33.9 | 32.5 (27.3-37.8) | 975.5 (p<0.001) | 96.9 (96.3-97.4) | 8.4-62.9 |
| Intermediate-risk populations | 45 | 9,259 | 0.0-92.2 | 58.0 | 57.1 (50.1-63.9) | 1,906.5 (p<0.001) | 97.7 (97.3-98.0) | 13.7-94.6 |
| Women | 29 | 8,267 | 20.5-92.2 | 70.6 | 66.4 (59.1-73.3) | 1,283.4 (p<0.001) | 97.8 (97.4-98.2) | 25.4-96.4 |
| Men | 13 | 849 | 9.1-88.5 | 43.0 | 43.6 (31.4-56.2) | 150.1 (p<0.001) | 92.0 (88.1-94.6) | 4.3-88.7 |
| Mixed | 3 | 143 | 0.0-45.0 | 21.0 | 18.2 (0.5-43.4) | 29.1 (p<0.001) | 93.1 (83.3-97.2) | 0.0-100 |
| Higher-risk populations | 40 | 13,476 | 4.3-99.0 | 63.7 | 61.4 (53.4-69.1) | 3,264.9 (p<0.001) | 98.8 (98.7-98.9) | 13.4-98.2 |
| FSWs | 39 | 13,036 | 4.3-99.0 | 65.0 | 62.5 (54.8-70.0) | 2,898.0 (p<0.001) | 98.7 (98.5-98.8) | 15.7-97.9 |
| MSM | 1<sup>§</sup> | 440 | - | - | 22.3 (18.5-26.3)<sup>§</sup> | - | - | - |
| HIV negative populations | 51 | 38,533 | 6.0-89.0 | 44.0 | 47.4 (43.2-51.5) | 3,198.9 (p<0.001) | 98.4 (98.2-98.6) | 19.7-75.9 |
| Women | 34 | 33,699 | 17.0-89.0 | 46.5 | 52.1 (47.5-56.7) | 2,283.9 (p<0.001) | 98.6 (98.3-98.7) | 25.6-78.0 |
| Men | 13 | 3,119 | 11.0-80.4 | 31.6 | 37.9 (30.2-46.0) | 227.1 (p<0.001) | 94.7 (92.5-96.3) | 10.9-69.8 |
| Mixed | 4 | 1,715 | 6.0-57.9 | 35.6 | 35.8 (23.0-49.8) | 84.4 (p<0.001) | 96.4 (93.5-98.1) | 0.0-93.1 |
| HIV positive individuals and in individuals in HIV discordant couples | 42 | 15,521 | 17.5-95.2 | 70.2 | 71.3 (66.5-75.9) | 1,514.7 (p<0.001) | 97.3 (96.8-97.7) | 38.9-94.9 |
| Women | 20 | 6,183 | 42.0-95.2 | 79.7 | 76.7 (70.1-82.7) | 587.4 (p<0.001) | 96.8 (95.9-97.5) | 43.2-97.9 |
| Men | 15 | 4,306 | 44.0-94.2 | 61.4 | 72.0 (63.7-79.6) | 311.0 (p<0.001) | 95.5 (93.9-96.7) | 36.2-96.8 |
| Mixed | 7 | 5,032 | 17.5-70.2 | 62.3 | 54.6 (47.4-61.8) | 108.9 (p<0.001) | 94.5 (91.0-96.6) | 31.0-77.1 |
| STI clinic attendees and symptomatic populations<sup>¶</sup> | 72 | 11,996 | 14.7-93.3 | 60.7 | 61.2 (56.5-65.9) | 1,826.8 (p<0.001) | 96.1 (95.6-96.6) | 22.9-93.0 |
| Women | 26 | 4,038 | 38.0-93.3 | 75.3 | 69.2 (62.7-75.4) | 393.3 (p<0.001) | 93.6 (91.8-95.1) | 35.2-94.7 |
| Men | 38 | 6,585 | 14.7-86.4 | 56.6 | 52.9 (46.9-58.8) | 791.4 (p<0.001) | 95.3 (94.3-96.1) | 18.8-85.6 |
| Mixed | 8 | 1,373 | 85.0-85.0 | 79.4 | 75.8 (68.5-82.5) | 35.9 (p<0.001) | 80.5 (62.3-89.9) | 51.8-93.7 |
| Other populations | 16 | 6,506 | 11.2-89.4 | 48.1 | 50.3 (41.9-58.7) | 546.4 (p<0.001) | 97.3 (96.5-97.9) | 17.4-58.7 |
| Women | 9 | 5,642 | 27.8-89.4 | 48.7 | 53.8 (45.6-61.9) | 260.2 (p<0.001) | 96.9 (95.6-97.9) | 24.8-81.5 |
| Men | 3 | 803 | 11.2-38.6 | 23.7 | 23.4 (10.5-39.4) | 48.2 (p<0.001) | 95.9 (91.0-98.1) | 0.0-100 |
| Mixed | 4 | 61 | 46.7-83.3 | 75.7 | 71.1 (54.4-85.5) | 5.1 (p=0.164) | 41.2 (0.0-80.2) | 13.7-100 |

\*Q: the Cochran's Q statistic is a measure assessing the existence of heterogeneity in seroprevalence.
†I<sup>2</sup>: a measure that assesses the magnitude of between-study variation that is due to actual differences in seroprevalence across studies rather than chance.
‡Prediction interval: a measure that estimates the distribution (95% interval) of true seroprevalence around the estimated mean.
§No meta-analysis was done due to the small number of studies (n < 3).
¶Symptomatic populations include patients with clinical manifestations related to an STI.
Abbreviations: CI = Confidence interval, FSWs = Female sex workers, HIV = Human immunodeficiency virus, HSV-2 = Herpes simplex virus type 2, MSM = Men who have sex with men, STI = Sexually transmitted disease.

In higher-risk populations (n=40), almost all studies were conducted among female sex workers (FSWs; n=39) with seroprevalence ranging between 4.3-99.0% with a median of 65.0% (Table 2). High seroprevalence was observed in HIV positive individuals and in individuals in HIV discordant couples, ranging between 42.0-95.2% with a median of 79.7% among women (n=20), and between 44.0-94.2% with a median of 61.4% among men (n=15).

Tables 2, 3, and S9 summarize HSV-2 seroprevalence measures for further populations and subpopulations, including by population type, country, subregion, age, sex, and year of publication.

**Table 3.** Pooled mean estimates for herpes simplex virus type 2 seroprevalence among general populations in sub-Saharan Africa.

| Population classification | Outcome measures | Sample size | HSV-2 seroprevalence (%) |  | Pooled mean HSV-2 seroprevalence | Heterogeneity measures |  |  |
| --- | --- | --- | --- | --- | --- | --- | --- | --- |
|  | Total n | Total N | Range | Median | Mean (%) (95% CI) | Q* (p-value) | I<sup>2</sup> (%) (95% CI) | Prediction interval<sup>‡</sup> (%) |
| **African countries** |  |  |  |  |  |  |  |  |
| Benin | 16 | 3,991 | 1.0-57.0 | 28.0 | 24.6 (17.1-32.9) | 447.1 (p<0.001) | 96.6 (95.6-97.4) | 1.1-63.5 |
| Burkina Faso | 14 | 5,932 | 4.1-40.5 | 20.3 | 19.5 (14.7-24.8) | 297.3 (p<0.001) | 95.6 (94.0-96.8) | 3.5-43.8 |
| Cameroon | 23 | 2,309 | 3.0-90.9 | 59.0 | 53.6 (40.9-66.2) | 804.9 (p<0.001) | 97.3 (96.6-97.8) | 2.4-99.6 |
| Ethiopia | 10 | 2,444 | 7.9-59.5 | 28.5 | 27.3 (16.8-39.3) | 277.4 (p<0.001) | 96.8 (95.4-97.7) | 0.2-73.7 |
| Kenya | 71 | 34,605 | 1.9-91.3 | 42.4 | 39.9 (34.3-45.9) | 8,018.6 (p<0.001) | 99.1 (99.1-99.2) | 3.0-86.2 |
| Malawi | 55 | 9,571 | 0.0-87.5 | 50.0 | 45.1 (38.2-52.0) | 2,192.8 (p<0.001) | 97.5 (97.2-97.8) | 4.5-90.2 |
| Nigeria | 14 | 2,205 | 8.7-61.3 | 29.3 | 28.5 (16.1-42.7) | 486.9 (p<0.001) | 97.3 (96.5-98.0) | 0.0-85.6 |
| South Africa | 79 | 44,449 | 1.5-92.5 | 31.0 | 34.1 (27.3-41.9) | 19,846.8 (p<0.001) | 99.6 (99.6-99.6) | 0.0-93.4 |
| Tanzania | 41 | 18,731 | 7.0-68.0 | 36.8 | 37.4 (33.7-41.1) | 1,008.2 (p<0.001) | 96.0 (95.3-96.7) | 16.1-61.6 |
| Uganda | 59 | 52,216 | 9.9-90.7 | 53.4 | 50.5 (44.2-56.8) | 12,120.9 (p<0.001) | 99.5 (99.5-99.6) | 8.5-92.1 |
| Zambia | 42 | 25,973 | 1.0-80.0 | 41.5 | 36.3 (28.2-44.8) | 7,909.4 (p<0.001) | 99.5 (99.4-99.5) | 0.3-88.7 |
| Zimbabwe | 42 | 20,530 | 0.1-71.0 | 30.6 | 26.0 (17.5-35.5) | 8,678.9 (p<0.001) | 99.5 (99.5-99.6) | 0.0-88.6 |
| Other countries<sup>§</sup> | 41 | 7,585 | 2.6-97.4 | 22.2 | 31.1 (24.5-38.1) | 1,564.8 (p<0.001) | 97.4 (97.0-97.8) | 0.1-77.3 |
| **African subregions** |  |  |  |  |  |  |  |  |
| Eastern Africa | 188 | 110,399 | 1.9-91.3 | 42.4 | 41.9 (38.4-45.3) | 25,111.0 (p<0.001) | 99.3 (99.2-99.3) | 4.9-85.6 |
| Southern Africa | 226 | 100,925 | 0.0-92.5 | 40.3 | 35.8 (31.6-40.0) | 43,422.5 (p<0.001) | 99.5 (99.5-99.5) | 0.0-92.9 |
| Western Africa | 68 | 16,005 | 1.0-97.4 | 21.9 | 25.0 (20.9-29.4) | 2,442.9 (p<0.001) | 97.3 (96.9-97.6) | 1.2-63.7 |
| Central Africa | 25 | 3,212 | 3.0-90.9 | 59.0 | 52.4 (40.4-64.3) | 1,065.0 (p<0.001) | 97.7 (97.3-98.1) | 2.6-98.9 |
| **Age group** |  |  |  |  |  |  |  |  |
| <20 years | 88 | 40,217 | 0.0-46.0 | 11.0 | 12.4 (10.6-14.3) | 2,419.1 (p<0.001) | 96.4 (96.0-96.8) | 1.0-32.6 |
| 20-30 years | 112 | 49,099 | 2.9-91.3 | 36.8 | 34.2 (30.7-37.8) | 7,583.3 (p<0.001) | 98.5 (98.4-98.6) | 5.0-72.7 |
| 30-40 years | 69 | 26,412 | 16.4-90.4 | 60.6 | 57.1 (54.8-63.3) | 3,210.2 (p<0.001) | 97.9 (97.6-98.1) | 24.6-89.2 |
| 40-50 years | 48 | 12,470 | 20.0-92.5 | 61.7 | 64.6 (59.5-69.5) | 1,445.1 (p<0.001) | 96.7 (96.2-97.2) | 29.8-92.5 |
| >50 years | 15 | 4,054 | 39.6-80.9 | 60.6 | 58.2 (50.3-65.9) | 321.3 (p<0.001) | 95.6 (94.1-96.8) | 26.3-86.8 |
| Mixed | 175 | 98,289 | 0.1-97.4 | 35.0 | 36.2 (32.1-40.3) | 31,073.2 (p<0.001) | 99.4 (99.4-99.5) | 0.4-87.7 |
| **Year of publication category** |  |  |  |  |  |  |  |  |
| ≤2005 | 124 | 27,633 | 1.0-90.9 | 45.5 | 42.1 (38.1-46.2) | 5,477.8 (p<0.001) | 97.8 (97.6-97.9) | 6.2-83.8 |
| 2005-2015 | 283 | 142,060 | 0.0-97.4 | 38.4 | 35.7 (32.6-38.9) | 42,499.0 (p<0.001) | 99.3 (99.3-99.3) | 0.6-86.0 |
| >2015 | 100 | 60,848 | 1.5-92.5 | 35.8 | 35.9 (30.1-41.8) | 23,039.3 (p<0.001) | 99.6 (99.5-99.6) | 0.0-90.9 |
| **All studies** | **507** | **230,541** | **0.0-97.4** | **39.5** | **37.3 (34.9-39.7)** | **73,247.9 (p<0.001)** | **99.3 (99.3-99.3)** | **0.7-88.0** |

\*Q: the Cochran's Q statistic is a measure assessing the existence of heterogeneity in seroprevalence.
†I<sup>2</sup>: a measure that assesses the magnitude of between-study variation that is due to actual differences in seroprevalence across studies rather than chance.
‡Prediction interval: a measure that estimates the distribution (95% interval) of true seroprevalence around the estimated mean.
§ Other countries: Chad, Cote D'Ivoire, Eritrea, Gabon, Gambia, Ghana, Mali, Namibia, Rwanda, Senegal.
Abbreviations: CI = Confidence interval, HSV-2 = Herpes simplex virus type 2.

### Pooled mean estimates for HSV-2 seroprevalence

Tables 2, 3, and S9 show results of seroprevalence meta-analyses across populations and subpopulations. Pooled mean seroprevalence was lowest at 37.3% (95% CI: 34.9-39.7%) in general populations (n=507), followed by 47.4% (95% CI: 43.2-51.5%) in HIV negative populations (n=51), 57.1% (95% CI: 50.1-63.9%) in intermediate-risk populations (n=45), 61.2% (95% CI: 56.5-65.9%) in STI clinic attendees and symptomatic populations (n=72), 61.4% (95% CI: 53.4-69.1%) in higher-risk populations (n=40; mainly FSWs), and 71.3% (95% CI: 66.5-75.9%) in HIV positive individuals and in individuals in HIV discordant couples (n=42; Table 2).

Among general populations, the pooled mean seroprevalence varied across African subregions (Table 3), with the lowest being 25.0% (95% CI: 20.9-29.4%) in Western Africa (n=68), followed by 35.8% (95% CI: 31.6-40.0%) in Southern Africa (n=226), 41.9% (95% CI: 38.4-45.3%) in Eastern Africa (n=188), and 52.4% (95% CI: 40.4-64.3%) in Central Africa (n=25). Similar hierarchy was observed across subregions for women and men, with women having consistently higher pooled mean seroprevalence compared to men (Table S9).

Across age groups (Table 3), pooled mean seroprevalence increased gradually from 12.4% (95% CI: 10.6-14.3%) in those <20 years-old (n=88), followed by 34.2% (95% CI: 30.7-37.8%) in those 20-30 years-old (n=112), 57.1% (95% CI: 54.8-63.3%) in those 30-40 years-old (n=69), 64.6% (95% CI: 59.5-69.5%) in those 40-50 years-old (n=48), to then decrease slightly to 58.2% (95% CI: 50.3-65.9%) in those >50 years-old. A similar trend was observed across age groups for each of women and men, but seroprevalence grow faster with age for young women (Table S9).

Heterogeneity was evident in almost all meta-analyses (p-value<0.001), and was confirmed by the wide prediction intervals (Tables 2, 3, and S9). Most heterogeneity was attributed to true variation in seroprevalence rather than chance (I^2^>50%). Forest plots for meta-analyses across African subregions stratified by population type are in Figure S2.

### Predictors of HSV-2 seroprevalence

Table 4 shows results of the meta-regression analyses for HSV-2 seroprevalence. Nine variables were eligible for inclusion in the multivariable model (p-value<0.1 in univariable analysis). Two multivariable models were conducted, one including year of publication as a categorical variable and one including it as a linear term.

**Table 4.**
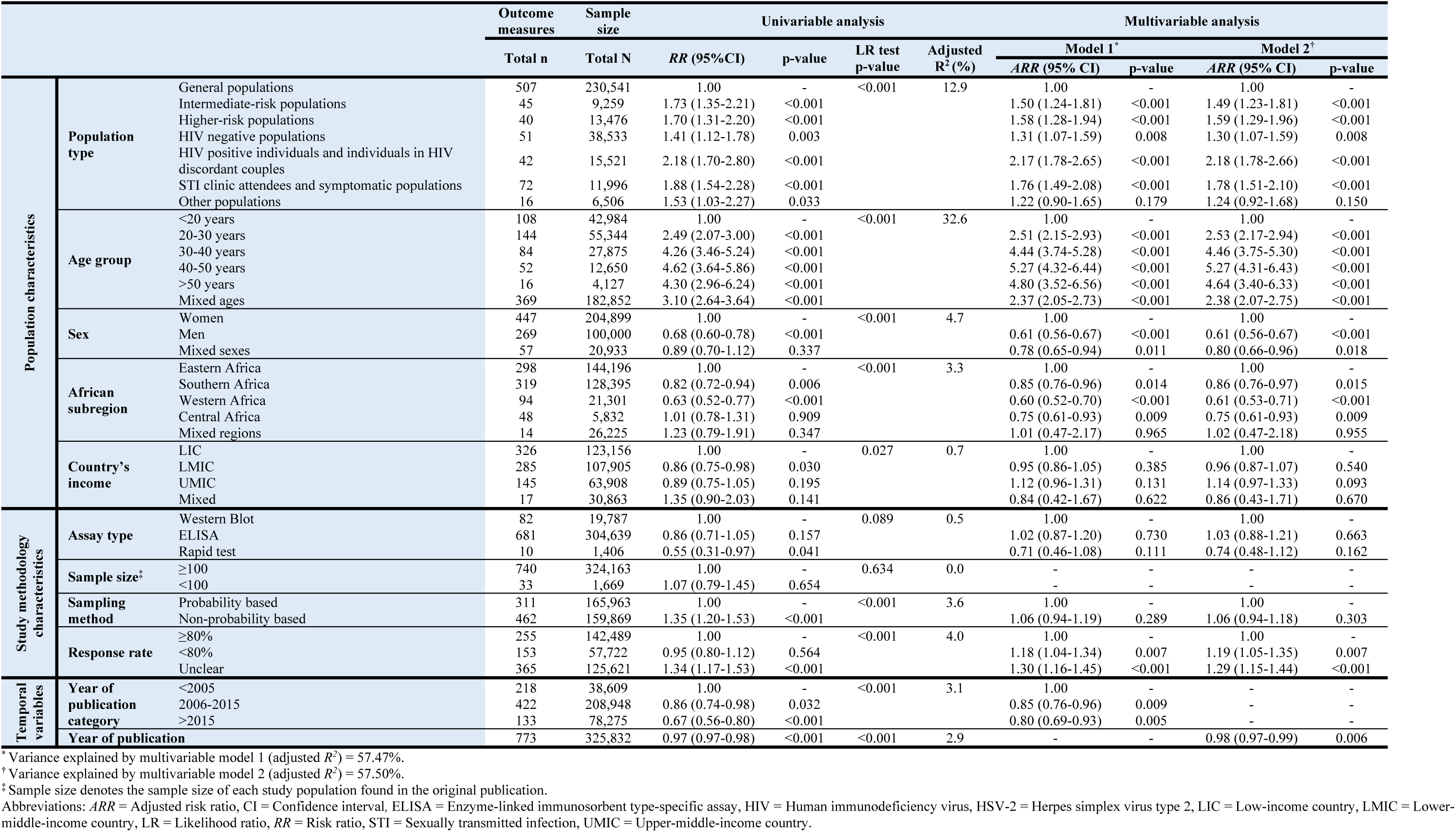
Univariable and multivariable meta-regression analyses for herpes simplex virus type 2 seroprevalence in sub-Saharan Africa.

The model including year of publication as a categorical variable explained 57.5% of seroprevalence variation and included population type, age group, sex, African subregion, country’s income, assay type, sampling method, response rate, and year of publication category (Table 4). Compared to general populations who had the lowest seroprevalence, seroprevalence was highest in HIV positive individuals and in individuals in HIV discordant couples [adjusted risk ratio (ARR) of 2.17 (95% CI: 1.78-2.65)], followed by STI clinic attendees and symptomatic populations, higher-risk populations, intermediate-risk populations, and HIV negative populations.

Compared to women, men had a 0.61-fold (95% CI: 0.56-0.67) lower seroprevalence. Seroprevalence increased rapidly with age at young age, but the increase plateaued by age 40-50 years. Seroprevalence was highest in Eastern Africa, followed by Southern Africa, Central Africa, and lowest in Western Africa. Country’s income was not associated with seroprevalence.

Studies that had a lower or unknown response rate had a higher seroprevalence. Meanwhile, assay type, study sample size, and study sampling method were not associated with seroprevalence.

Compared to those published before the year 2005, studies published in 2006-2015 [ARR of 0.85 (95% CI: 0.76-0.96)] and after 2015 [ARR of 0.80 (95% CI: 0.69-0.93)] had lower seroprevalence.

The model including year of publication as a linear term (Table 4) showed similar results and indicated declining seroprevalence with time [ARR of 0.98 (95% CI: 0.97-0.99)]. The model explained 57.5% of seroprevalence variation

Sensitivity analyses including year of data collection as a categorical variable or as a linear term (in replacement of year of publication) arrived at similar results (Table S10).

### Overview and meta-analyses of HSV-2 isolation in genital ulcer disease and in genital herpes

Table S11 summarizes extracted proportions of HSV-2 detection in GUD and in genital herpes. In GUD cases (n=31), proportion of HSV-2 detection ranged between 8.3-100% with a median of 49.1% and a pooled mean proportion of 50.7% (95% CI: 44.7-56.8%) (Table 5). Among women (n=8), it ranged between 35.0-100% with a median of 49.5% and a pooled proportion of 59.0% (95% CI: 44.7-72.6%), and among men (n=13), it ranged between 8.3-72.2% with a median of 49.1% and a pooled proportion of 47.3% (95% CI: 37.2-57.5%).

**Table 5.** Pooled proportions of herpes virus type 2 (HSV-2) virus isolation in clinically-diagnosed GUD and in clinically-diagnosed genital herpes in Africa.

| Population type | Outcome measures | Sample size | Proportion of HSV-2 isolation (%) |  | Pooled proportion of HSV-2 isolation (%) | Heterogeneity measures |  |  |
| --- | --- | --- | --- | --- | --- | --- | --- | --- |
|  | Total n | Total N | Range | Median | Mean (95% CI) | Q* (p-value) | I<sup>2</sup>† (%) (95% CI) | Prediction Interval‡ (%) |
| Patients with GUD | 31 | 4,296 | 8.3-100 | 49.1 | 50.7 (44.7-56.8) | 423.6 (p<0.001) | 92.9 (91.0-94.4) | 19.7-81.5 |
| Women | 8 | 816 | 35.0-100 | 49.5 | 59.0 (44.7-72.6) | 88.1 (p<0.001) | 92.1 (86.7-95.2) | 13.0-100 |
| Men | 13 | 1,915 | 8.3-72.2 | 49.1 | 47.3 (37.2-57.5) | 225.1 (p<0.001) | 94.7 (92.4-96.2) | 11.4-84.8 |
| Mixed sexes | 10 | 1,565 | 22.4-73.0 | 50.0 | 49.1 (39.0-59.2) | 110.3 (p<0.001) | 91.8 (87.1-94.8) | 15.5-83.2 |
| Patients with genital herpes | 11 | 1,380 | 53.0-100 | 100 | 97.3 (84.4-100) | 586.5 (p<0.001) | 98.3 (97.8-98.7) | 22.8-100 |
| Men | 6 | 715 | 53.0-100 | 100 | 97.3 (73.4-100) | 318.8 (p<0.001) | 98.4 (97.7-98.9) | 0.0-100 |
| Mixed sexes | 5 | 665 | 91.4-100 | 85.4 | 97.8 (91.9-100) | 23.9 (p<0.001) | 83.3 (62.0-92.6) | 66.2-100 |

\*Q: The Cochran's Q statistic is a measure assessing the existence of heterogeneity in pooled outcome measures, here proportions of HSV-2 virus isolation.
†I<sup>2</sup>: A measure assessing the magnitude of between-study variation that is due to true differences in proportions of HSV-2 virus isolation across studies rather than sampling variation.
‡Prediction interval: A measure quantifying the distribution 95% interval of true proportions of HSV-2 virus isolation around the estimated pooled mean.
Abbreviations: CI = Confidence interval, GUD = Genital ulcer disease, HSV-2 = Herpes simplex virus type 2.

In genital herpes cases (n=11), proportion of HSV-2 detection ranged between 53.0-100% with a median of 100% and a pooled mean proportion of 97.3% (95% CI: 84.4-100%) (Table 5). No study distinguished between primary and recurrent genital herpes episodes.

These meta-analyses showed evidence of heterogeneity (p-value<0.001, I²>50%, and wide prediction intervals). Forest plots are in Figure S3.

### Quality assessment

Quality assessment of diagnostic methods excluded 51 publications due to potential issues in the implemented diagnostic assays. Quality assessment of included seroprevalence measures is summarized in Table S12. Briefly, 288 studies (89.4%) had high precision, 88 studies (27.3%) and 85 studies (26.4%) had low ROB in the sampling method and in the response rate domains, respectively. Twenty-three studies (7.1%) had high ROB in both quality domains.

## Discussion

This systematic review presented a detailed assessment of HSV-2 epidemiology in sub-Saharan Africa. Strikingly, the results demonstrate that HSV-2 seroprevalence has been declining by about 2% per year over the last three decades (Table 4). This decline is in line with the observed declines in HIV epidemics in SSA during the same timeframe.(62) Drivers of the decline in HIV prevalence have been subject to debate, with various mechanisms posited including natural epidemic dynamics,(63) increased HIV-associated mortality,(64, 65) impact of interventions,(65) heterogeneity in host susceptibility to HIV infection,(66, 67) and reductions in sexual risk behavior.(65, 68–71) Considering that HSV-2 seroprevalence has been shown to provide an *objective proxy biomarker* of population-level sexual risk behavior,(19–24) our finding of rapidly declining HSV-2 seroprevalence suggests that sexual risk behavior has been declining, and that this decline has reduced the transmission of both HIV and HSV-2 infections. With evidence of HSV-2 infection increasing risk of HIV acquisition and transmission,(10–13) it remains to be seen whether the declining HSV-2 incidence may have also contributed to the declines seen in HIV incidence in SSA.

Despite declining HSV-2 transmission in SSA, incidence rate is still high (Table 1), and much higher than that found elsewhere in the world.(1) For instance, the incidence rate in the United States (<1 per 100 person-years)(72) is an order of magnitude lower than that in SSA. The results further demonstrate that age plays a profound effect in HSV-2 epidemiology—age alone accounted for 30% of the seroprevalence variation (Table 4). HSV-2 infection in SSA is typically acquired at a young age not long after sexual debut, especially so for women, manifesting in a rapidly increasing seroprevalence with age before plateauing at high levels by ages 40-50 (Tables 1, 3, 4, and S9).

Population risk classification was also found to play an important role in HSV-2 epidemiology, accounting for 13% of seroprevalence variation (Table 4). Both incidence rate and seroprevalence were found at much higher levels in specific at-risk populations, such as female sex workers (Tables 2 and 4), with both measures displaying the “classical hierarchy” of STI exposure by sexual risk behavior, also seen with other STIs.(73, 74) Despite its prominence amongst higher risk populations, HSV-2 infection is also widely disseminated in SSA with high levels of infection even in the lower risk general population where over a quarter of men and nearly half of women are infected (Tables 2 and 4).

Although HSV-2 seroprevalence is high everywhere in SSA, there are still considerable variations by subregion. Infection levels were highest in Eastern Africa followed by Southern Africa, Central Africa, and lowest in Western Africa (Table 4). Incidentally, this pattern is also seen for HIV infection with Eastern Africa and Southern Africa being most affected and Western Africa least affected.(75, 76) This further suggests a strong link between HSV-2 and HIV epidemiologies,(24) reflecting a similar mode of transmission and hinting at a biological/epidemiological synergy.(10–14) The results further demonstrate that women are almost twice as likely to be infected as men (Table 4), reflecting a higher bio-anatomical susceptibility to the infection.(77, 78)

Another finding of this study is that HSV-2 infection is the cause of half of GUD cases in SSA (Table 5), confirming that this infection is the main cause of this disease in this part of the world where nearly 60 million individuals are estimated to be affected with HSV-related GUD.(79)

Although HSV-2 seroprevalence is declining (Table 4), it will likely remain the main cause of GUD in SSA, as other causes such as syphilis have also been declining.(80, 81) HSV-2 infection (as opposed to HSV-1 infection) was also found to account for nearly all cases of genital herpes (>97%; Table 5). This finding is presumably due to the nature of HSV-1 being widely acquired in childhood in SSA through oral transmission,(33) and distinguishes this region from other global regions where there is an increasing role for HSV-1 in genital herpes,(33–36) with some countries already observing HSV-1 as the cause of a large proportion of cases of genital herpes.(33–36)

This study had limitations, principally the unavailability of data for 15 of 45 African countries. There was also less data for Central and Western Africa than for Eastern and Southern Africa, in addition to data for seroprevalence eclipsing those of GUD and genital herpes. Despite these limitations, a large volume of data was available to sufficiently power an array of analyses. Included studies exhibited heterogeneity (Tables 2, 3, and S9); however, more than half of this heterogeneity (57%) was subsequently explained through meta-regressions (Table 4). Studies differed by assay type, sample size, sampling method, and response rate, yet none of these study characteristics appeared to affect seroprevalence with the exception of response rate where studies with lower or unknown response rate had a higher seroprevalence (Table 4). Overall, these limitations should not pose a barrier to the critical interpretation of this study’s results, or its findings.

## Conclusions

In conclusion, HSV-2 seroprevalence is declining rapidly in SSA. Yet, HSV-2 incidence and seroprevalence remain at high levels, with over a third of the population being infected. Age and subregion within SSA play a critical role in HSV-2 epidemiology and explain much of the observed variation in seroprevalence. The geographical distribution of this infection was also found to be similar to that of HIV infection, and the declines in HSV-2 seroprevalence mirrored those for HIV prevalence. These findings suggest that the declines in both infections were driven by reductions in sexual risk behavior following the massive expansion of the HIV epidemic in this continent, and may suggest that some of the declines in HIV incidence could have been attributed to the declines in HSV-2 incidence. HSV-2 infection was found to be the etiological cause of half of GUD cases in this region, and virtually all cases of genital herpes. These findings demonstrate the urgent need for both prophylactic and therapeutic HSV-2 vaccines to tackle the disease burden of this infection,(82) and argue for further acceleration of ongoing efforts for vaccine development.(26, 27, 83)

## Data Availability

All relevant data are available within the manuscript and its supplementary materials.

## Contributors

MH and FAH conducted the systematic search, data extraction, and data analysis. MH wrote the first draft of the paper. CJ and KJL contributed to the systematic search, data extraction, and interpretation of the results. LJA conceived the study and led the data extraction and analysis and interpretation of the results. All authors contributed towards drafting and revising the manuscript.

## Acknowledgements

The authors gratefully acknowledge Professor Emeritus Rhoda Ashley Morrow from the University of Washington, for her support in assessing the quality of study diagnostic methods. The authors are also grateful for Ms. Adona Canlas for administrative support. The authors are grateful for pilot funding by the Biomedical Research Program and infrastructure support provided by the Biostatistics, Epidemiology, and Biomathematics Research Core, both at Weill Cornell Medicine-Qatar. This publication was made also possible by NPRP grant number 9-040-3-008 from the Qatar National Research Fund (a member of Qatar Foundation). The findings achieved herein are solely the responsibility of the authors. KL thanks the National Institute for Health Research, Health Protection Research Unit in Evaluation of Interventions at the University of Bristol, in partnership with Public Health England, for research support. The authors alone are responsible for the views expressed in this article and they do not necessarily represent the views, decisions or policies of the institutions with which they are affiliated, the NHS, the NIHR, the Department of Health and Social Care or Public Health England.

---

**Table S1.**
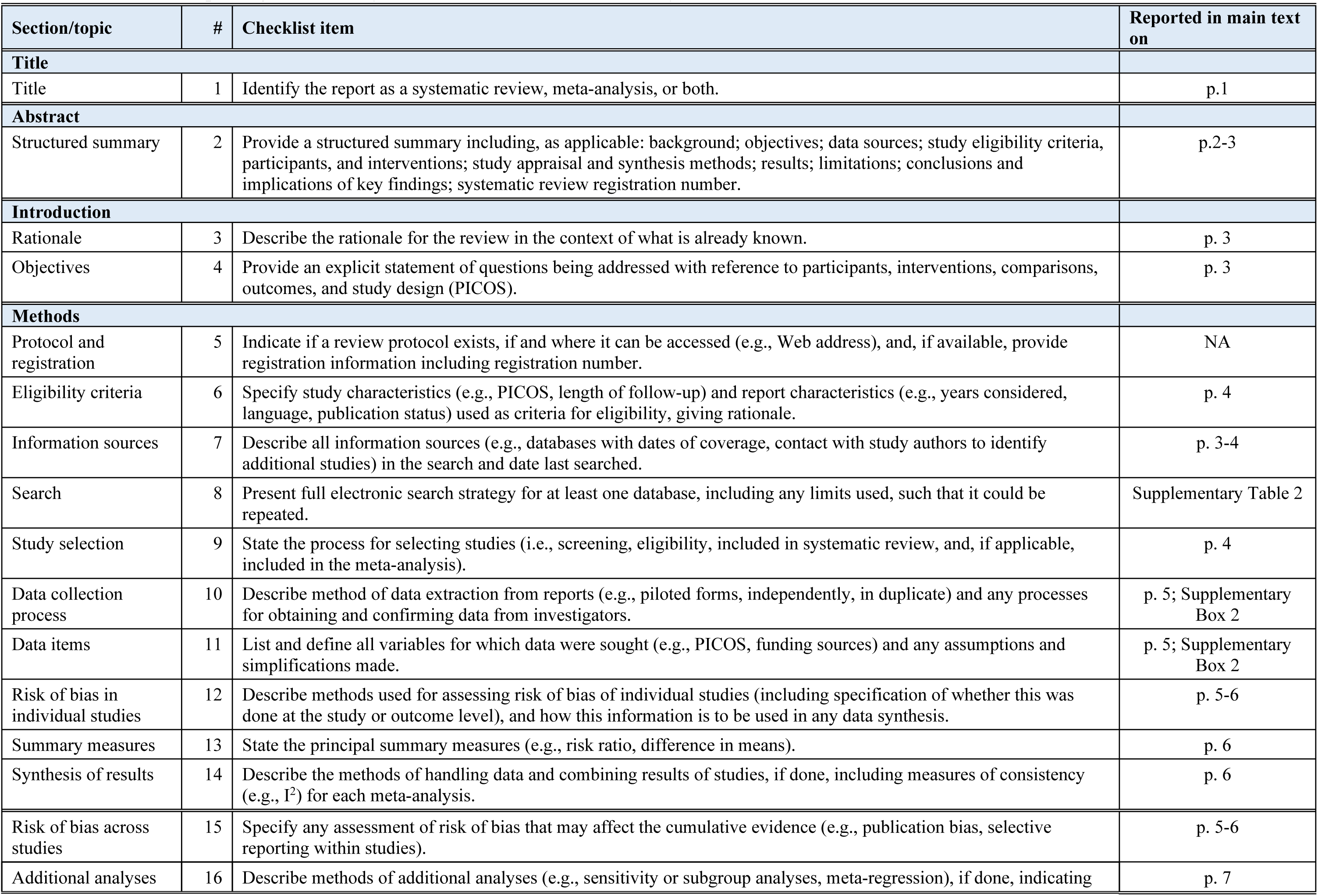
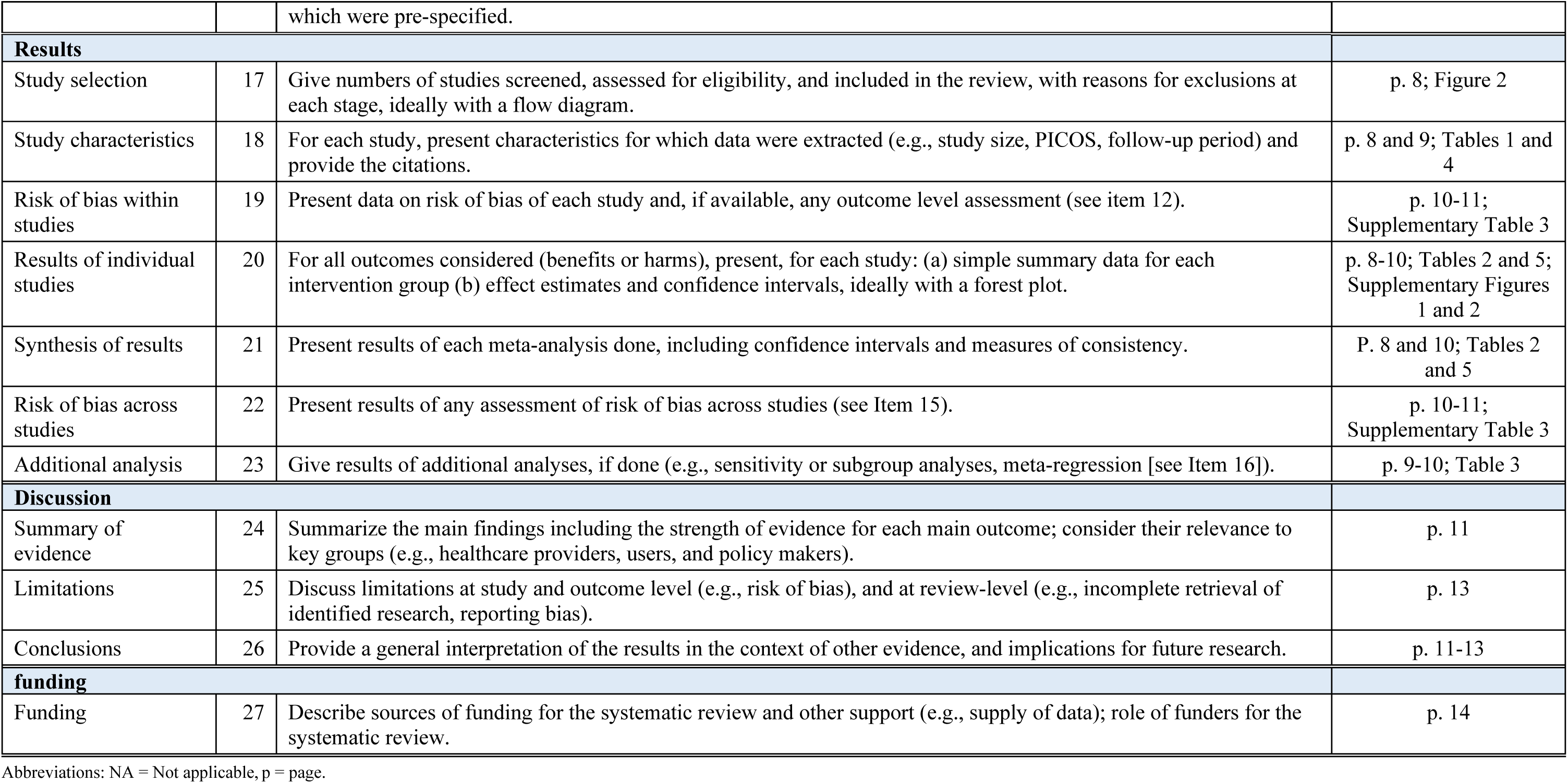
Preferred Reporting Items for Systematic Reviews and Meta-analyses (PRISMA) checklist.(1)

**Table S2.** Data sources and search criteria for systematically reviewing HSV-2 epidemiology in sub-Saharan Africa.

| **PubMed (last searched: August 23<sup>rd</sup>, 2020)** |
| --- |
| (Simplexvirus[MeSH] OR Herpes Simplex[MeSH] OR Herpes Genitalis[MeSH] OR Herpes Hominis[Text] OR HSV type-2[Text] OR HSV type 2[Text] OR HSV2[Text] OR HSV-2[Text] OR HSV [Text] OR Human herpes virus[Text] OR Herpes simplex virus type 2[Text] OR Herpes simplex virus type-2[Text] OR herpes simplex virus 2[Text] OR herpes simplex virus-2[Text] OR herpes simplex type 2[Text] OR herpes simplex type-2[Text] OR herpes simplex 2[Text] OR herpes simplex-2[Text] OR Herpesvirus type 2[Text] OR Herpesvirus type-2[Text] OR Herpesvirus 2[Text] OR Herpesvirus-2[Text] OR Herpes virus type 2[Text] OR Herpes virus type-[Text] OR Herpes virus [Text] OR Herpes virus-2[Text] OR genital herpes[Text] OR Herpes Genitalis[Text] OR Stomatitis Herpetic[Text] OR Herpes Labialis[Text]) AND (Africa South of the Sahara [MeSH] OR Comoros [MeSH] OR Ethiopia[MeSH] OR Madagascar[MeSH] OR Mauritius[MeSH] OR Sao Tome and Principe [MeSH] OR “Seychelles”[ MeSH] OR Angola*[Text] OR Benin*[Text] OR Botswan*[Text] OR Batswana[Text] OR Burkina fas*[Text] OR Burkina*[Text] OR Burundi*[Text] OR Cameroon*[Text] OR Cabo Verde*[Text] OR cape verd*[Text] OR Central Africa Republic[Text] Central Africa*[Text] OR Chad*[Text] OR Comor*[Text] OR Congo*[Text] OR Cote d’Ivoire[Text] OR Ivorian*[Text] OR Democratic Republic of Congo[Text] OR Equatorial Guinea*[Text] OR Equatoguinean*[Text] OR Eritr*[Text] OR Ethiop*[Text] OR Gabon*[Text] OR Gambia*[Text] OR Ghana*[Text] OR Ghinea*[Text] OR Guinea-Bissau[Text] OR Kenya*[Text] OR Lesotho*[Text] OR Basotho*[Text] OR Liberia*[Text] OR Madagascar*[Text] OR Malagasy*[Text] OR Malawi*[Text] OR Mali*[Text] OR Maurit*[Text] OR Mozambi*[Text] OR Namibia*[Text] OR Niger*[Text] OR Nigeria*[Text] OR Rwanda*[Text] OR Sao Tome and Principe*[Text] OR Sao Tome*[Text] OR Senegal*[Text] OR Seychell*[Text] OR Sierra Leone*[Text] OR South Africa*[Text] OR Swazi*[Text] OR Togo*[Text] OR Uganda*[Text] OR United Republic of Tanzania[Text] OR Tanzan*[Text] OR Zambia*[Text] OR Zimbabwe*[Text] OR Mauritania*[Text]) |
| **Embase (last searched: August 23<sup>rd</sup>, 2020)** |
| (exp Herpes simplex/ or exp Herpesviridae/) OR (Herpes simplex or Herpes simplex virus or HSV type-2 or HSV type 2 or HSV2 or HSV-2 or HSV 2 or human herpes virus or Herpes simplex virus type 2 or Herpes simplex virus type-2 or herpes simplex virus 2 or herpes simplex virus-2 or herpes simplex type 2 or herpes simplex type-2 or herpes simplex 2 or herpes simplex-2 or Herpesvirus type 2 or Herpesvirus type-2 or Herpesvirus 2 or Herpesvirus-2 or Herpes virus type 2 or Herpes virus type-2 or Herpes virus 2 or Herpes virus-2 or genital herpes or Herpes Genitalis or herpes labialis or herpetic stomatitis).mp.) AND (exp "Africa south of the Sahara" or exp Southern African/ or African/ or exp Central African/ or exp West African/ or exp South African/ or exp Central African Republic/ or exp East African/ or exp Mauritius/ or exp Mauritania/ or exp "Sao Tome and Principe"/ or exp Seychelles/) or (angola* or Benin* or Botswan* or Batswana* or Burkina Fas* or Burkina* or Burundi* or Cameroon* or Cabo verde* or Cape Verd* or Central Africa* or "Central African Republic*" or Chad* or Comor* or Congo* or Cote D'ivoire* or Ivorian* or "Democratic Republic of Congo*" or Equatorial Guinea* or Equatoguinean* or Ethiop* or Eritr* or Gabon* or Gambia* or Ghana* or Ghinea* or Guinea-Bissau* or Kenya* or Lesotho* or Basotho* or Liberia* or Madagascar* or Malagasy* or Malawi* or Mali* or Maurit* or Mozambi* or Namibia* or Niger* or Nigeria* or Rwanda* or "Sao Tome and Principe*" or Sao Tome* or Senegal* or Seychell* or Sierra Leone* or South Africa* or Swazi* or Togo* or Uganda* or United republic of Tanzania* or Tanza* or Zambia* or Zimbabwe* or Mauritania*).mp.) |

Abbreviations: HSV-2 = Herpes simplex virus type 2.
Abbreviations: HSV-2 = Herpes simplex virus type 2

## Box S1.

List of the 45 countries included in our definition for sub-Saharan Africa by subregion.(2)

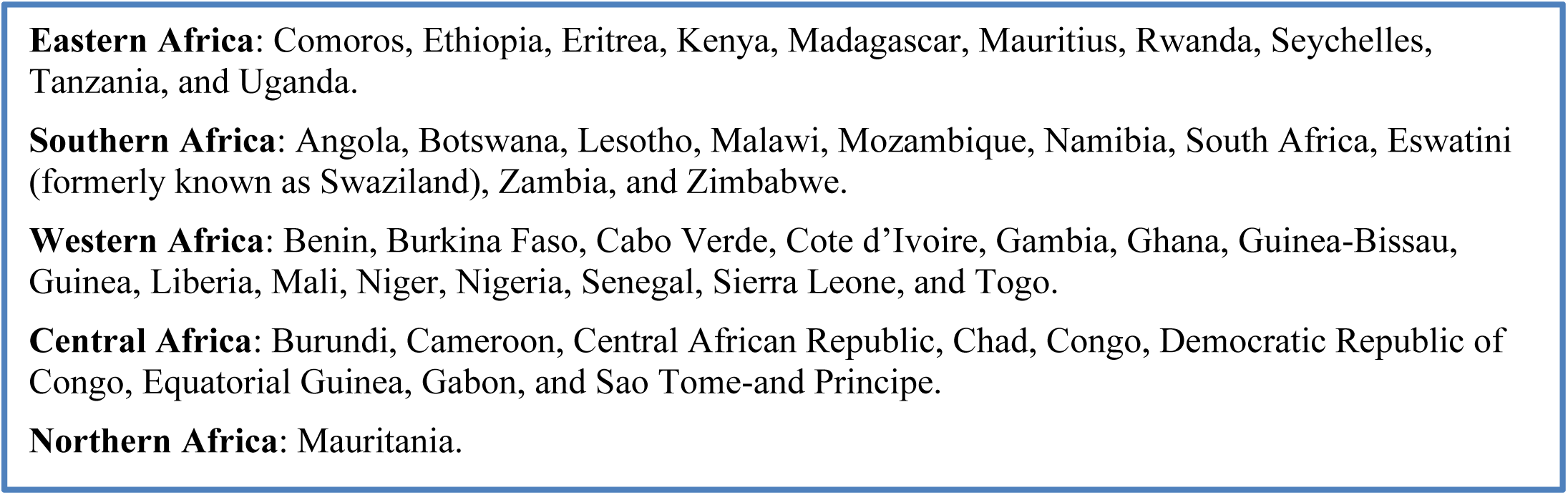

## Box S2.

List of variables extracted from the relevant publications meeting the inclusion criteria.

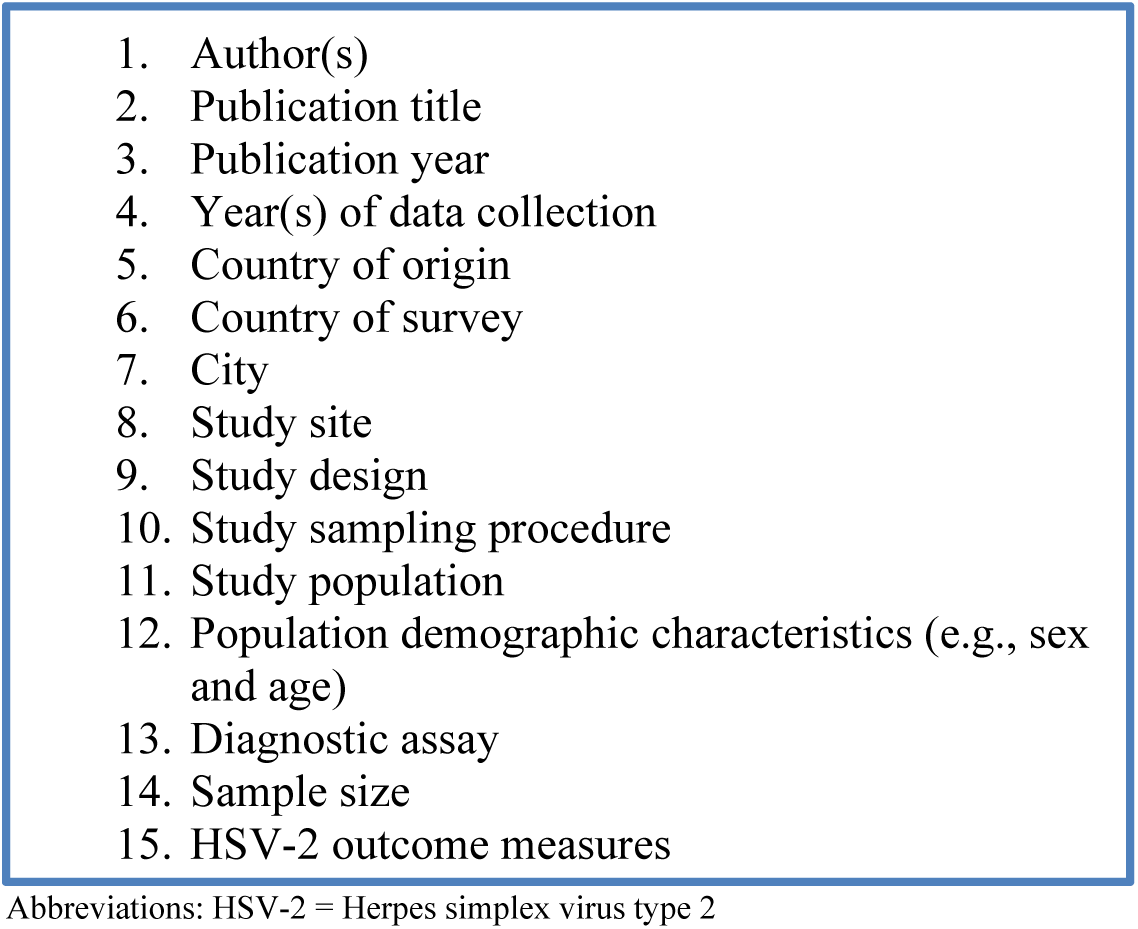

## Box S3.

Definitions of population type classifications.

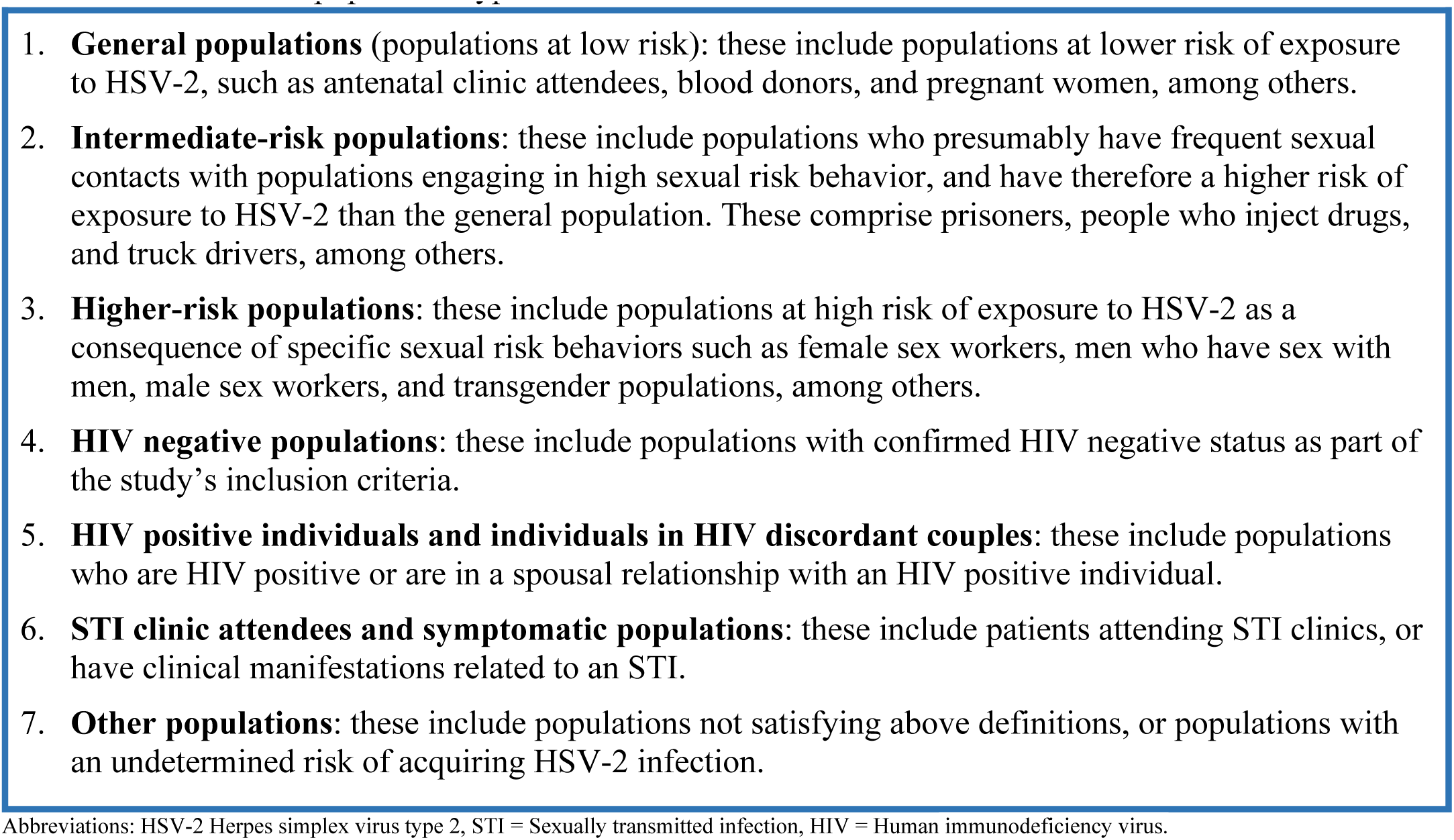

## Box S4.

Variables included in the univariable and multivariable meta-regression analyses.

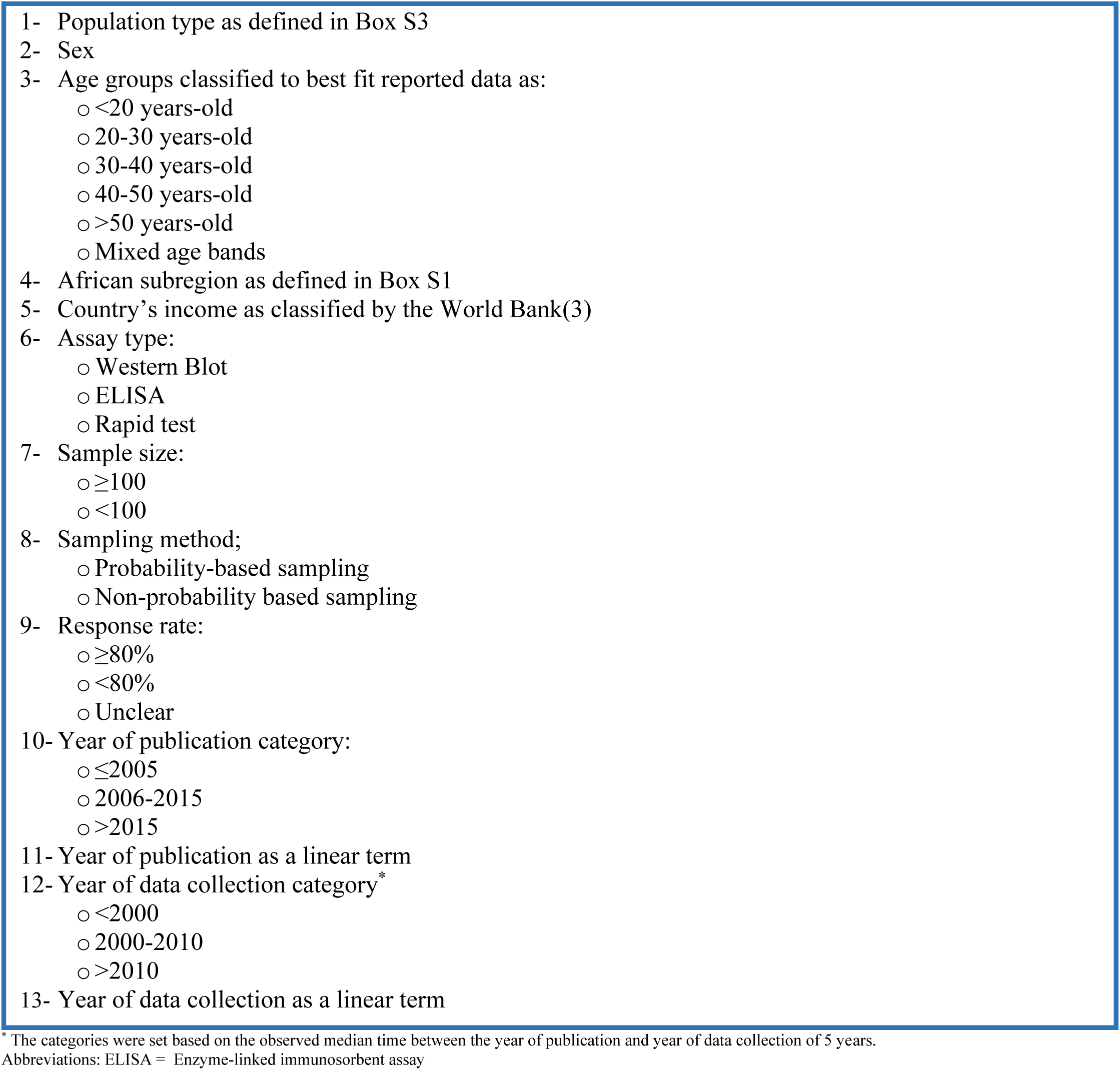

**Table S3.** Studies reporting HSV-2 seroconversion rate or incidence rate in sub-Saharan Africa.

| Author, year | Year(s) of data collection | Country | Original study design | Population characteristics | HSV-2 serological assay | Sample size | Follow-up duration | Person-years of follow-up | HSV-2 seroconversion rate (%) | HSV-2 incidence rate (per 100 person-years) |
| --- | --- | --- | --- | --- | --- | --- | --- | --- | --- | --- |
| **General populations** |  |  |  |  |  |  |  |  |  |  |
| Abdool Karim, 2015(4) | 2007-10 | South Africa | RCT | Women enrolled in a clinical trial | ELISA | 422 | 18 months | 561.3 | 20.6 | 15.5 |
| Akinyi, 2017(5) | 2007-10 | Kenya | Cohort | Participants in the "Incidence Cohort" study | ELISA | 673 | 1 year | - | - | 7.3 |
| Biraro, 2013(6) | 1990-94 | Uganda | Cohort | Male samples collected between 1990-1994 | ELISA | - | 18 months | 3,404.0 | - | 2.3 |
| Biraro, 2013(6) | 1995-99 | Uganda | Cohort | Male samples collected between 1995-1999 | ELISA | - | 18 months | 4,662.0 | - | 2.0 |
| Biraro, 2013(6) | 2000-04 | Uganda | Cohort | Male samples collected between 2000-2004 | ELISA | - | 18 months | 4,933.0 | - | 2.2 |
| Biraro, 2013(6) | 2005-07 | Uganda | Cohort | Male samples collected between 2005-2007 | ELISA | - | 18 months | 2,107.0 | - | 2.8 |
| Biraro, 2013(6) | 1990-94 | Uganda | Cohort | Female samples collected between 1990-1994 | ELISA | - | 18 months | 2,420.0 | - | 4.3 |
| Biraro, 2013(6) | 1995-99 | Uganda | Cohort | Female samples collected between 1995-1999 | ELISA | - | 18 months | 3,385.0 | - | 3.9 |
| Biraro, 2013(6) | 2000-04 | Uganda | Cohort | Female samples collected between 2000-2004 | ELISA | - | 18 months | 4,103.0 | - | 3.6 |
| Biraro, 2013(6) | 2005-07 | Uganda | Cohort | Female samples collected between 2005-2007 | ELISA | - | 18 months | 1,878.0 | - | 3.7 |
| De Baetselier, 2015(7) | 2009-10 | South Africa | Cohort | Females enrolled in an RCT | ELISA | 407 | 56 weeks | 229.5 | 13.4 | 18.3 |
| del Mar Pujades, 2002(8) | 1991-94 | Tanzania | RCT | Males enrolled in an RCT | ELISA | 221 | 2 years | - | 11.3 | - |
| del Mar Pujades, 2002(8) | 1991-94 | Tanzania | RCT | Females enrolled in an RCT | ELISA | 206 | 2 years | - | 17.5 | - |
| Hallfors, 2017(9) | 2011-14 | Kenya | RCT | Orphans in grades 7 and 8 | ELISA | 357 | 3 years | - | 30.8 | - |
| Heffron, 2011(10) | 2006-07 | Zambia | Cohort | Non-migrant male farmers | ELISA | 484 | 16 months | - | 6.8 | - |
| Jewkes, 2008(11) | 2003-04 | South Africa | RCT | Individuals in the intervention arm | ELISA | - | 2 years | 1,759.2 | - | 3.2 |
| Jewkes, 2008(11) | 2003-04 | South Africa | RCT | Individuals in the control arm | ELISA | - | 3 years | 1,623.3 | - | 4.6 |
| Kamali, 1999(12) | 1990-93 | Uganda | Cohort | 15-24 years old males and females | WB | 373 | 4 years | - | 20.9 | - |
| Kamali, 2003(13) | 1994-00 | Uganda | RCT | Individuals in intervention arm A | EIA | - | 6 years | 4,381.6 | - | 2.3 |
| Kamali, 2003(13) | 1994-00 | Uganda | RCT | Individuals in intervention arm B | EIA | - | 6 years | 4,595.5 | - | 3.6 |
| Kamali, 2003(13) | 1994-00 | Uganda | RCT | Individuals in control arm C | EIA | - | 6 years | 4,628.6 | - | 3.5 |
| Kebede, 2004(14) | 1997-02 | Ethiopia | Cohort | HSV-2 seronegative males and females | ELISA | 953 | 5 years | 3,225.5 | 58.0 | 1.8 |
| McFarland, 1999(15) | 1993-97 | Zimbabwe | Cohort | Male factory workers | WB | 1,444 | 4 years | 3,316.0 | 14.1 | 6.2 |
| Mensche, 2020(16) | 2007-13 | Malawi | Cohort | School students | ELISA | 2,072 | 6 years | - | 8.5 | - |
| Munjoma, 2010(17) | 2002-04 | Zimbabwe | Cohort | Pregnant women | ELISA | 173 | 10 months | 144.2 | 11.9 | 13.9 |
| Nakubulwa, 2016(18) | 2013-14 | Uganda | Cohort | Pregnant women | ELISA | 191 | - | - | 7.8 | - |
| Pettifor, 2016(19) | 2011-12 | South Africa | RCT | Individuals in the control arm | ELISA | 1,114 | 3 years | 2,525.0 | 9.1 | 4.0 |
| Pettifor, 2016(19) | 2011-12 | South Africa | RCT | Individuals in the intervention arm | ELISA | 1,214 | 3 years | 2,675.0 | 8.8 | 4.0 |
| Radebe, 2011(20) | 2008-10 | South Africa | RCT | Male students | ELISA | - | 54 months | - | - | 2.9 |
| Radebe, 2011(20) | 2008-10 | South Africa | RCT | Female students | ELISA | - | 54 months | - | - | 6.4 |
| Rosenberg, 2018(21) | 2013-15 | South Africa | Cohort | 18-25 years old women | ELISA | 645 | 2 years | - | 6.8 | - |
| Sobngwi, 2009(22) | 2002-04 | South Africa | RCT | Individuals in the control arm | ELISA | - | 21 months | 1,003.0 | - | 3.5 |
| Sobngwi, 2009(22) | 2002-04 | South Africa | RCT | Individuals in the intervention arm | ELISA | - | 21 months | 995.0 | - | 2.3 |
| Stoner, 2018(23) | 2011-15 | South Africa | Cohort | <20 years old young women | ELISA | 1,963 | 4 years | - | 5.9 | - |
| Tobian, 2009(24) | 2002-06 | Uganda | RCT | 15-49 years old men | ELISA | - | 2 years | 5,793.7 | - | 4.9 |
| Tobian, 2012(25) | 2002-07 | Uganda | RCT | Spouses of circumcised men | WB | 359 | 3 years | 656.5 | 11.1 | 6.1 |
| Tobian, 2012(25) | 2002-07 | Uganda | RCT | Spouses of uncircumcised men | WB | 363 | 4 years | 648.5 | 11.3 | 6.3 |
| van de Wijgert, 2009(26) | 1999-04 | Zimbabwe | Cohort | Zimbabwean women | ELISA | - | 5 years | - | - | 8.6 |
| Wagner, 1994(27) | - | Uganda | Cohort | Seronegative adults | WB | 36 | 1 year | - | 16.7 | - |
| **Intermediate-risk populations** |  |  |  |  |  |  |  |  |  |  |
| Kapiga, 2013(28) | 2008-10 | Tanzania | Cohort | Women working in food facilities and bars | ELISA | 450 | 1 year | 339.0 | 21.5 | 28.6 |
| Meque, 2014(29) | 2009-12 | Mozambique | Cohort | Women working in service facilities | ELISA | 151 | 1 year | - | 20.5 | - |
| Ondondo, 2014(30) | 2005-06 | Kenya | Cohort | Fishermen | ELISA | - | 1 year | - | 23.6 | - |
| Riedner, 2006(31) | 2000-02 | Tanzania | Cohort | Women working in a bar | EIA | - | 27 months | 98.0 | - | 17.3 |
| Tassiopoulos, 2007(32) | 2002-03 | Tanzania | Cohort | Individuals working in a hotel and bar | ELISA | 360 | 1 year | 337.8 | 13.3 | 14.2 |
| **Higher-risk populations** |  |  |  |  |  |  |  |  |  |  |
| Braunstein, 2011b(33) | 2007-09 | Rwanda | Cohort | FSWs in Rwanda | ELISA | 182 | 2 years | 150.0 | 1.9 | 8.7 |
| Chohan, 2009(34) | 1993-06 | Kenya | Cohort | FSWs in Kenya | ELISA | 297 | 28 months | 499.0 | 38.7 | 23.0 |
| Kaul, 2007(35) | 1998-02 | Kenya | RCT | FSWs in Kenya | ELISA | 121 | 4.4 years | - | 22.3 | - |
| Masese, 2014(36) | 1993-11 | Kenya | Cohort | FSWs in Kenya | ELISA | 406 | 5 years | 809.0 | 40.4 | 21.0 |
| Ramjee, 2005(37) | 1996-97 | South Africa | RCT | FSWs in South Africa | ELISA | 44 | 3.6 years | - | 54.9 | - |
| Traore, 2013(38) | 2009-11 | Burkina Faso | RCT | FSWs in Burkina Faso | ELISA | - | 1 year | - | - | 11.0 |
| **HIV negative populations** |  |  |  |  |  |  |  |  |  |  |
| De Bruyn, 2011(39) | 2002-05 | South Africa | RCT | Women in Durban | ELISA | 495 | 2 years | 689.0 | 15.4 | 11.0 |
| De Bruyn, 2011(39) | 2002-05 | South Africa | RCT | Women in Johannesburg | ELISA | 328 | 3 years | 443.0 | 10.4 | 7.7 |
| De Bruyn, 2011(39) | 2002-05 | Zimbabwe | RCT | Women in Harare | ELISA | 1,193 | 4 years | 949.0 | 8.4 | 5.2 |
| McCormack, 2010(40) | 2005-08 | 4 African countries* | RCT | Females in the intervention arm receiving 2% gel | ELISA | 297 | 1 year | - | 11.5 | - |
| McCormack, 2010(40) | 2005-09 | 4 African countries* | RCT | Females in the intervention arm receiving 0.5% gel | ELISA | 919 | 1 year | - | 11.9 | - |
| McCormack, 2010(40) | 2005-09 | 4 African countries* | RCT | Females in the control arm | ELISA | 888 | 1 year | - | 13.0 | - |
| Mehta, 2012(41) | 2002-05 | Kenya | RCT | Circumcised men | ELISA | 986 | 2 years | 1,493.5 | 8.7 | 5.8 |
| Mehta, 2012(41) | 2002-05 | Kenya | RCT | Uncircumcised men | ELISA | 1,035 | 2 years | 1,628.5 | 9.7 | 6.1 |
| Mehta, 2013(42) | 2002-07 | Kenya | RCT | Men enrolled in an RCT | ELISA | 2,044 | 6 years | - | 33.5 | - |
| Mlisana, 2012(43) | 2004-05 | South Africa | Cohort | HIV Negative women | ELISA | - | 2 years | - | - | 26.0 |
| Perti, 2014(44) | - | South Africa | Cohort | HIV negative pregnant women | WB | 91 | 6 weeks | - | 0.0 | - |
| **HIV positive individuals and individuals in HIV discordant couples** |  |  |  |  |  |  |  |  |  |  |
| Celum, 2014(45) | 2008-11 | Kenya and Uganda | RCT | Partners of HIV positive persons in an RCT | WB | 1,041 | 2 years | 1,422.2 | 7.6 | 5.6 |
| Celum, 2014(45) | 2008-11 | Kenya and Uganda | RCT | Partners of HIV positive persons in an RCT | WB | 481 | 2 years | 672.0 | 10.8 | 7.7 |
| Cowan, 2008a(46) | 1997-00 | Zimbabwe | RCT | HIV positive mothers with HIV positive infants | WB | 50 | 6 weeks | - | 22.0 | - |
| Cowan, 2008a(46) | 1997-00 | Zimbabwe | RCT | HIV positive mothers with HIV negative infants | WB | 106 | 6 weeks | - | 15.1 | - |
| del Mar Pujades, 2002(8) | 1991-94 | Tanzania | RCT | HIV positive females enrolled in a large RCT | ELISA | 32 | 2 years | - | 38.7 | - |
| del Mar Pujades, 2002(8) | 1991-94 | Tanzania | RCT | HIV positive Males enrolled in a large RCT | ELISA | 22 | 2 years | - | 31.8 | - |
| Muiru, 2013(47) | 2007-09 | Kenya | Cohort | Participants from HIV discordant couples | ELISA | 382 | 2 years | 512.6 | 19.8 | 14.8 |
| **Other populations** |  |  |  |  |  |  |  |  |  |  |
| Okuku, 2011(48) | 2005-08 | Kenya | Cohort | Women from different risk populations | ELISA | 164 | 4 years | - | 23.8 | 22.1 |
| Okuku, 2011(48) | 2005-08 | Kenya | Cohort | Men from different risk populations | ELISA | 443 | 4 years | - | 11.9 | 9.0 |
| van de Wiggert, 2009(26) | 1999-04 | Uganda | Cohort | Women from different risk populations | ELISA | - | 4 years | - | - | 10.6 |

\*The four African countries were: South Africa, Tanzania, Uganda, and Zambia.
Abbreviations: EIA = Enzyme immunoassay, ELISA = Enzyme-linked immunosorbent assay, FSWs = Female sex workers, HIV = Human immunodeficiency virus, HSV-2 = Herpes simplex virus type 2, RCT = Randomized controlled trial, WB = Western blot.

**Figure S1.**
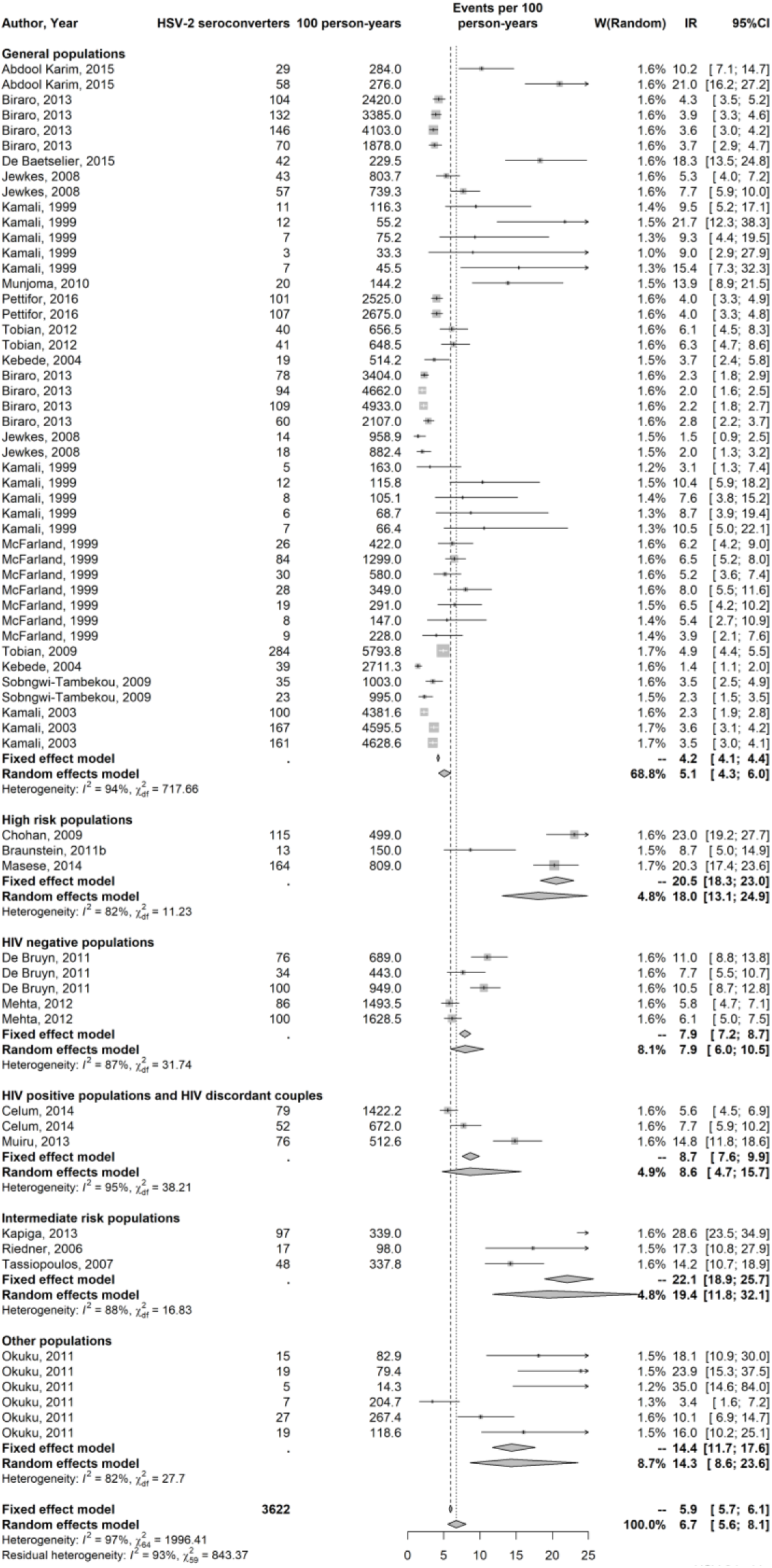
Forest plot presenting the outcome of the pooled mean herpes simplex virus type 2 (HSV-2) incidence rate by population type in sub-Saharan Africa.

**Table S4.**
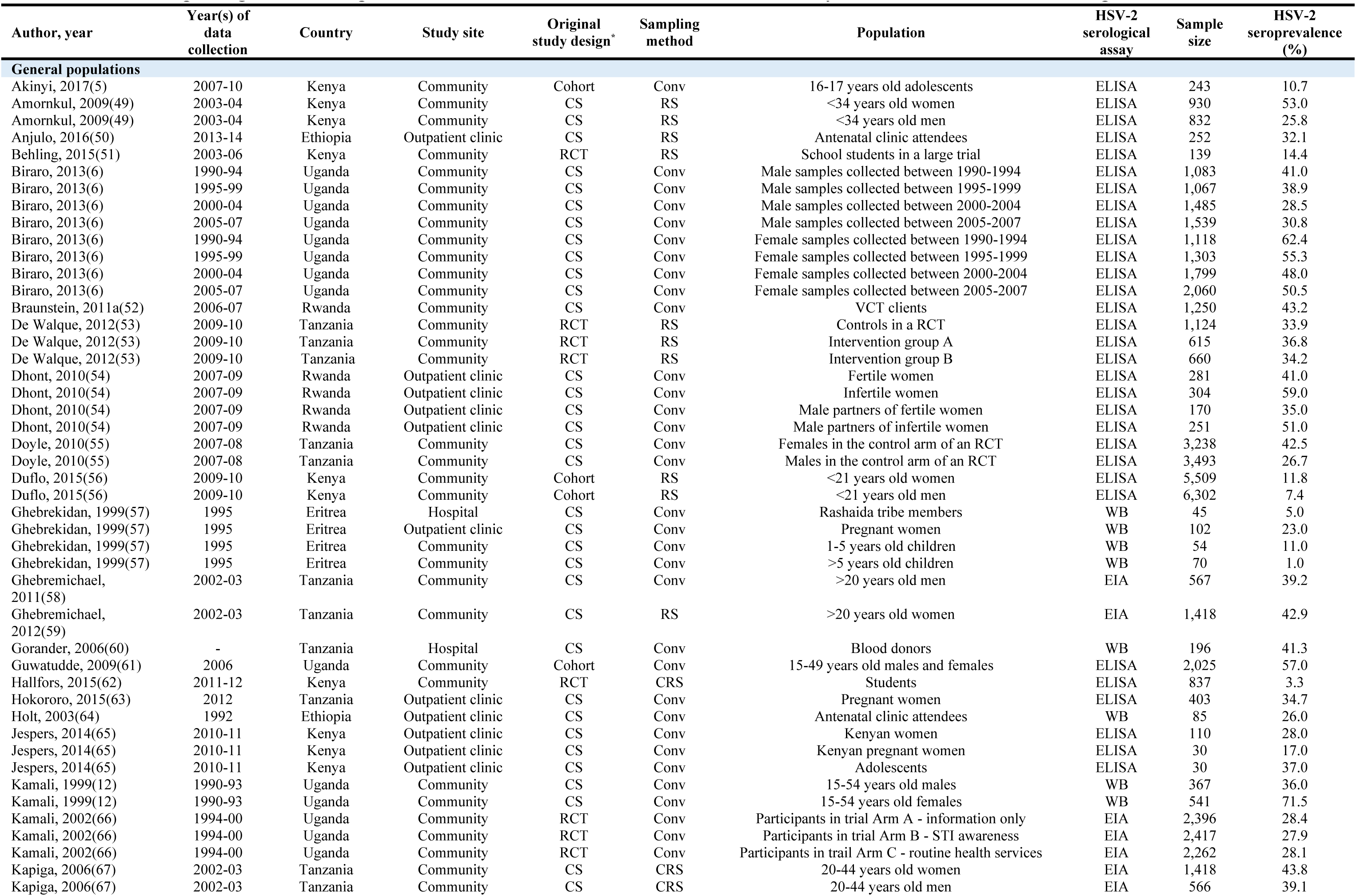
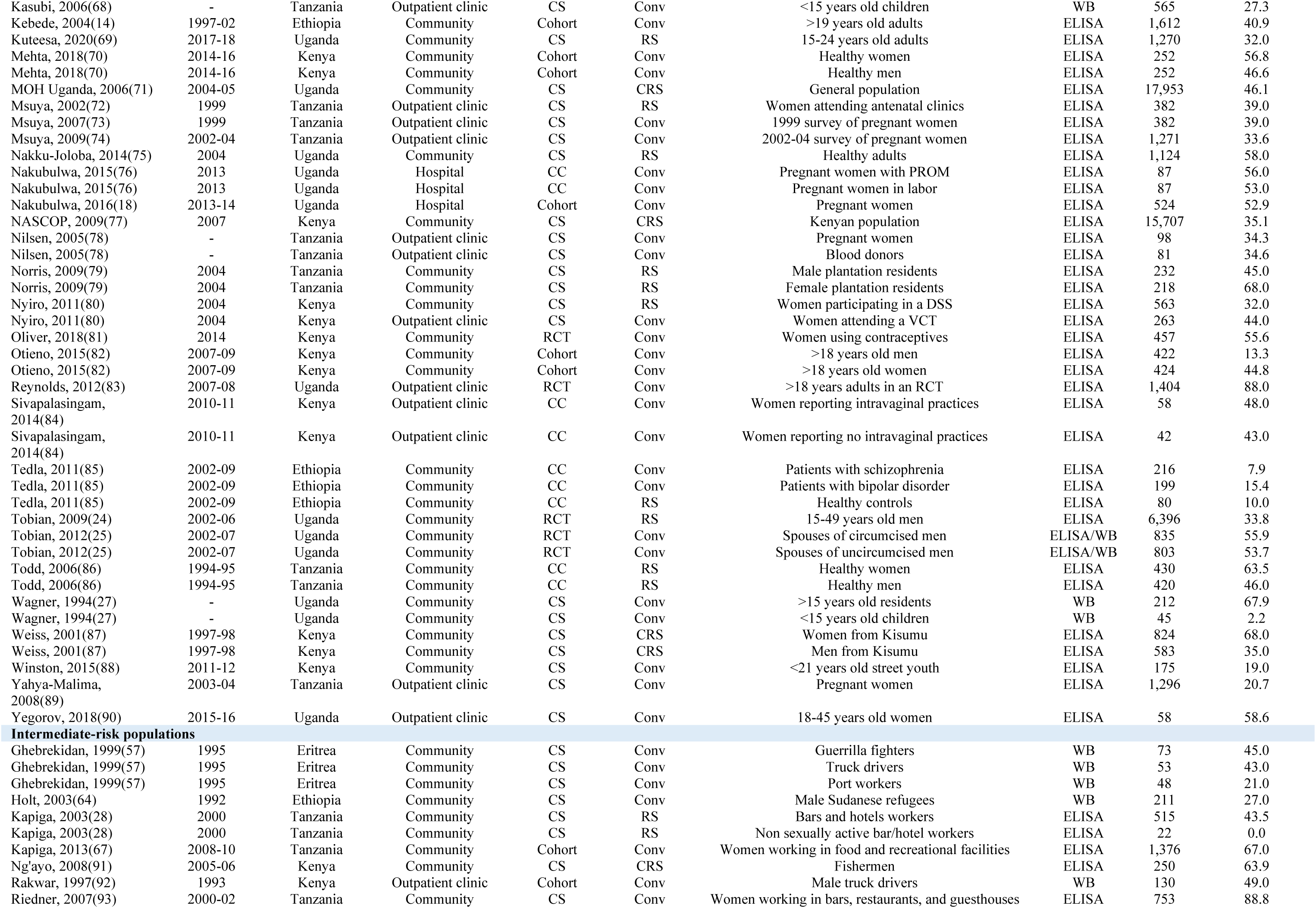
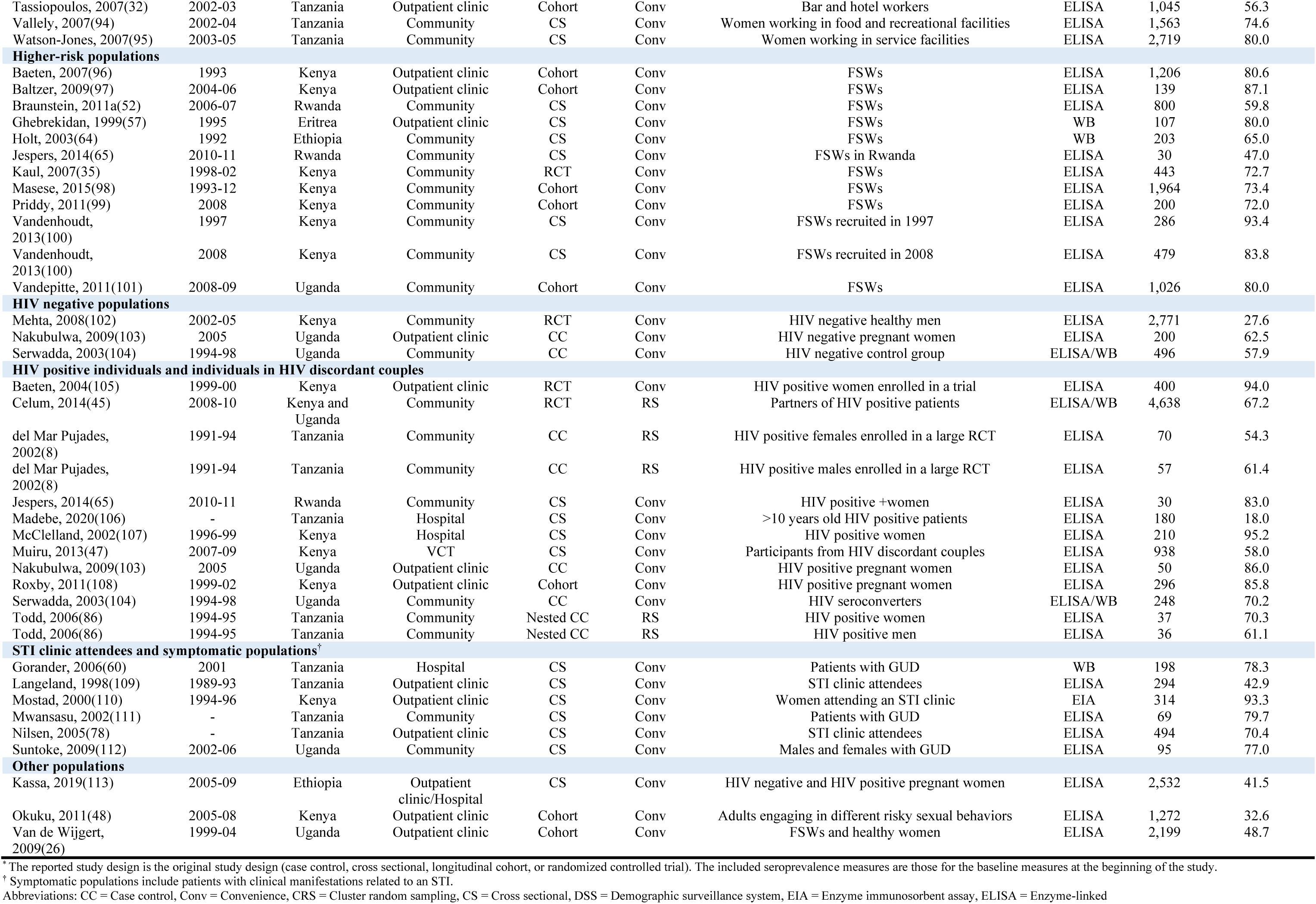
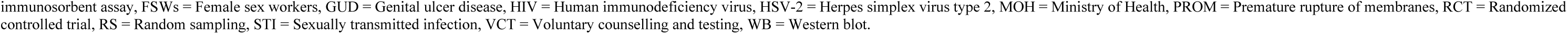
Studies reporting HSV-2 seroprevalence in Eastern Africa. This table includes only overall and not stratified seroprevalence measures.

| Author, year | Year(s) of data collection | Country | Study site | Original study design* | Sampling method | Population | HSV-2 serological assay | Sample size | HSV-2 seroprevalence (%) |
| --- | --- | --- | --- | --- | --- | --- | --- | --- | --- |
| **General populations** |  |  |  |  |  |  |  |  |  |
| Akinyi, 2017(5) | 2007-10 | Kenya | Community | Cohort | Conv | 16-17 years old adolescents | ELISA | 243 | 10.7 |
| Amornkul, 2009(49) | 2003-04 | Kenya | Community | CS | RS | <34 years old women | ELISA | 930 | 53.0 |
| Amornkul, 2009(49) | 2003-04 | Kenya | Community | CS | RS | <34 years old men | ELISA | 832 | 25.8 |
| Anjulo, 2016(50) | 2013-14 | Ethiopia | Outpatient clinic | CS | RS | Antenatal clinic attendees | ELISA | 252 | 32.1 |
| Behling, 2015(51) | 2003-06 | Kenya | Community | RCT | RS | School students in a large trial | ELISA | 139 | 14.4 |
| Biraro, 2013(6) | 1990-94 | Uganda | Community | CS | Conv | Male samples collected between 1990-1994 | ELISA | 1,083 | 41.0 |
| Biraro, 2013(6) | 1995-99 | Uganda | Community | CS | Conv | Male samples collected between 1995-1999 | ELISA | 1,067 | 38.9 |
| Biraro, 2013(6) | 2000-04 | Uganda | Community | CS | Conv | Male samples collected between 2000-2004 | ELISA | 1,485 | 28.5 |
| Biraro, 2013(6) | 2005-07 | Uganda | Community | CS | Conv | Male samples collected between 2005-2007 | ELISA | 1,539 | 30.8 |
| Biraro, 2013(6) | 1990-94 | Uganda | Community | CS | Conv | Female samples collected between 1990-1994 | ELISA | 1,118 | 62.4 |
| Biraro, 2013(6) | 1995-99 | Uganda | Community | CS | Conv | Female samples collected between 1995-1999 | ELISA | 1,303 | 55.3 |
| Biraro, 2013(6) | 2000-04 | Uganda | Community | CS | Conv | Female samples collected between 2000-2004 | ELISA | 1,799 | 48.0 |
| Biraro, 2013(6) | 2005-07 | Uganda | Community | CS | Conv | Female samples collected between 2005-2007 | ELISA | 2,060 | 50.5 |
| Braunstein, 2011a(52) | 2006-07 | Rwanda | Community | CS | Conv | VCT clients | ELISA | 1,250 | 43.2 |
| De Walque, 2012(53) | 2009-10 | Tanzania | Community | RCT | RS | Controls in a RCT | ELISA | 1,124 | 33.9 |
| De Walque, 2012(53) | 2009-10 | Tanzania | Community | RCT | RS | Intervention group A | ELISA | 615 | 36.8 |
| De Walque, 2012(53) | 2009-10 | Tanzania | Community | RCT | RS | Intervention group B | ELISA | 660 | 34.2 |
| Dhont, 2010(54) | 2007-09 | Rwanda | Outpatient clinic | CS | Conv | Fertile women | ELISA | 281 | 41.0 |
| Dhont, 2010(54) | 2007-09 | Rwanda | Outpatient clinic | CS | Conv | Infertile women | ELISA | 304 | 59.0 |
| Dhont, 2010(54) | 2007-09 | Rwanda | Outpatient clinic | CS | Conv | Male partners of fertile women | ELISA | 170 | 35.0 |
| Dhont, 2010(54) | 2007-09 | Rwanda | Outpatient clinic | CS | Conv | Male partners of infertile women | ELISA | 251 | 51.0 |
| Doyle, 2010(55) | 2007-08 | Tanzania | Community | CS | Conv | Females in the control arm of an RCT | ELISA | 3,238 | 42.5 |
| Doyle, 2010(55) | 2007-08 | Tanzania | Community | CS | Conv | Males in the control arm of an RCT | ELISA | 3,493 | 26.7 |
| Duflo, 2015(56) | 2009-10 | Kenya | Community | Cohort | RS | <21 years old women | ELISA | 5,509 | 11.8 |
| Duflo, 2015(56) | 2009-10 | Kenya | Community | Cohort | RS | <21 years old men | ELISA | 6,302 | 7.4 |
| Ghebrekidan, 1999(57) | 1995 | Eritrea | Hospital | CS | Conv | Rashaida tribe members | WB | 45 | 5.0 |
| Ghebrekidan, 1999(57) | 1995 | Eritrea | Outpatient clinic | CS | Conv | Pregnant women | WB | 102 | 23.0 |
| Ghebrekidan, 1999(57) | 1995 | Eritrea | Community | CS | Conv | 1-5 years old children | WB | 54 | 11.0 |
| Ghebrekidan, 1999(57) | 1995 | Eritrea | Community | CS | Conv | >5 years old children | WB | 70 | 1.0 |
| Ghebremichael, 2011(58) | 2002-03 | Tanzania | Community | CS | Conv | >20 years old men | EIA | 567 | 39.2 |
| Ghebremichael, 2012(59) | 2002-03 | Tanzania | Community | CS | RS | >20 years old women | EIA | 1,418 | 42.9 |
| Gorander, 2006(60) | - | Tanzania | Hospital | CS | Conv | Blood donors | WB | 196 | 41.3 |
| Guwatudde, 2009(61) | 2006 | Uganda | Community | Cohort | Conv | 15-49 years old males and females | ELISA | 2,025 | 57.0 |
| Hallfors, 2015(62) | 2011-12 | Kenya | Community | RCT | CRS | Students | ELISA | 837 | 3.3 |
| Hokororo, 2015(63) | 2012 | Tanzania | Outpatient clinic | CS | Conv | Pregnant women | ELISA | 403 | 34.7 |
| Holt, 2003(64) | 1992 | Ethiopia | Outpatient clinic | CS | Conv | Antenatal clinic attendees | WB | 85 | 26.0 |
| Jespers, 2014(65) | 2010-11 | Kenya | Outpatient clinic | CS | Conv | Kenyan women | ELISA | 110 | 28.0 |
| Jespers, 2014(65) | 2010-11 | Kenya | Outpatient clinic | CS | Conv | Kenyan pregnant women | ELISA | 30 | 17.0 |
| Jespers, 2014(65) | 2010-11 | Kenya | Outpatient clinic | CS | Conv | Adolescents | ELISA | 30 | 37.0 |
| Kamali, 1999(12) | 1990-93 | Uganda | Community | CS | Conv | 15-54 years old males | WB | 367 | 36.0 |
| Kamali, 1999(12) | 1990-93 | Uganda | Community | CS | Conv | 15-54 years old females | WB | 541 | 71.5 |
| Kamali, 2002(66) | 1994-00 | Uganda | Community | RCT | Conv | Participants in trial Arm A - information only | EIA | 2,396 | 28.4 |
| Kamali, 2002(66) | 1994-00 | Uganda | Community | RCT | Conv | Participants in trial Arm B - STI awareness | EIA | 2,417 | 27.9 |
| Kamali, 2002(66) | 1994-00 | Uganda | Community | RCT | Conv | Participants in trial Arm C - routine health services | EIA | 2,262 | 28.1 |
| Kapiga, 2006(67) | 2002-03 | Tanzania | Community | CS | CRS | 20-44 years old women | EIA | 1,418 | 43.8 |
| Kapiga, 2006(67) | 2002-03 | Tanzania | Community | CS | CRS | 20-44 years old men | EIA | 566 | 39.1 |
| Kasubi, 2006(68) | - | Tanzania | Outpatient clinic | CS | Conv | <15 years old children | WB | 565 | 27.3 |
| Kebede, 2004(14) | 1997-02 | Ethiopia | Community | Cohort | Conv | >19 years old adults | ELISA | 1,612 | 40.9 |
| Kuteesa, 2020(69) | 2017-18 | Uganda | Community | CS | RS | 15-24 years old adults | ELISA | 1,270 | 32.0 |
| Mehta, 2018(70) | 2014-16 | Kenya | Community | Cohort | Conv | Healthy women | ELISA | 252 | 56.8 |
| Mehta, 2018(70) | 2014-16 | Kenya | Community | Cohort | Conv | Healthy men | ELISA | 252 | 46.6 |
| MOH Uganda, 2006(71) | 2004-05 | Uganda | Community | CS | CRS | General population | ELISA | 17,953 | 46.1 |
| Msuya, 2002(72) | 1999 | Tanzania | Outpatient clinic | CS | RS | Women attending antenatal clinics | ELISA | 382 | 39.0 |
| Msuya, 2007(73) | 1999 | Tanzania | Outpatient clinic | CS | Conv | 1999 survey of pregnant women | ELISA | 382 | 39.0 |
| Msuya, 2009(74) | 2002-04 | Tanzania | Outpatient clinic | CS | Conv | 2002-04 survey of pregnant women | ELISA | 1,271 | 33.6 |
| Nakku-Joloba, 2014(75) | 2004 | Uganda | Community | CS | RS | Healthy adults | ELISA | 1,124 | 58.0 |
| Nakubulwa, 2015(76) | 2013 | Uganda | Hospital | CC | Conv | Pregnant women with PROM | ELISA | 87 | 56.0 |
| Nakubulwa, 2015(76) | 2013 | Uganda | Hospital | CC | Conv | Pregnant women in labor | ELISA | 87 | 53.0 |
| Nakubulwa, 2016(18) | 2013-14 | Uganda | Hospital | Cohort | Conv | Pregnant women | ELISA | 524 | 52.9 |
| NASCOP, 2009(77) | 2007 | Kenya | Community | CS | CRS | Kenyan population | ELISA | 15,707 | 35.1 |
| Nilsen, 2005(78) | - | Tanzania | Outpatient clinic | CS | Conv | Pregnant women | ELISA | 98 | 34.3 |
| Nilsen, 2005(78) | - | Tanzania | Outpatient clinic | CS | Conv | Blood donors | ELISA | 81 | 34.6 |
| Norris, 2009(79) | 2004 | Tanzania | Community | CS | RS | Male plantation residents | ELISA | 232 | 45.0 |
| Norris, 2009(79) | 2004 | Tanzania | Community | CS | RS | Female plantation residents | ELISA | 218 | 68.0 |
| Nyiro, 2011(80) | 2004 | Kenya | Community | CS | RS | Women participating in a DSS | ELISA | 563 | 32.0 |
| Nyiro, 2011(80) | 2004 | Kenya | Outpatient clinic | CS | Conv | Women attending a VCT | ELISA | 263 | 44.0 |
| Oliver, 2018(81) | 2014 | Kenya | Community | RCT | Conv | Women using contraceptives | ELISA | 457 | 55.6 |
| Otieno, 2015(82) | 2007-09 | Kenya | Community | Cohort | Conv | >18 years old men | ELISA | 422 | 13.3 |
| Otieno, 2015(82) | 2007-09 | Kenya | Community | Cohort | Conv | >18 years old women | ELISA | 424 | 44.8 |
| Reynolds, 2012(83) | 2007-08 | Uganda | Outpatient clinic | RCT | Conv | >18 years adults in an RCT | ELISA | 1,404 | 88.0 |
| Sivapalasingam, 2014(84) | 2010-11 | Kenya | Outpatient clinic | CC | Conv | Women reporting intravaginal practices | ELISA | 58 | 48.0 |
| Sivapalasingam, 2014(84) | 2010-11 | Kenya | Outpatient clinic | CC | Conv | Women reporting no intravaginal practices | ELISA | 42 | 43.0 |
| Tedla, 2011(85) | 2002-09 | Ethiopia | Community | CC | Conv | Patients with schizophrenia | ELISA | 216 | 7.9 |
| Tedla, 2011(85) | 2002-09 | Ethiopia | Community | CC | Conv | Patients with bipolar disorder | ELISA | 199 | 15.4 |
| Tedla, 2011(85) | 2002-09 | Ethiopia | Community | CC | RS | Healthy controls | ELISA | 80 | 10.0 |
| Tobian, 2009(24) | 2002-06 | Uganda | Community | RCT | RS | 15-49 years old men | ELISA | 6,396 | 33.8 |
| Tobian, 2012(25) | 2002-07 | Uganda | Community | RCT | Conv | Spouses of circumcised men | ELISA/WB | 835 | 55.9 |
| Tobian, 2012(25) | 2002-07 | Uganda | Community | RCT | Conv | Spouses of uncircumcised men | ELISA/WB | 803 | 53.7 |
| Todd, 2006(86) | 1994-95 | Tanzania | Community | CC | RS | Healthy women | ELISA | 430 | 63.5 |
| Todd, 2006(86) | 1994-95 | Tanzania | Community | CC | RS | Healthy men | ELISA | 420 | 46.0 |
| Wagner, 1994(27) | - | Uganda | Community | CS | Conv | >15 years old residents | WB | 212 | 67.9 |
| Wagner, 1994(27) | - | Uganda | Community | CS | Conv | <15 years old children | WB | 45 | 2.2 |
| Weiss, 2001(87) | 1997-98 | Kenya | Community | CS | CRS | Women from Kisumu | ELISA | 824 | 68.0 |
| Weiss, 2001(87) | 1997-98 | Kenya | Community | CS | CRS | Men from Kisumu | ELISA | 583 | 35.0 |
| Winston, 2015(88) | 2011-12 | Kenya | Community | CS | Conv | <21 years old street youth | ELISA | 175 | 19.0 |
| Yahya-Malima, 2008(89) | 2003-04 | Tanzania | Outpatient clinic | CS | Conv | Pregnant women | ELISA | 1,296 | 20.7 |
| Yegorov, 2018(90) | 2015-16 | Uganda | Outpatient clinic | CS | Conv | 18-45 years old women | ELISA | 58 | 58.6 |
| **Intermediate-risk populations** |  |  |  |  |  |  |  |  |  |
| Ghebrekidan, 1999(57) | 1995 | Eritrea | Community | CS | Conv | Guerrilla fighters | WB | 73 | 45.0 |
| Ghebrekidan, 1999(57) | 1995 | Eritrea | Community | CS | Conv | Truck drivers | WB | 53 | 43.0 |
| Ghebrekidan, 1999(57) | 1995 | Eritrea | Community | CS | Conv | Port workers | WB | 48 | 21.0 |
| Holt, 2003(64) | 1992 | Ethiopia | Community | CS | Conv | Male Sudanese refugees | WB | 211 | 27.0 |
| Kapiga, 2003(28) | 2000 | Tanzania | Community | CS | RS | Bars and hotels workers | ELISA | 515 | 43.5 |
| Kapiga, 2003(28) | 2000 | Tanzania | Community | CS | RS | Non sexually active bar/hotel workers | ELISA | 22 | 0.0 |
| Kapiga, 2013(67) | 2008-10 | Tanzania | Community | Cohort | Conv | Women working in food and recreational facilities | ELISA | 1,376 | 67.0 |
| Ng'ayo, 2008(91) | 2005-06 | Kenya | Community | CS | CRS | Fishermen | ELISA | 250 | 63.9 |
| Rakwar, 1997(92) | 1993 | Kenya | Outpatient clinic | Cohort | Conv | Male truck drivers | WB | 130 | 49.0 |
| Riedner, 2007(93) | 2000-02 | Tanzania | Community | CS | Conv | Women working in bars, restaurants, and guesthouses | ELISA | 753 | 88.8 |
| Tassiopoulos, 2007(32) | 2002-03 | Tanzania | Outpatient clinic | Cohort | Conv | Bar and hotel workers | ELISA | 1,045 | 56.3 |
| Valley, 2007(94) | 2002-04 | Tanzania | Community | CS | Conv | Women working in food and recreational facilities | ELISA | 1,563 | 74.6 |
| Watson-Jones, 2007(95) | 2003-05 | Tanzania | Community | CS | Conv | Women working in service facilities | ELISA | 2,719 | 80.0 |
| **Higher-risk populations** |  |  |  |  |  |  |  |  |  |
| Baeten, 2007(96) | 1993 | Kenya | Outpatient clinic | Cohort | Conv | FSWs | ELISA | 1,206 | 80.6 |
| Baltzer, 2009(97) | 2004-06 | Kenya | Outpatient clinic | Cohort | Conv | FSWs | ELISA | 139 | 87.1 |
| Braunstein, 2011a(52) | 2006-07 | Rwanda | Community | CS | Conv | FSWs | ELISA | 800 | 59.8 |
| Ghebreyordan, 1999(57) | 1995 | Eritrea | Outpatient clinic | CS | Conv | FSWs | WB | 107 | 80.0 |
| Holt, 2003(64) | 1992 | Ethiopia | Community | CS | Conv | FSWs | WB | 203 | 65.0 |
| Jespersen, 2014(65) | 2010-11 | Rwanda | Community | CS | Conv | FSWs in Rwanda | ELISA | 30 | 47.0 |
| Kaul, 2007(35) | 1998-02 | Kenya | Community | RCT | Conv | FSWs | ELISA | 443 | 72.7 |
| Masese, 2015(98) | 1993-12 | Kenya | Community | Cohort | Conv | FSWs | ELISA | 1,964 | 73.4 |
| Priddy, 2011(99) | 2008 | Kenya | Community | Cohort | Conv | FSWs | ELISA | 200 | 72.0 |
| Vandenhoudt, 2013(100) | 1997 | Kenya | Community | CS | Conv | FSWs recruited in 1997 | ELISA | 286 | 93.4 |
| Vandenhoudt, 2013(100) | 2008 | Kenya | Community | CS | Conv | FSWs recruited in 2008 | ELISA | 479 | 83.8 |
| Vandepitte, 2011(101) | 2008-09 | Uganda | Community | Cohort | Conv | FSWs | ELISA | 1,026 | 80.0 |
| **HIV negative populations** |  |  |  |  |  |  |  |  |  |
| Mehta, 2008(102) | 2002-05 | Kenya | Community | RCT | Conv | HIV negative healthy men | ELISA | 2,771 | 27.6 |
| Nakubulwa, 2009(103) | 2005 | Uganda | Outpatient clinic | CC | Conv | HIV negative pregnant women | ELISA | 200 | 62.5 |
| Serwadda, 2003(104) | 1994-98 | Uganda | Community | CC | Conv | HIV negative control group | ELISA/WB | 496 | 57.9 |
| **HIV positive individuals and individuals in HIV discordant couples** |  |  |  |  |  |  |  |  |  |
| Baeten, 2004(105) | 1999-00 | Kenya | Outpatient clinic | RCT | Conv | HIV positive women enrolled in a trial | ELISA | 400 | 94.0 |
| Celum, 2014(45) | 2008-10 | Kenya and Uganda | Community | RCT | RS | Partners of HIV positive patients | ELISA/WB | 4,638 | 67.2 |
| del Mar Pujades, 2002(8) | 1991-94 | Tanzania | Community | CC | RS | HIV positive females enrolled in a large RCT | ELISA | 70 | 54.3 |
| del Mar Pujades, 2002(8) | 1991-94 | Tanzania | Community | CC | RS | HIV positive males enrolled in a large RCT | ELISA | 57 | 61.4 |
| Jespersen, 2014(65) | 2010-11 | Rwanda | Community | CS | Conv | HIV positive +women | ELISA | 30 | 83.0 |
| Madebe, 2020(106) | - | Tanzania | Hospital | CS | Conv | >10 years old HIV positive patients | ELISA | 180 | 18.0 |
| McClelland, 2002(107) | 1996-99 | Kenya | Hospital | CS | Conv | HIV positive women | ELISA | 210 | 95.2 |
| Muiru, 2013(47) | 2007-09 | Kenya | VCT | CS | Conv | Participants from HIV discordant couples | ELISA | 938 | 58.0 |
| Nakubulwa, 2009(103) | 2005 | Uganda | Outpatient clinic | CC | Conv | HIV positive pregnant women | ELISA | 50 | 86.0 |
| Roxby, 2011(108) | 1999-02 | Kenya | Outpatient clinic | Cohort | Conv | HIV positive pregnant women | ELISA | 296 | 85.8 |
| Serwadda, 2003(104) | 1994-98 | Uganda | Community | CC | Conv | HIV seroconverters | ELISA/WB | 248 | 70.2 |
| Todd, 2006(86) | 1994-95 | Tanzania | Community | Nested CC | RS | HIV positive women | ELISA | 37 | 70.3 |
| Todd, 2006(86) | 1994-95 | Tanzania | Community | Nested CC | RS | HIV positive men | ELISA | 36 | 61.1 |
| **STI clinic attendees and symptomatic populations<sup>†</sup>** |  |  |  |  |  |  |  |  |  |
| Gorander, 2006(60) | 2001 | Tanzania | Hospital | CS | Conv | Patients with GUD | WB | 198 | 78.3 |
| Langeland, 1998(109) | 1989-93 | Tanzania | Outpatient clinic | CS | Conv | STI clinic attendees | ELISA | 294 | 42.9 |
| Mostad, 2000(110) | 1994-96 | Kenya | Outpatient clinic | CS | Conv | Women attending an STI clinic | EIA | 314 | 93.3 |
| Mwansasu, 2002(111) | - | Tanzania | Community | CS | Conv | Patients with GUD | ELISA | 69 | 79.7 |
| Nilsen, 2005(78) | - | Tanzania | Outpatient clinic | CS | Conv | STI clinic attendees | ELISA | 494 | 70.4 |
| Suntoke, 2009(112) | 2002-06 | Uganda | Community | CS | Conv | Males and females with GUD | ELISA | 95 | 77.0 |
| **Other populations** |  |  |  |  |  |  |  |  |  |
| Kassa, 2019(113) | 2005-09 | Ethiopia | Outpatient clinic/Hospital | CS | Conv | HIV negative and HIV positive pregnant women | ELISA | 2,532 | 41.5 |
| Okuku, 2011(48) | 2005-08 | Kenya | Outpatient clinic | Cohort | Conv | Adults engaging in different risky sexual behaviors | ELISA | 1,272 | 32.6 |
| Van de Wijgert, 2009(26) | 1999-04 | Uganda | Outpatient clinic | Cohort | Conv | FSWs and healthy women | ELISA | 2,199 | 48.7 |

<sup>†</sup>The reported study design is the original study design (case control, cross sectional, longitudinal cohort, or randomized controlled trial). The included seroprevalence measures are those for the baseline measures at the beginning of the study.
<sup>†</sup>Symptomatic populations include patients with clinical manifestations related to an STI.
Abbreviations: CC = Case control, Conv = Convenience, CRS = Cluster random sampling, CS = Cross sectional, DSS = Demographic surveillance system, EIA = Enzyme immunoassay, ELISA = Enzyme-linked
immunosorbent assay, FSWs = Female sex workers, GUD = Genital ulcer disease, HIV = Human immunodeficiency virus, HSV-2 = Herpes simplex virus type 2, MOH = Ministry of Health, PROM = Premature rupture of membranes, RCT = Randomized controlled trial, RS = Random sampling, STI = Sexually transmitted infection, VCT = Voluntary counselling and testing, WB = Western blot.

**Table S5.** Studies reporting HSV-2 seroprevalence in Southern Africa. This table includes only overall and not stratified seroprevalence measures,

| Author, year | Year(s) of data collection | Country | Study site | Original study design* | Sampling method | Population | HSV-2 serological assay | Sample size | HSV-2 seroprevalence (%) |
| --- | --- | --- | --- | --- | --- | --- | --- | --- | --- |
| **General populations** |  |  |  |  |  |  |  |  |  |
| Abbai, 2018(114) | 2017 | South Africa | Outpatient clinic | CS | Conv | Pregnant women | ELISA | 248 | 71.0 |
| Abdool Karim, 2014(4) | 2010 | South Africa | Community | CS | Conv | >14 years old males | ELISA | 1,252 | 2.6 |
| Abdool Karim, 2014(4) | 2017 | South Africa | Community | CS | Conv | >13 years old females | ELISA | 1,423 | 10.7 |
| Achilles, 2016(115) | - | Zimbabwe | Community | CS | Conv | 18-34 years old Zimbabwean women | ELISA | 200 | 34.0 |
| Austrian, 2016(116) | 2016-16 | Zambia | Community | RCT | CRS | 15-19 years old females | ELISA | 2,360 | 7.0 |
| Baird, 2012(117) | 2007 | Malawi | Community | RCT | CRS | Schoolgirls as a control group | RDT | 796 | 3.0 |
| Baird, 2012(117) | 2007 | Malawi | Community | RCT | CRS | School dropouts as a control group | RDT | 208 | 8.0 |
| Birdthistle, 2008(118) | 2004 | Zimbabwe | Community | CS | CRS | 14-20 years old females | ELISA | 746 | 11.7 |
| Bradley, 2018(119) | 2013-15 | South Africa | Community | RCT | RS | South African women | ELISA | 12,179 | 60.0 |
| Bradley, 2018(119) | 2013-15 | South Africa | Community | RCT | RS | South African men | ELISA | 5,472 | 27.0 |
| Bradley, 2018(119) | 2013-15 | Zambia | Community | RCT | RS | Zambian women | ELISA | 14,002 | 50.0 |
| Bradley, 2018(119) | 2013-15 | Zambia | Community | RCT | RS | Zambian men | ELISA | 5,309 | 22.0 |
| Chatterjee, 2010(120) | 1998-01 | South Africa | Outpatient clinic | CC | Conv | Women without cervical cancer | ELISA | 407 | 65.1 |
| Cowan, 2008b(121) | 2003 | Zimbabwe | Community | RCT | RS | Healthy students | ELISA | 6,791 | 0.2 |
| Crucitti, 2011(122) | - | Zambia | Community | CS | Conv | 13-16 years old females | ELISA | 450 | 5.6 |
| De Baetselier, 2015(7) | 2009-10 | South Africa | Outpatient clinic | Cohort | Conv | Healthy women | ELISA | 701 | 41.1 |
| Delany-Moretlwe, 2010(123) | 2003 | South Africa | Outpatient clinic | CS | Conv | Women at a family planning clinic | ELISA | 210 | 65.7 |
| Fearon, 2017(124) | 2011-12 | South Africa | Community | RCT | Conv | 13-20 years old students | ELISA | 2,326 | 4.6 |
| Ferrand, 2010a(125) | 2009 | Zimbabwe | Outpatient clinic | CS | Conv | Primary health care attendees | ELISA | 506 | 4.2 |
| Ferrand, 2010a(125) | 2009 | Zimbabwe | Outpatient clinic | CS | Conv | ANC attendees | ELISA | 88 | 14.0 |
| Ferrand, 2010b(126) | 2007-08 | Zimbabwe | Hospital | CS | Conv | 10-18 years old patients | ELISA | 301 | 1.3 |
| Francis, 2018(127) | 2016-17 | South Africa | Community | CS | RS | 15-24 years old females | ELISA | 259 | 28.7 |
| Francis, 2018(127) | 2016-17 | South Africa | Community | CS | RS | 15-24 years old males | ELISA | 188 | 16.8 |
| Glynn, 2008(128) | 1988-90 | Malawi | Community | CS | Conv | Sample collected from women in 1988-90 | ELISA | 343 | 56.0 |
| Glynn, 2008(128) | 1988-90 | Malawi | Community | CS | Conv | Samples collected from men in 1988-90 | ELISA | 334 | 33.2 |
| Glynn, 2008(128) | 1998-01 | Malawi | Community | CS | Conv | Samples collected from women in 1998-01 | ELISA | 384 | 66.7 |
| Glynn, 2008(128) | 1998-01 | Malawi | Community | CS | Conv | Samples collected from men in 1998-01 | ELISA | 335 | 57.0 |
| Glynn, 2008(128) | 2002-05 | Malawi | Community | CS | Conv | Samples collected from women in 2002-05 | ELISA | 205 | 55.6 |
| Glynn, 2008(128) | 2002-05 | Malawi | Community | CS | Conv | Samples collected from men in 2002-05 | ELISA | 178 | 42.1 |
| Glynn, 2008(128) | 1999-00 | Malawi | Community | CS | Conv | Samples collected from ANC attendees in 1999-00 | ELISA | 981 | 47.6 |
| Glynn, 2014(129) | 2007 | Malawi | Community | CS | Conv | 15-30 years old women | ELISA | 3,419 | 25.5 |
| Gray, 2011(130) | 2007 | South Africa | Community | RCT | Conv | >18 years old healthy individuals | WB | 801 | 31.0 |
| Gregson, 2001(131) | 1998 | Zimbabwe | Community | CS | Conv | Healthy population | ELISA | 144 | 63.2 |
| Gwanzura, 2002(132) | 2006 | Zimbabwe | Community | CS | RS | Blood donors | IB | 299 | 9.7 |
| Hazel, 2015(133) | 2009 | Namibia | Community | CS | Conv | Mobile rural pastoralists | RDT | 402 | 35.0 |
| Heffron, 2011(10) | 2006-07 | Zambia | Community | Cohort | Conv | Male sugar cane farmers | ELISA | 1,062 | 54.4 |
| Heffron, 2011(10) | 2006-07 | Zambia | Community | Cohort | Conv | Male migrant farmers | ELISA | 498 | 62.5 |
| Heffron, 2011(10) | 2006-07 | Zambia | Community | Cohort | Conv | Male non-migrant farmers | ELISA | 564 | 47.3 |
| Jespers, 2014(65) | 2010-11 | South Africa | Outpatient clinic | CS | Conv | South African women | ELISA | 109 | 40.0 |
| Jespers, 2014(65) | 2010-11 | South Africa | Outpatient clinic | CS | Conv | South African pregnant women | ELISA | 30 | 37.0 |
| Jespers, 2014(65) | 2010-11 | South Africa | Outpatient clinic | CS | Conv | South African adolescents | ELISA | 30 | 3.0 |
| Jespers, 2014(65) | 2010-11 | South Africa | Outpatient clinic | CS | Conv | South African women engaging in vaginal practices | ELISA | 31 | 45.0 |
| Jewkes, 2008(11) | 2003-04 | South Africa | Community | RCT | CRS | Women in the intervention arm in a trial | ELISA | 715 | 27.6 |
| Jewkes, 2008(11) | 2003-04 | South Africa | Community | RCT | CRS | Women in the control arm in a trial | ELISA | 701 | 31.0 |
| Jewkes, 2008(11) | 2003-04 | South Africa | Community | RCT | CRS | Men in the intervention arm in a trial | ELISA | 694 | 10.3 |
| Jewkes, 2008(11) | 2003-04 | South Africa | Community | RCT | CRS | Men in the control arm in a trial | ELISA | 666 | 10.0 |
| Kapina, 2009(134) | 2003-04 | Zambia | Community | Cohort | Conv | Healthy women | ELISA | 239 | 38.9 |
| Kenyon, 2013(135) | 2000 | South Africa | Community | CS | RS | Students or unemployed women | ELISA | 771 | 53.3 |
| Kenyon, 2013(135) | 2000 | South Africa | Community | CS | RS | Students or unemployed men | ELISA | 718 | 17.0 |
| Kharsany, 2020(136) | 2014-15 | South Africa | Community | CS | RS | Healthy adults | ELISA | 9,786 | 57.8 |
| Kjetland, 2005(137) | 1998-99 | Zimbabwe | Community | CS | Conv | Women in rural Zimbabwe | ELISA | 476 | 64.5 |
| Kurewa, 2010(138) | 2002-03 | Zimbabwe | Outpatient clinic | CS | Conv | Pregnant women | ELISA | 678 | 51.1 |
| Luseno, 2014(139) | 2012 | Zimbabwe | Community | CS | Conv | Female adolescent orphans | ELISA | 287 | 6.0 |
| Mbizvo, 2002(140) | 1999-00 | Zimbabwe | Outpatient clinic | CS | Conv | Women presenting to polyclinics | ELISA | 389 | 42.2 |
| McFarland, 1999(15) | 1993-97 | Zimbabwe | Community | CS | Conv | >18 years old male factory workers | IB | 2,397 | 39.8 |
| Menezes, 2018(141) | 2012-13 | South Africa | Outpatient clinic | RCT | Conv | Young healthy females | ELISA | 388 | 46.0 |
| Munjoma, 2010(17) | 2002 | Zimbabwe | Outpatient clinic | CS | Conv | Husbands of pregnant women | ELISA | 43 | 46.0 |
| NIMH Collaborative, 2007(142) | 2001 | Zimbabwe | Community | CS | RS | 16-30 years old healthy women | ELISA | 891 | 58.6 |
| NIMH Collaborative, 2007(142) | 2001 | Zimbabwe | Community | CS | RS | 16-30 years old healthy men | ELISA | 683 | 26.6 |
| Pascoe, 2015(143) | 2007 | Zimbabwe | Community | CS | RS | 18-22 years old females | ELISA | 2,505 | 11.2 |
| Price, 2016(144) | 2011-12 | South Africa | Community | RCT | RS | Females going to school | ELISA | 2,533 | 5.0 |
| Sobngwi-Tambekou, 2009(22) | 2002-04 | South Africa | Community | RCT | RS | Healthy males | ELISA | 3,274 | 5.9 |
| Mensch, 2020(16) | 2007-13 | Malawi | Community | Cohort | RS | School students | ELISA | 2,392 | 13.4 |
| van de Wijgert, 2009(26) | 1999-04 | Zimbabwe | Outpatient clinic | Cohort | Conv | 18-35 years old women | ELISA | 2,240 | 53.2 |
| Wand, 2012(145) | 2002-05 | South Africa | Community | Cohort | Conv | Women in Durban | ELISA | 3,492 | 73.0 |
| Weiss, 2001(87) | 1997-98 | Zambia | Community | CS | CRS | Women from Ndola | ELISA | 885 | 55.0 |
| Weiss, 2001(87) | 1997-98 | Zambia | Community | CS | CRS | Men from Ndola | ELISA | 607 | 36.0 |
| **Intermediate-risk populations** |  |  |  |  |  |  |  |  |  |
| Meque, 2014(29) | 2009-12 | Mozambique | Community | CS | Conv | Women working in service facilities | ELISA | 409 | 60.6 |
| **Higher-risk populations** |  |  |  |  |  |  |  |  |  |
| Cowan, 2005(146) | - | Zimbabwe | Community | CS | Conv | FSWs | ELISA | 369 | 77.8 |
| Ramjee, 2005(37) | 1996-97 | South Africa | Community | RCT | Conv | FSWs | ELISA | 416 | 84.0 |
| **HIV negative populations** |  |  |  |  |  |  |  |  |  |
| Abdool Karim, 2015(147) | 2007-10 | South Africa | Community | Cohort | Conv | HIV negative women | ELISA | 888 | 51.4 |
| Balkus, 2016(148) | 2005-08 | 4 African countries<sup>†</sup> | Community | RCT | Conv | Adult women from 4 African countries | ELISA | 2,830 | 42.0 |
| Barnabas, 2018(149) | 2013-15 | South Africa | Community | Cohort | Conv | Young women from Cape Town | ELISA | 149 | 21.0 |
| Benjamin, 2008(150) | 2005 | South Africa | Community | CC | Conv | HIV negative blood donors | ELISA | 102 | 19.6 |
| Benjamin, 2008(150) | 2005 | South Africa | Community | CS | Conv | Blood donors in 2005 | ELISA | 200 | 8.5 |
| Gust, 2016(151) | 2007-10 | Botswana | Community | RCT | Conv | All male and female participants in a study | ELISA | 1,201 | 35.6 |
| Mavedzenge, 2011(152) | 2003-05 | South Africa | Community | Cohort | Conv | Women in Johannesburg | ELISA | 1,008 | 65.0 |
| Mavedzenge, 2011(152) | 2003-05 | Zimbabwe | Community | Cohort | Conv | Women in Harare | ELISA | 2,455 | 51.0 |
| Mlisana, 2012(43) | 2004-05 | South Africa | Outpatient clinic | CS | Conv | Healthy women | ELISA | 242 | 86.0 |
| Perti, 2014(44) | - | South Africa | Hospital | CS | Conv | HIV negative pregnant women | ELISA/WB | 255 | 43.5 |
| Wand, 2013(153) | 2003-06 | South Africa | Community | Cohort | Conv | Women born before 1960 | ELISA | 110 | 89.0 |
| Wand, 2013(153) | 2003-06 | South Africa | Community | Cohort | Conv | Women born between 1960-1964 | ELISA | 257 | 81.8 |
| Wand, 2013(153) | 2003-06 | South Africa | Community | Cohort | Conv | Women born between 1965-1969 | ELISA | 409 | 81.6 |
| Wand, 2013(153) | 2003-06 | South Africa | Community | Cohort | Conv | Women born between 1970-1974 | ELISA | 559 | 81.4 |
| Wand, 2013(153) | 2003-06 | South Africa | Community | Cohort | Conv | Women born between 1975-1979 | ELISA | 770 | 76.7 |
| Wand, 2013(153) | 2003-06 | South Africa | Community | Cohort | Conv | Women born between 1980-1984 | ELISA | 1,043 | 66.2 |
| Wand, 2013(153) | 2003-06 | South Africa | Community | Cohort | Conv | Women born after 1985 | ELISA | 324 | 43.3 |
| Sutcliffe, 2002(154) | 1998 | Malawi | Community | CC | Conv | HIV negative males | ELISA | 280 | 64.3 |
| **HIV positive individuals and individuals in HIV discordant couples** |  |  |  |  |  |  |  |  |  |
| Benjamin, 2008(150) | 2005 | South Africa | Community | CC | Conv | HIV positive blood donors | ELISA | 106 | 72.6 |
| Benjamin, 2008(150) | 2001-02 | South Africa | Community | CS | Conv | HIV positive blood donors in 2001-2002 | ELISA | 625 | 69.3 |
| Cowan, 2008a(46) | 1997-00 | Zimbabwe | Outpatient clinic | CC | RS | HIV positive mothers with HIV positive infants | ELISA/WB | 478 | 86.2 |
| Cowan, 2008a(46) | 1997-00 | Zimbabwe | Outpatient clinic | CC | RS | HIV positive mothers with HIV negative infants | ELISA/WB | 970 | 80.7 |
| Lewis, 2012(155) | 2007 | South Africa | Outpatient clinic | CS | Conv | HIV positive patients | ELISA | 1,106 | 85.2 |
| Lowe, 2019(156, 157) | 2016 | South Africa | Outpatient clinic | CS | Conv | HIV positive women | ELISA | 385 | 52.5 |
| Perti, 2014(44) | - | South Africa | Hospital | CS | Conv | HIV positive pregnant women | ELISA/WB | 132 | 87.9 |
| Rabenau, 2010(158) | 2007 | Lesotho | Outpatient clinic | CS | Conv | HIV positive patients | ELISA | 205 | 78.5 |
| Sutcliffe, 2002(154) | 1998 | Malawi | Community | CC | Conv | HIV positive males | ELISA | 279 | 88.1 |
| Varo, 2016(159) | 2008-10 | Malawi | Hospital | CS | RS | <14 years old HIV positive children | ELISA | 91 | 4.4 |
| **STI clinic attendees and symptomatic populations<sup>‡</sup>** |  |  |  |  |  |  |  |  |  |
| Chen, 2000(160) | 1993-94 | South Africa | Outpatient clinic | CS | Conv | Men with GUD | WB | 498 | 49.2 |
| Chen, 2000(160) | 1993-94 | South Africa | Outpatient clinic | CS | Conv | Men with urethritis | WB | 554 | 42.1 |
| Esber, 2017(161) | 2015 | Malawi | Outpatient clinic | CS | Conv | Symptomatic women attending clinic | ELISA | 197 | 50.0 |
| Hoyo, 2005(162) | 1998-99 | Malawi | Outpatient clinic | CS | Conv | STI clinic attendees with GUD | WB | 136 | 80.0 |
| Kufa, 2020(157) | 2017-19 | South Africa | Outpatient clinic | CS | Conv | Men attending an STI clinic | ELISA | 847 | 51.0 |
| Kularatne, 2018(163) | 2007-15 | South Africa | Outpatient clinic | CS | Conv | Patients attending an STI clinic | ELISA | 771 | 80.2 |
| Lewis, 2008(164) | 2006 | South Africa | Community | CS | Conv | Male STI clinic attendees | ELISA | 303 | 57.1 |
| Lewis, 2013(165) | 2007-12 | South Africa | Outpatient clinic | CS | Conv | Men with urethral discharge | ELISA | 1,204 | 55.6 |
| Lewis, 2013(165) | 2007-12 | South Africa | Outpatient clinic | CS | Conv | Women with vaginal discharge | ELISA | 1,216 | 77.7 |
| Mhlono, 2010(166) | 2006-07 | South Africa | Outpatient clinic | CS | Conv | Females in Cape Town with VDS | ELISA | 92 | 38.0 |
| Mhlono, 2010(166) | 2006-07 | South Africa | Outpatient clinic | CS | Conv | Males in Cape Town with MUS Syndrome | ELISA | 279 | 30.5 |
| Mhlono, 2010(166) | 2006-07 | South Africa | Outpatient clinic | CS | Conv | Females in Johannesburg with VDS | ELISA | 199 | 75.9 |
| Mhlono, 2010(166) | 2006-07 | South Africa | Outpatient clinic | CS | Conv | Males in Johannesburg with MUS Syndrome | ELISA | 211 | 59.7 |
| Morse, 1997(167) | 1993-94 | Lesotho | Outpatient clinic | CS | Conv | Patients with GUD | WB | 99 | 55.0 |
| O'Farrell, 2007(168) | 2004 | South Africa | Outpatient clinic | CS | Conv | Males with GUD | ELISA | 162 | 86.4 |
| O'Farrell, 2007(168) | 2004 | South Africa | Outpatient clinic | CS | Conv | Males attending an STI clinic | ELISA | 480 | 72.3 |
| Paz-Bailey, 2009(169) | 2005-06 | South Africa | Outpatient clinic | RCT | RS | Men with GUD | ELISA | 615 | 70.6 |
| Phiri, 2013(170) | 2004-06 | Malawi | Outpatient clinic | RCT | Conv | Males and females with GUD | ELISA | 417 | 72.0 |
| Zimba, 2011(171) | 2005 | Mozambique | Outpatient clinic | CS | Conv | Patients attending a clinic with US or VDS | ELISA | 346 | 85.0 |
| **Other populations** |  |  |  |  |  |  |  |  |  |
| Chattopadhyay, 2015(172) | 1998-01 | South Africa | Community | CC | Conv | Women with cervical cancer | ELISA | 429 | 68.0 |

\* The reported study design is the original study design (case control, cross sectional, longitudinal cohort, or randomized controlled trial). The included seroprevalence measures are those for the baseline measures at the beginning of the study.
<sup>†</sup> The four African countries are: Malawi, South Africa, Zambia, and Zimbabwe.
<sup>‡</sup> Symptomatic populations include patients with clinical manifestations related to an STI.

**Table S6.**
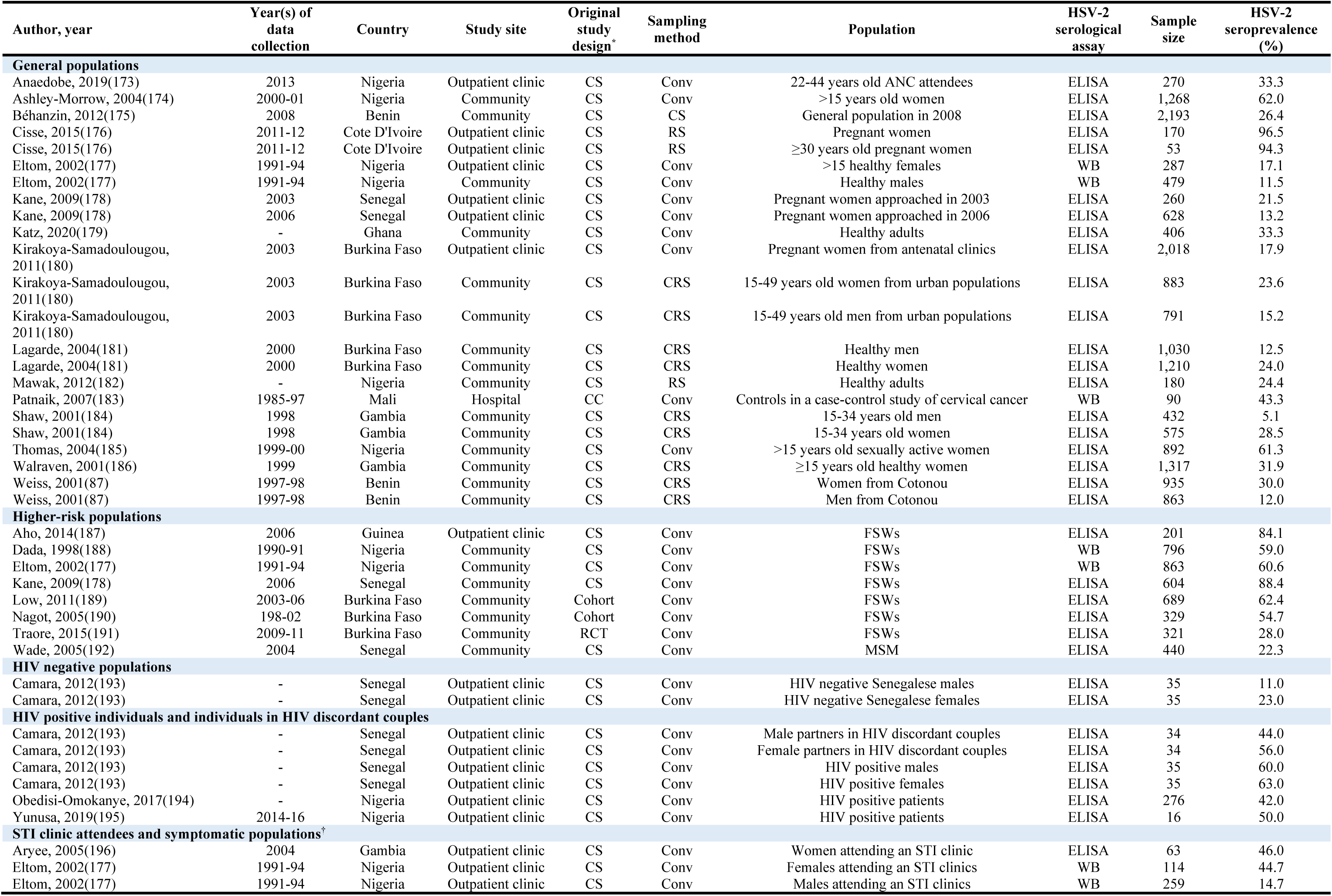
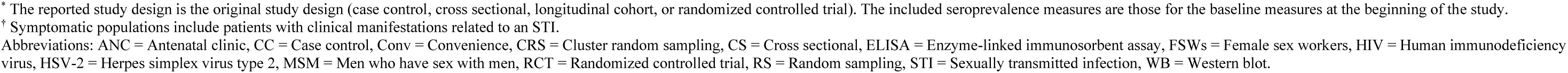
Studies reporting HSV-2 seroprevalence in Western Africa. This table includes only overall and not stratified seroprevalence measures.

| Author, year | Year(s) of data collection | Country | Study site | Original study design* | Sampling method | Population | HSV-2 serological assay | Sample size | HSV-2 seroprevalence (%) |
| --- | --- | --- | --- | --- | --- | --- | --- | --- | --- |
| **General populations** |  |  |  |  |  |  |  |  |  |
| Anaedobe, 2019(173) | 2013 | Nigeria | Outpatient clinic | CS | Conv | 22-44 years old ANC attendees | ELISA | 270 | 33.3 |
| Ashley-Morrow, 2004(174) | 2000-01 | Nigeria | Community | CS | Conv | >15 years old women | ELISA | 1,268 | 62.0 |
| Béhanzin, 2012(175) | 2008 | Benin | Community | CS | CS | General population in 2008 | ELISA | 2,193 | 26.4 |
| Cisse, 2015(176) | 2011-12 | Cote D'Ivoire | Outpatient clinic | CS | RS | Pregnant women | ELISA | 170 | 96.5 |
| Cisse, 2015(176) | 2011-12 | Cote D'Ivoire | Outpatient clinic | CS | RS | ≥30 years old pregnant women | ELISA | 53 | 94.3 |
| Eltom, 2002(177) | 1991-94 | Nigeria | Outpatient clinic | CS | Conv | >15 healthy females | WB | 287 | 17.1 |
| Eltom, 2002(177) | 1991-94 | Nigeria | Community | CS | Conv | Healthy males | WB | 479 | 11.5 |
| Kane, 2009(178) | 2003 | Senegal | Outpatient clinic | CS | Conv | Pregnant women approached in 2003 | ELISA | 260 | 21.5 |
| Kane, 2009(178) | 2006 | Senegal | Outpatient clinic | CS | Conv | Pregnant women approached in 2006 | ELISA | 628 | 13.2 |
| Katz, 2020(179) | - | Ghana | Community | CS | Conv | Healthy adults | ELISA | 406 | 33.3 |
| Kirakoya-Samadoulougou, 2011(180) | 2003 | Burkina Faso | Outpatient clinic | CS | Conv | Pregnant women from antenatal clinics | ELISA | 2,018 | 17.9 |
| Kirakoya-Samadoulougou, 2011(180) | 2003 | Burkina Faso | Community | CS | CRS | 15-49 years old women from urban populations | ELISA | 883 | 23.6 |
| Kirakoya-Samadoulougou, 2011(180) | 2003 | Burkina Faso | Community | CS | CRS | 15-49 years old men from urban populations | ELISA | 791 | 15.2 |
| Lagarde, 2004(181) | 2000 | Burkina Faso | Community | CS | CRS | Healthy men | ELISA | 1,030 | 12.5 |
| Lagarde, 2004(181) | 2000 | Burkina Faso | Community | CS | CRS | Healthy women | ELISA | 1,210 | 24.0 |
| Mawak, 2012(182) | - | Nigeria | Community | CS | RS | Healthy adults | ELISA | 180 | 24.4 |
| Patnaik, 2007(183) | 1985-97 | Mali | Hospital | CC | Conv | Controls in a case-control study of cervical cancer | WB | 90 | 43.3 |
| Shaw, 2001(184) | 1998 | Gambia | Community | CS | CRS | 15-34 years old men | ELISA | 432 | 5.1 |
| Shaw, 2001(184) | 1998 | Gambia | Community | CS | CRS | 15-34 years old women | ELISA | 575 | 28.5 |
| Thomas, 2004(185) | 1999-00 | Nigeria | Community | CS | Conv | >15 years old sexually active women | ELISA | 892 | 61.3 |
| Walraven, 2001(186) | 1999 | Gambia | Community | CS | CRS | ≥15 years old healthy women | ELISA | 1,317 | 31.9 |
| Weiss, 2001(87) | 1997-98 | Benin | Community | CS | CRS | Women from Cotonou | ELISA | 935 | 30.0 |
| Weiss, 2001(87) | 1997-98 | Benin | Community | CS | CRS | Men from Cotonou | ELISA | 863 | 12.0 |
| **Higher-risk populations** |  |  |  |  |  |  |  |  |  |
| Aho, 2014(187) | 2006 | Guinea | Outpatient clinic | CS | Conv | FSWs | ELISA | 201 | 84.1 |
| Dada, 1998(188) | 1990-91 | Nigeria | Community | CS | Conv | FSWs | WB | 796 | 59.0 |
| Eltom, 2002(177) | 1991-94 | Nigeria | Community | CS | Conv | FSWs | WB | 863 | 60.6 |
| Kane, 2009(178) | 2006 | Senegal | Community | CS | Conv | FSWs | ELISA | 604 | 88.4 |
| Low, 2011(189) | 2003-06 | Burkina Faso | Community | Cohort | Conv | FSWs | ELISA | 689 | 62.4 |
| Nagot, 2005(190) | 198-02 | Burkina Faso | Community | Cohort | Conv | FSWs | ELISA | 329 | 54.7 |
| Traore, 2015(191) | 2009-11 | Burkina Faso | Community | RCT | Conv | FSWs | ELISA | 321 | 28.0 |
| Wade, 2005(192) | 2004 | Senegal | Community | CS | Conv | MSM | ELISA | 440 | 22.3 |
| **HIV negative populations** |  |  |  |  |  |  |  |  |  |
| Camara, 2012(193) | - | Senegal | Outpatient clinic | CS | Conv | HIV negative Senegalese males | ELISA | 35 | 11.0 |
| Camara, 2012(193) | - | Senegal | Outpatient clinic | CS | Conv | HIV negative Senegalese females | ELISA | 35 | 23.0 |
| **HIV positive individuals and individuals in HIV discordant couples** |  |  |  |  |  |  |  |  |  |
| Camara, 2012(193) | - | Senegal | Outpatient clinic | CS | Conv | Male partners in HIV discordant couples | ELISA | 34 | 44.0 |
| Camara, 2012(193) | - | Senegal | Outpatient clinic | CS | Conv | Female partners in HIV discordant couples | ELISA | 34 | 56.0 |
| Camara, 2012(193) | - | Senegal | Outpatient clinic | CS | Conv | HIV positive males | ELISA | 35 | 60.0 |
| Camara, 2012(193) | - | Senegal | Outpatient clinic | CS | Conv | HIV positive females | ELISA | 35 | 63.0 |
| Obedisi-Omokanye, 2017(194) | - | Nigeria | Outpatient clinic | CS | Conv | HIV positive patients | ELISA | 276 | 42.0 |
| Yunusa, 2019(195) | 2014-16 | Nigeria | Outpatient clinic | CS | Conv | HIV positive patients | ELISA | 16 | 50.0 |
| **STI clinic attendees and symptomatic populations<sup>†</sup>** |  |  |  |  |  |  |  |  |  |
| Aryee, 2005(196) | 2004 | Gambia | Outpatient clinic | CS | Conv | Women attending an STI clinic | ELISA | 63 | 46.0 |
| Eltom, 2002(177) | 1991-94 | Nigeria | Outpatient clinic | CS | Conv | Females attending an STI clinics | WB | 114 | 44.7 |
| Eltom, 2002(177) | 1991-94 | Nigeria | Outpatient clinic | CS | Conv | Males attending an STI clinics | WB | 259 | 14.7 |

\* The reported study design is the original study design (case control, cross sectional, longitudinal cohort, or randomized controlled trial). The included seroprevalence measures are those for the baseline measures at the beginning of the study.
† Symptomatic populations include patients with clinical manifestations related to an STI.
Abbreviations: ANC = Antenatal clinic, CC = Case control, Conv = Convenience, CRS = Cluster random sampling, CS = Cross sectional, ELISA = Enzyme-linked immunosorbent assay, FSWs = Female sex workers, HIV = Human immunodeficiency virus, HSV-2 = Herpes simplex virus type 2, MSM = Men who have sex with men, RCT = Randomized controlled trial, RS = Random sampling, STI = Sexually transmitted infection, WB = Western blot.

**Table S7.** Studies reporting HSV-2 seroprevalence in Central Africa. This table includes only overall and not stratified seroprevalence measures.

| Author, year | Year(s) of data collection | Country | Study site | Original study design* | Sampling method | Population | HSV-2 serological assay | Sample size | HSV-2 seroprevalence (%) |
| --- | --- | --- | --- | --- | --- | --- | --- | --- | --- |
| **General populations** |  |  |  |  |  |  |  |  |  |
| Charpentier, 2011(197) | 2007 | Chad | Community | CS | CRS | General adult population | ELISA | 548 | 15.7 |
| Eis-Hubinger, 2002(198) | 1998 | Cameroon | Outpatient clinic | CS | RS | Patients attending a pulmonary clinic | WB | 188 | 72.3 |
| Ozouaki, 2006(199) | - | Gabon | Outpatient clinic | CS | Conv | Women of childbearing age | ELISA | 355 | 65.9 |
| Volpi, 2004(200) | 1997-98 | Cameroon | Outpatient clinic | CS | Conv | Males and females attending general clinic | ELISA | 238 | 34.0 |
| Weiss, 2001(87) | 1997-98 | Cameroon | Community | CS | CRS | Women from Yaounde | ELISA | 1,002 | 51.0 |
| Weiss, 2001(87) | 1997-98 | Cameroon | Community | CS | CRS | Men from Yaounde | ELISA | 887 | 27.0 |
| **Higher-risk populations** |  |  |  |  |  |  |  |  |  |
| Longo, 2017(201) | 2013 | Central African Republic | Community | CS | Conv | FSWs | ELISA | 345 | 11.9 |
| Nzila, 1991(202) | 1988 | Democratic Republic of Congo | Community | CS | Conv | FSWs | WB | 1,233 | 21.5 |
| **HIV negative populations** |  |  |  |  |  |  |  |  |  |
| LeGoff, 2011(203) | 2009 | Central African Republic | Hospital | CS | Conv | ≤17 years old HIV negative children | ELISA | 200 | 6.0 |
| **STI clinic attendees and symptomatic populations†** |  |  |  |  |  |  |  |  |  |
| Eis-Hubinger, 2002(198) | 1998 | Cameroon | Outpatient clinic | CS | RS | Patients attending an STI clinic | EIA | 161 | 67.1 |
| Mbopi-keou, 2000(204) | - | Central African Republic | Outpatient clinic | CS | Conv | Women attending an STI clinic | ELISA | 300 | 82.0 |
| Vandepitte, 2007(205) | 2002 | Democratic Republic of Congo | Outpatient clinic | CS | Conv | FSWs attending an STI clinic | ELISA | 501 | 58.5 |
| **Other populations** |  |  |  |  |  |  |  |  |  |
| Eis-Hubinger, 2002(198) | 1998 | Cameroon | Hospital | CS | RS | Patients with symptoms suggestive of HIV | EIA | 61 | 70.5 |

\*The reported study design is the original study design (case control, cross sectional, longitudinal cohort, or randomized controlled trial). The included seroprevalence measures are those for the baseline measures at the beginning of the study.
† Symptomatic populations include patients with clinical manifestations related to an STI.
Abbreviations: Conv = Convenience, CRS = Cluster random sampling, CS = Cross sectional, EIA = Enzyme immunoassay, ELISA = Enzyme-linked immunosorbent assay, FSWs = Female sex workers, HIV = Human immunodeficiency virus, HSV-2 = Herpes simplex virus type 2, RS = Random sampling, STI = Sexually transmitted infection, WB = Western blot.

**Table S8.** Studies reporting HSV-2 seroprevalence across several regions in sub-Saharan Africa. This table includes only overall and not stratified seroprevalence measures.

| Author, year | Year(s) of data collection | Country | Study site | Original study design* | Sampling method | Population | HSV-2 serological assay | Sample size | HSV-2 seroprevalence (%) |
| --- | --- | --- | --- | --- | --- | --- | --- | --- | --- |
| **HIV negative populations** |  |  |  |  |  |  |  |  |  |
| ECHO trial consortium(206) | 2015-17 | 4 African countries<sup>†</sup> | Community | RCT | Conv | Females using injectable contraceptives | ELISA | 2,588 | 38.7 |
| ECHO trial consortium(206) | 2015-17 | 4 African countries<sup>†</sup> | Community | RCT | Conv | Females using intrauterine contraceptives | ELISA | 2,587 | 39.4 |
| ECHO trial consortium(206) | 2015-17 | 4 African countries<sup>†</sup> | Community | RCT | Conv | Females using implantable contraceptives | ELISA | 2,584 | 37.4 |
| Marrazzo, 2015(207) | 2009-11 | 3 African countries<sup>‡</sup> | Community | RCT | Conv | Female trial participants | ELISA | 5,005 | 46.0 |
| McCormack, 2010(40) | 2005-09 | 4 African countries<sup>§</sup> | Outpatient clinic/Community | RCT | Conv | Female trial participants receiving 2% gel | ELISA | 2,725 | 60.0 |
| McCormack, 2010(40) | 2005-09 | 4 African countries<sup>§</sup> | Outpatient clinic/Community | RCT | Conv | Female trial participants receiving 0.5% gel | ELISA | 3,312 | 61.0 |
| McCormack, 2010(40) | 2005-09 | 4 African countries<sup>§</sup> | Outpatient clinic/Community | RCT | Conv | Female trial participants receiving placebo | ELISA | 3,311 | 60.0 |
| **HIV positive individuals and individuals in HIV discordant couples** |  |  |  |  |  |  |  |  |  |
| Heffron, 2012(208) | 2004-10 | 7 African countries<sup>¶</sup> | Community | Cohort | Conv | Males with an HIV positive partner | WB | 2,393 | 60.2 |
| Heffron, 2012(208) | 2004-10 | 7 African countries<sup>¶</sup> | Community | Cohort | Conv | Females with an HIV positive partner | WB | 1,283 | 84.8 |
| **STI clinic attendees and symptomatic populations<sup>‡</sup>** |  |  |  |  |  |  |  |  |  |
| LeGoff, 2007(209) | 2003-05 | 2 African countries<sup>**</sup> | Outpatient clinic | RCT | Conv | Women with GUD | ELISA | 437 | 79.0 |

\* The reported study design is the original study design (case control, cross sectional, longitudinal cohort, or randomized controlled trial). The included seroprevalence measures are those for the baseline measures at the beginning of the study.
<sup>†</sup> The four African countries are Eswatini, South Africa, Kenya, and Zambia.
<sup>‡</sup> The three African countries are South Africa, Uganda, and Zimbabwe.
<sup>§</sup> The four African countries are South Africa, Tanzania, Uganda, and Zimbabwe.
<sup>¶</sup> The seven African countries are: Botswana, Kenya, South Africa, Tanzania, Uganda, and Zambia.
<sup>‡</sup> Symptomatic populations include patients with clinical manifestations related to a sexually transmitted infection.
<sup>\*\*</sup> The two African countries are Ghana and Central African Republic
Abbreviations: Conv = Convenience, ELISA = Enzyme-linked immunosorbent assay, GUD = Genital ulcer disease, HIV = Human immunodeficiency virus, HSV-2 = Herpes simplex virus type 2, RCT = Randomized controlled trial, WB = Western blot.

**Table S9.**
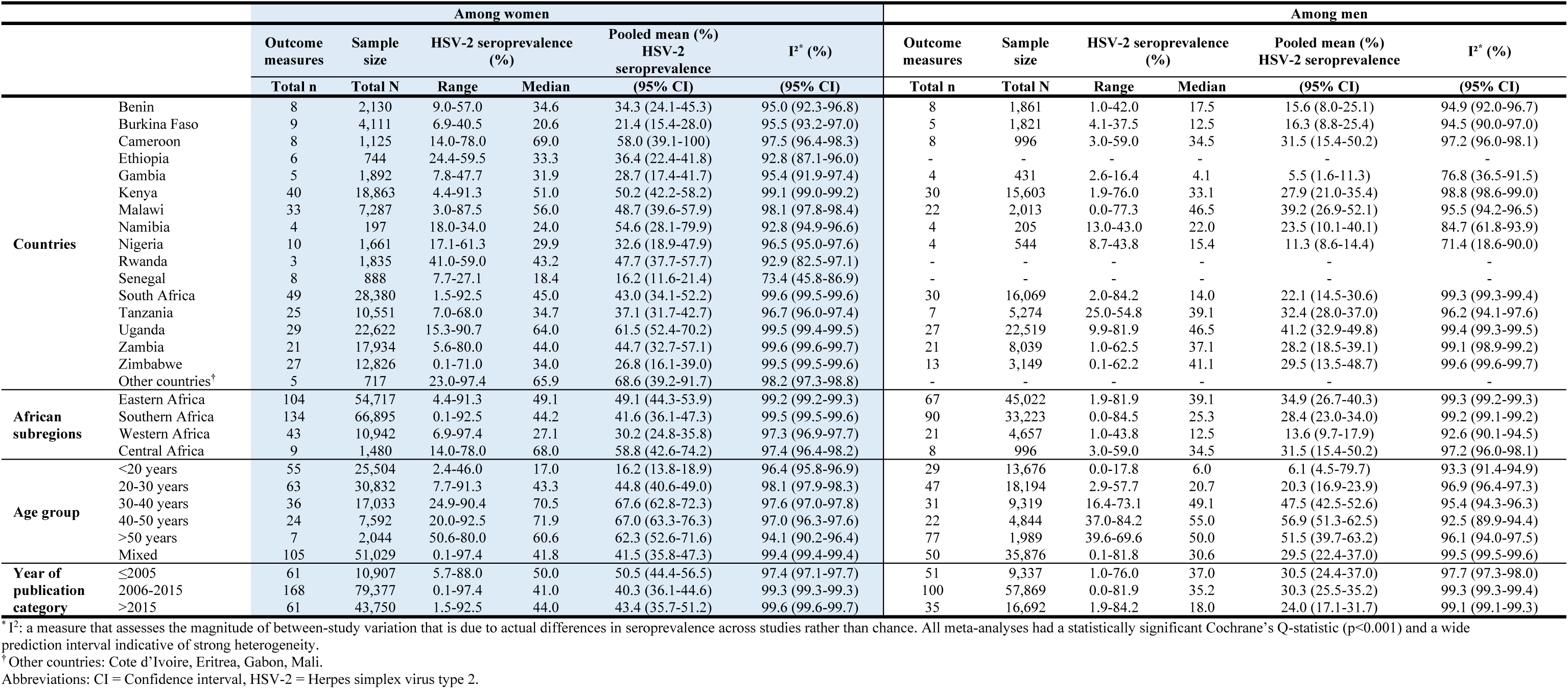
Pooled mean estimates for herpes simplex virus type 2 seroprevalence among general populations by sex stratification in sub-Saharan Africa.

**Figure S2.**
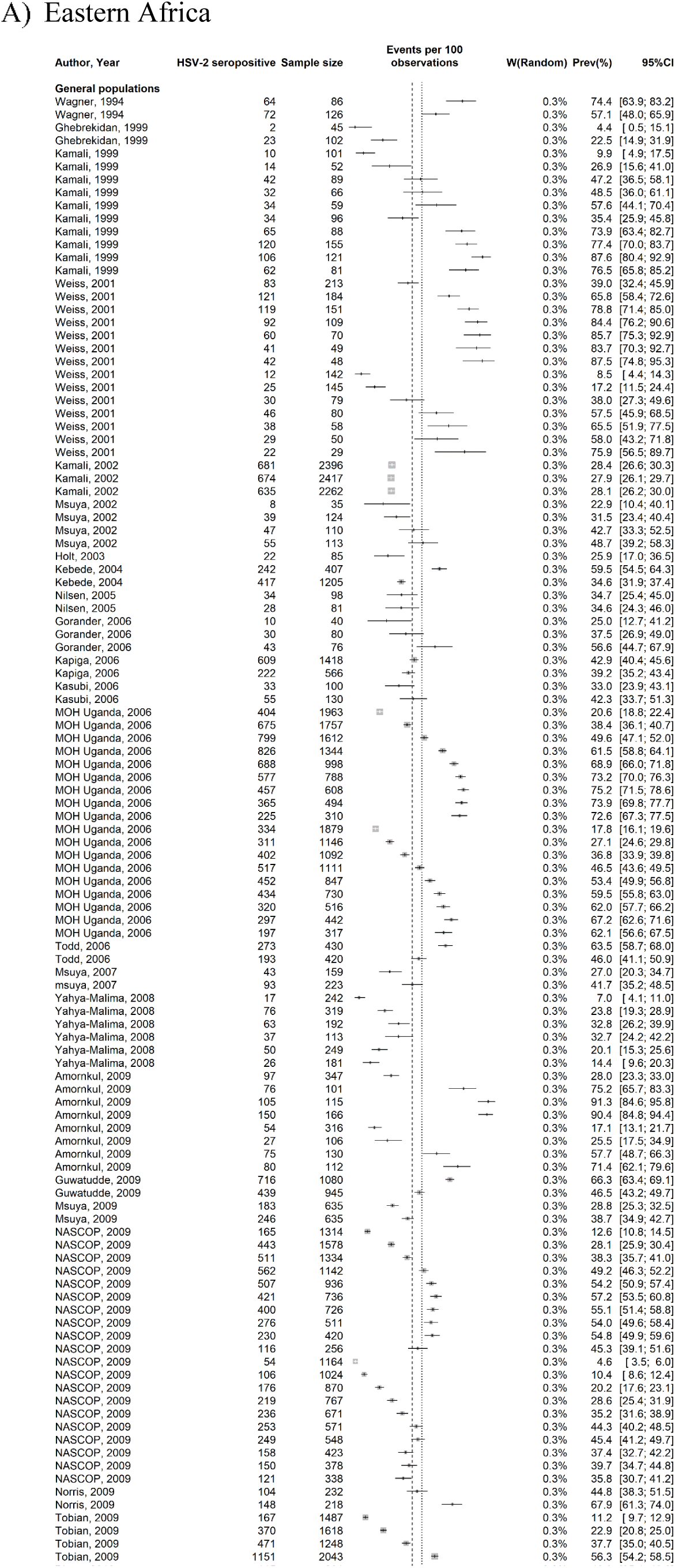
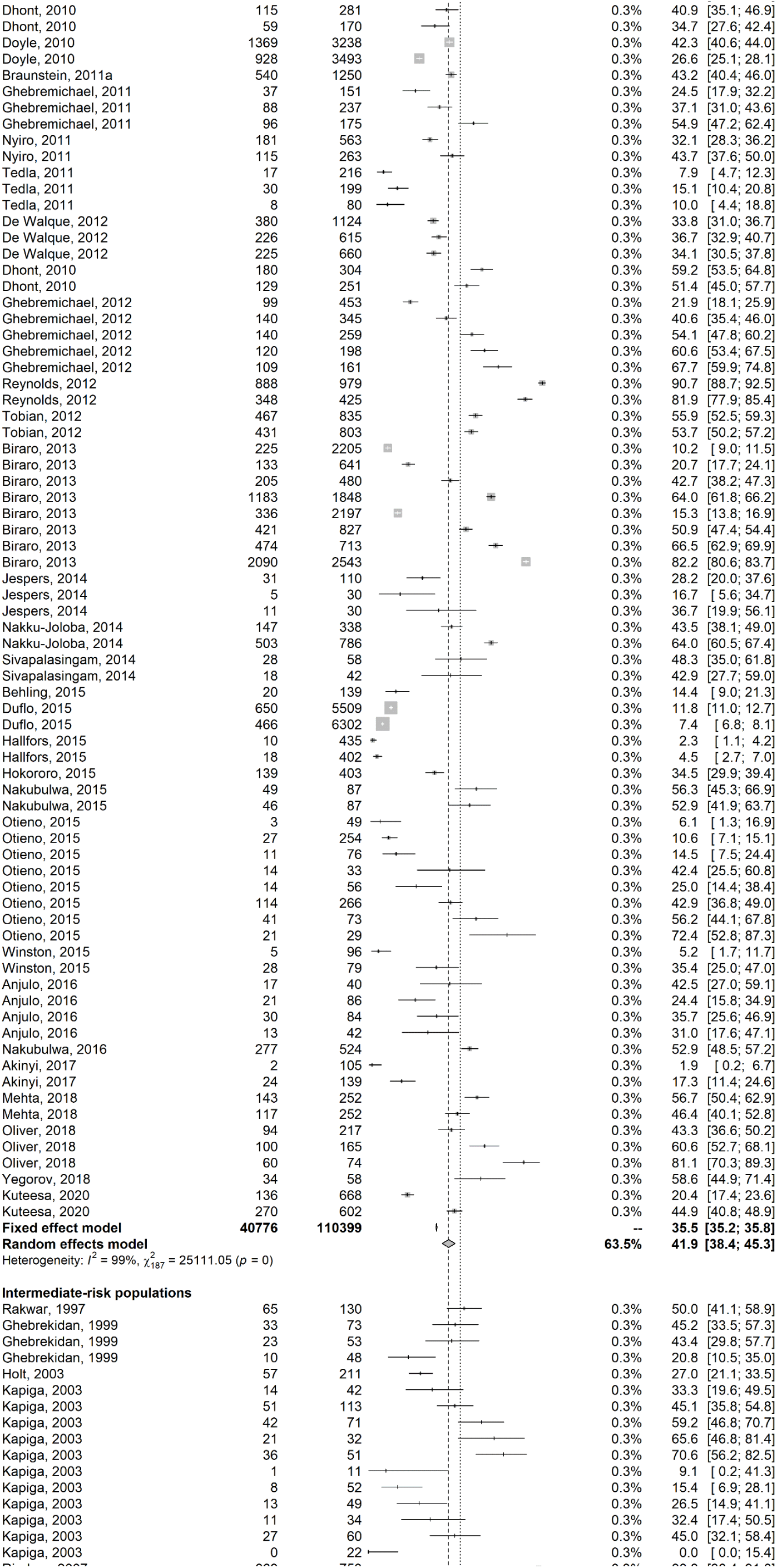
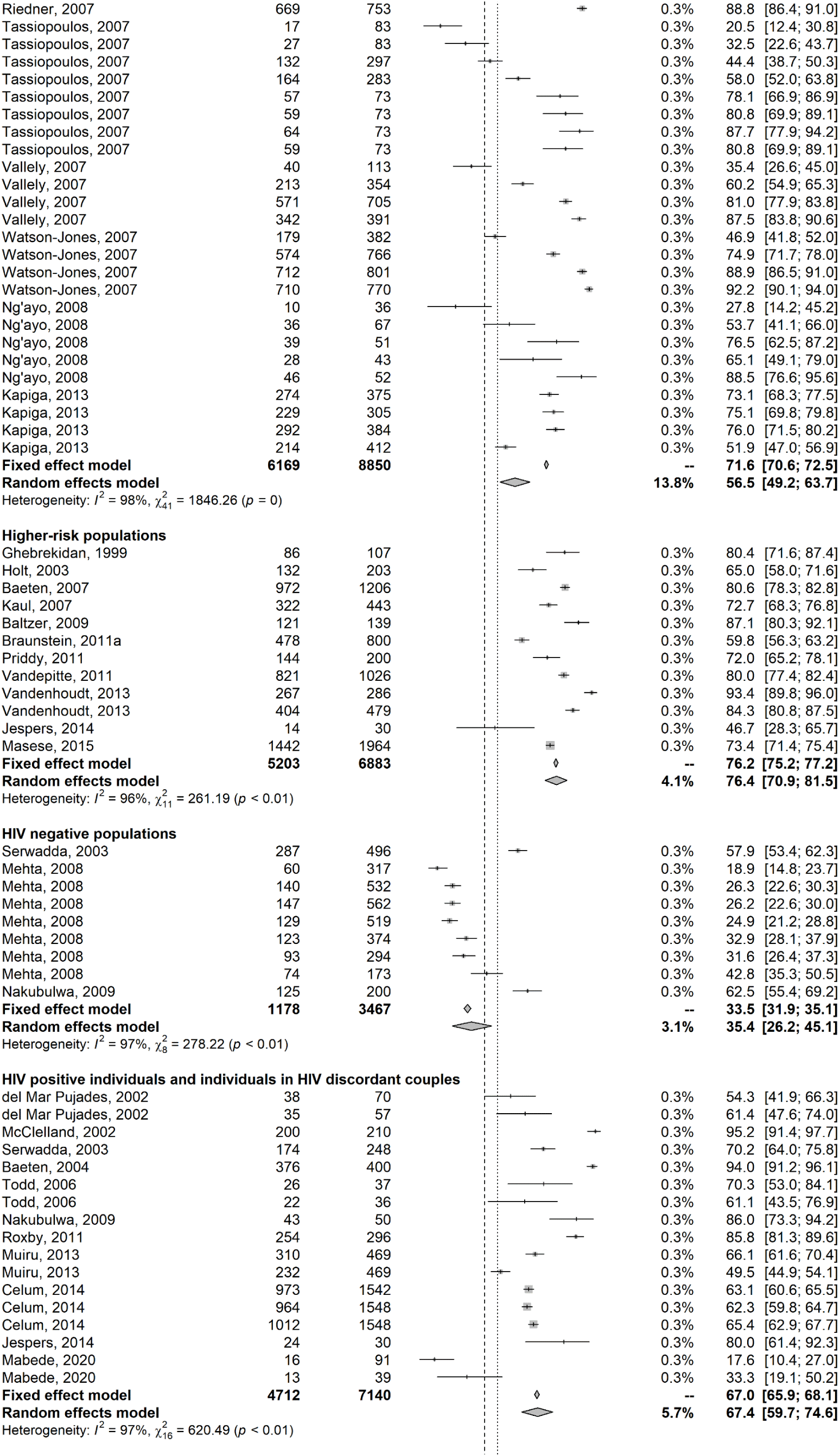
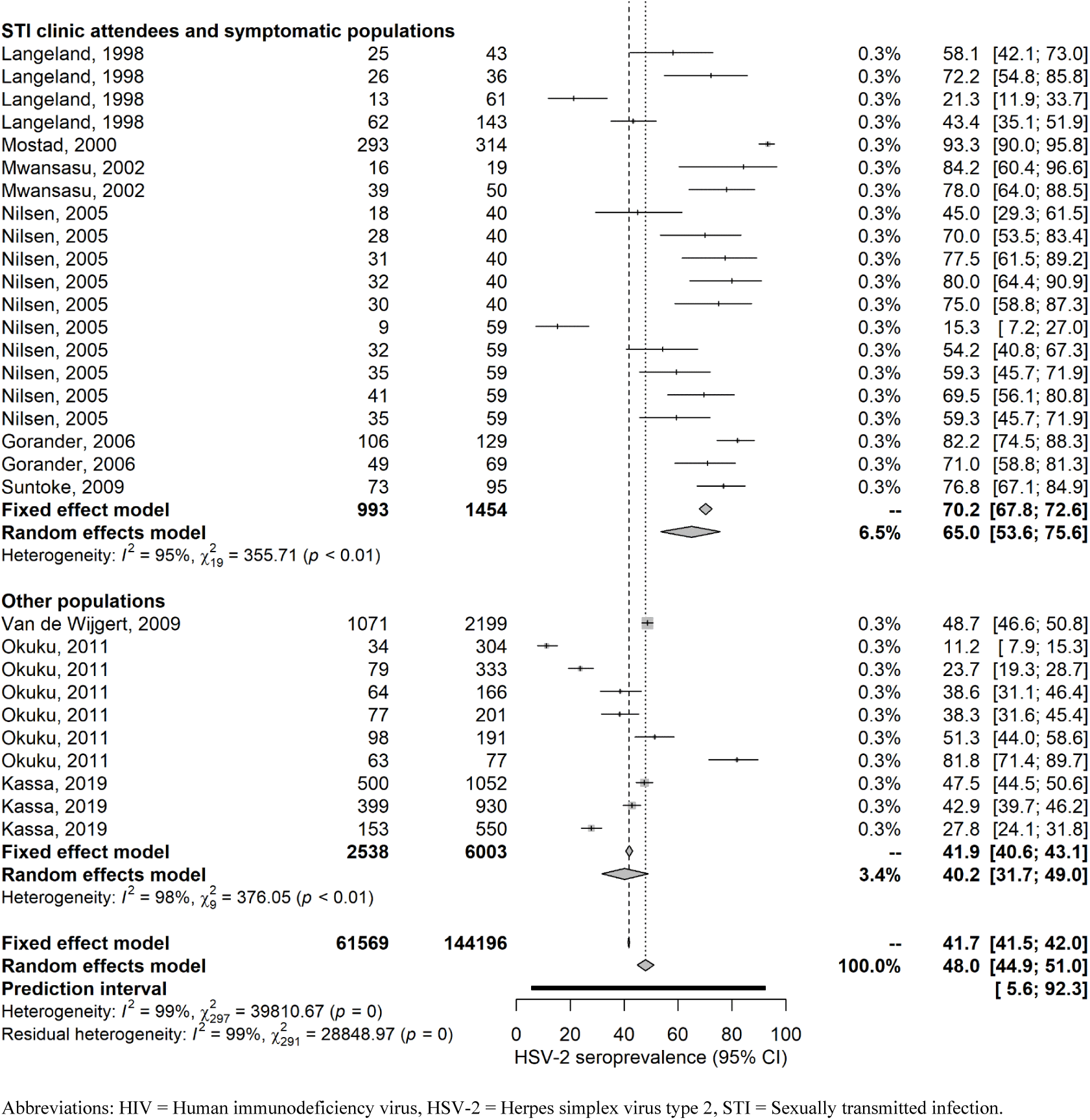
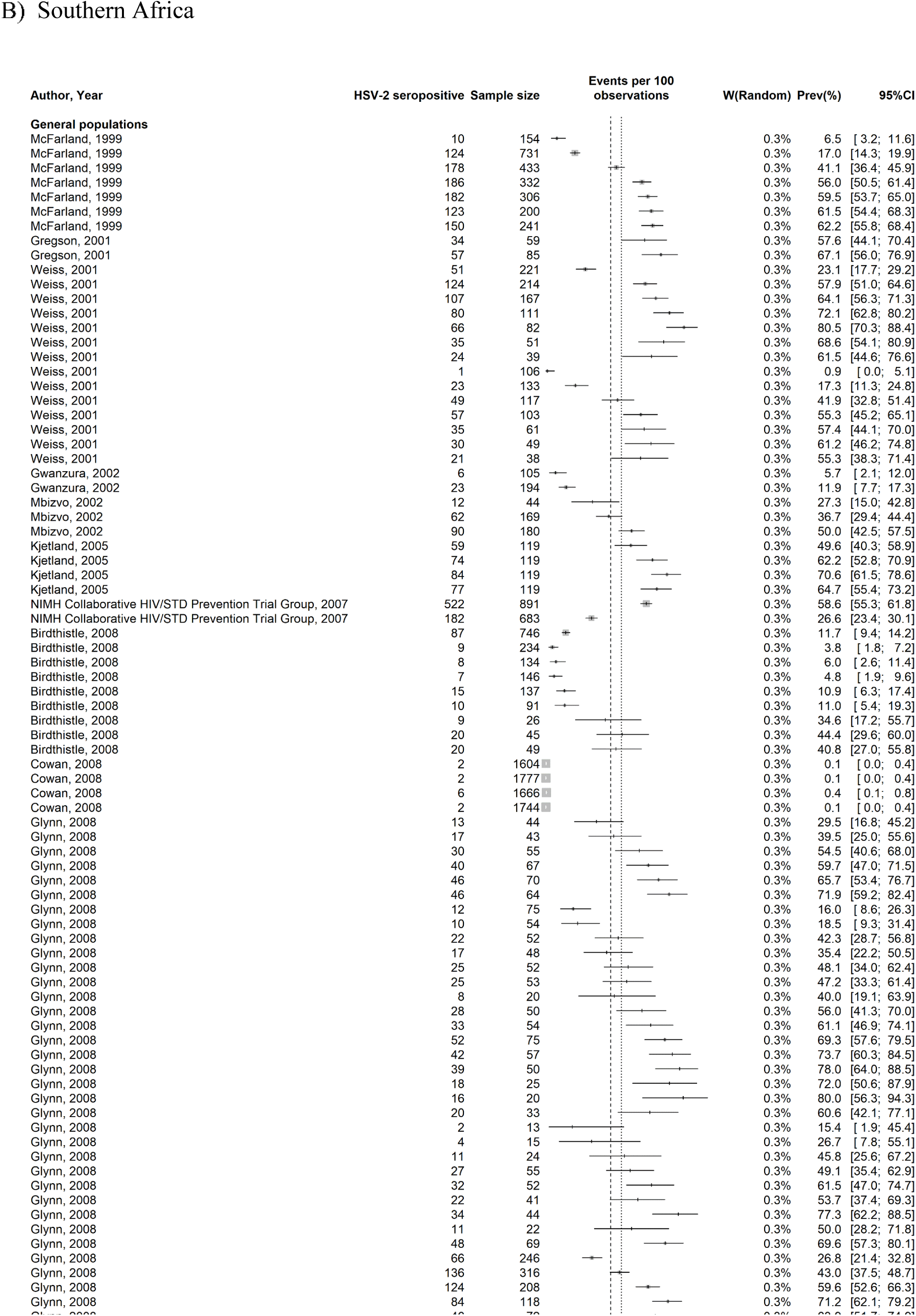
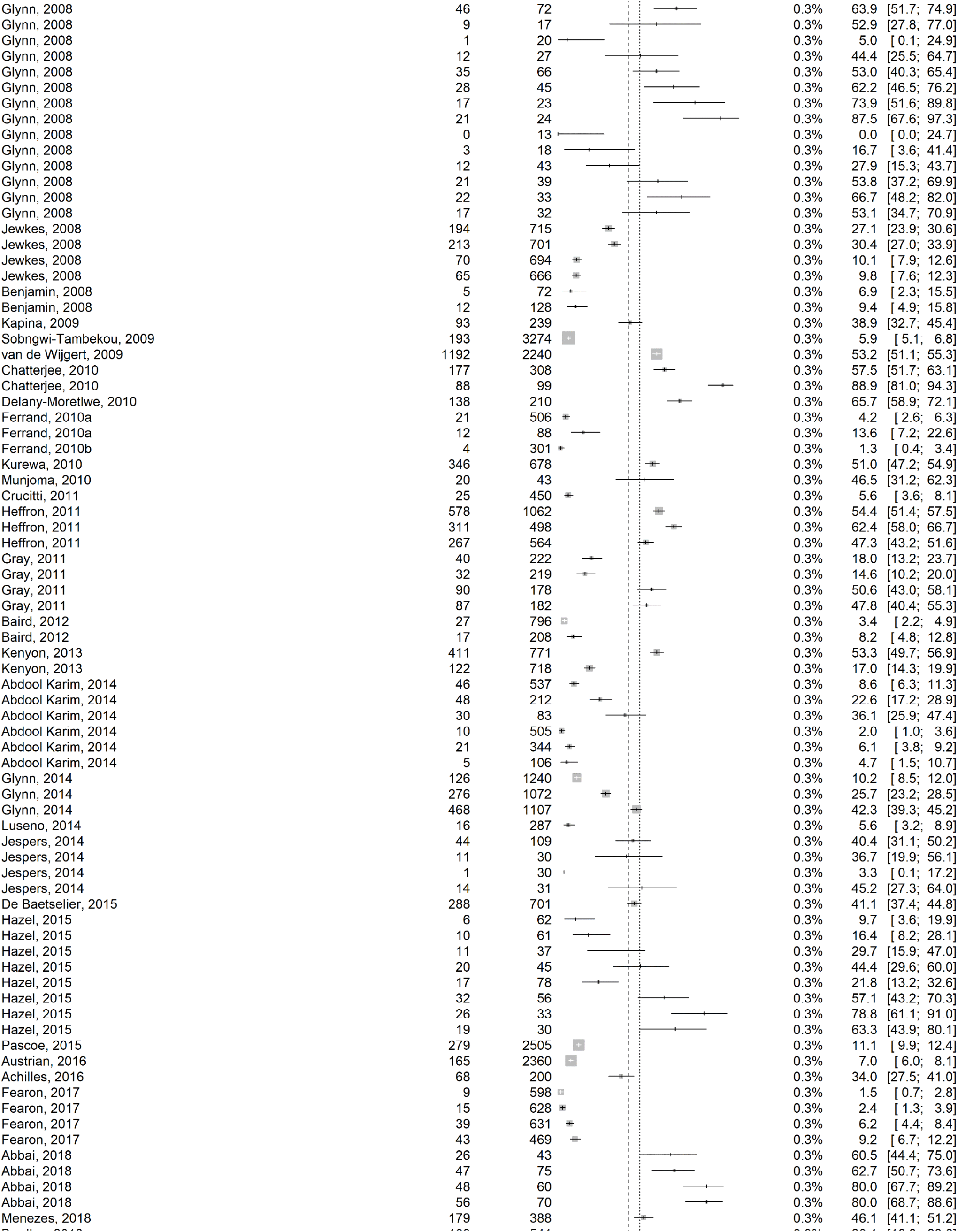
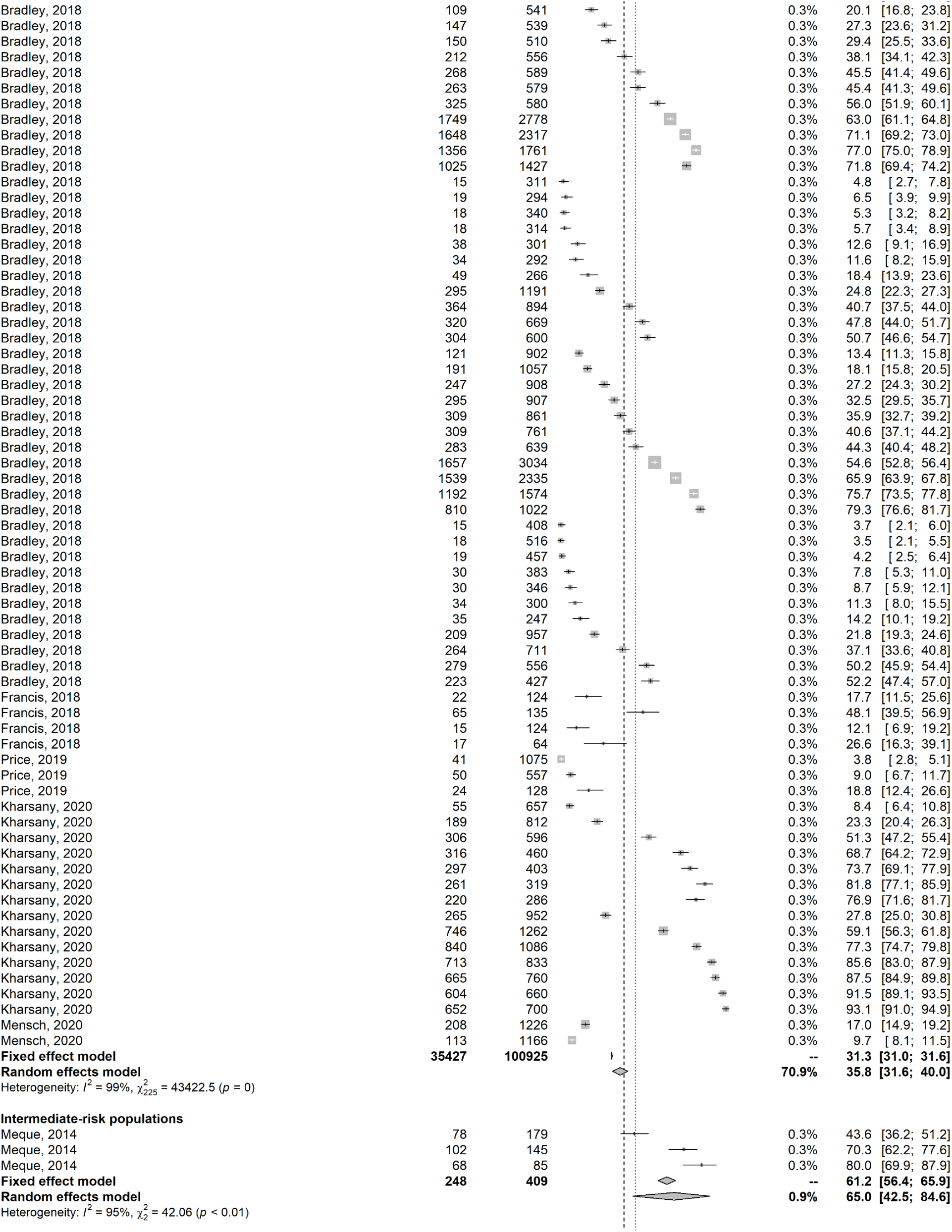
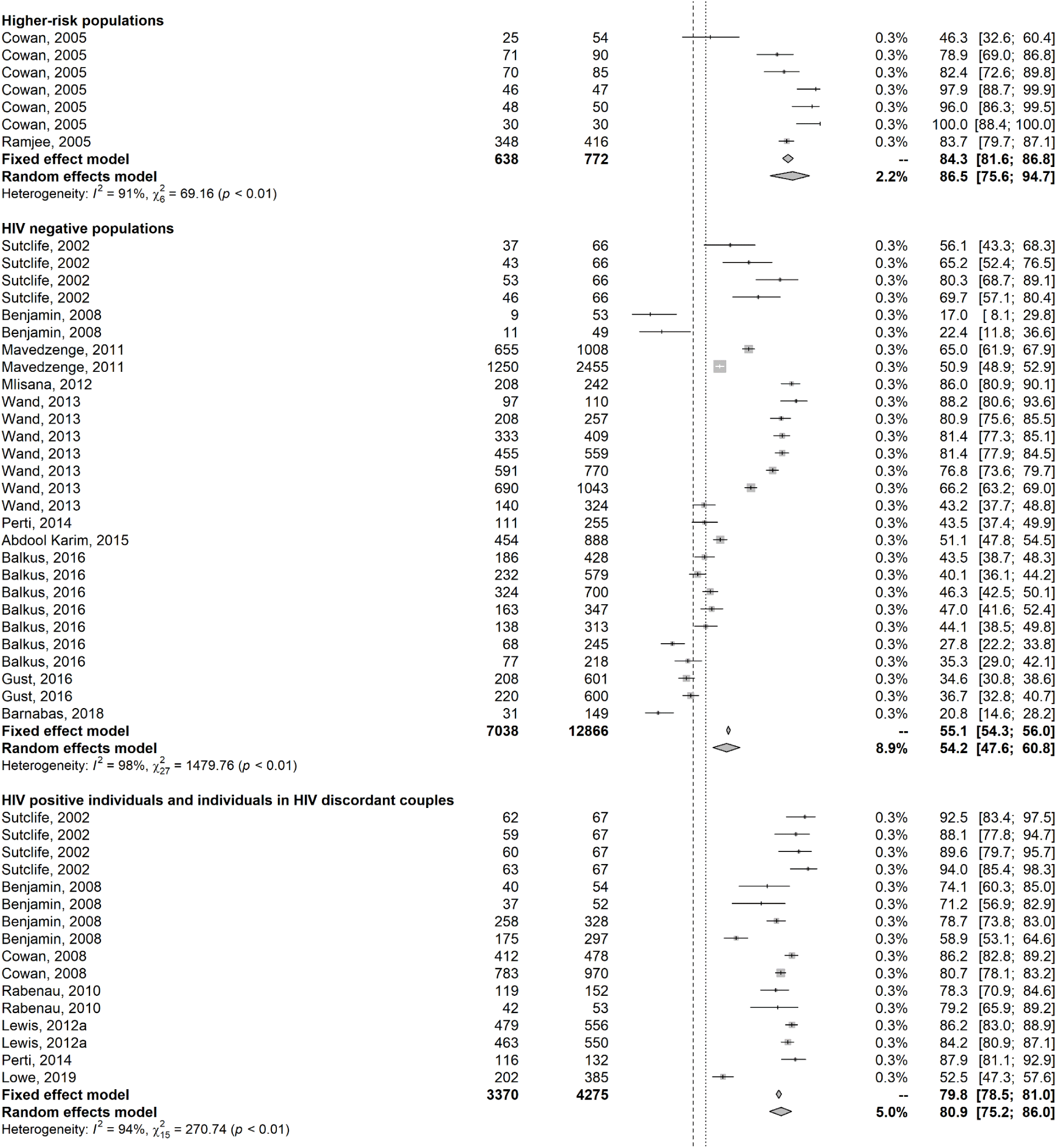
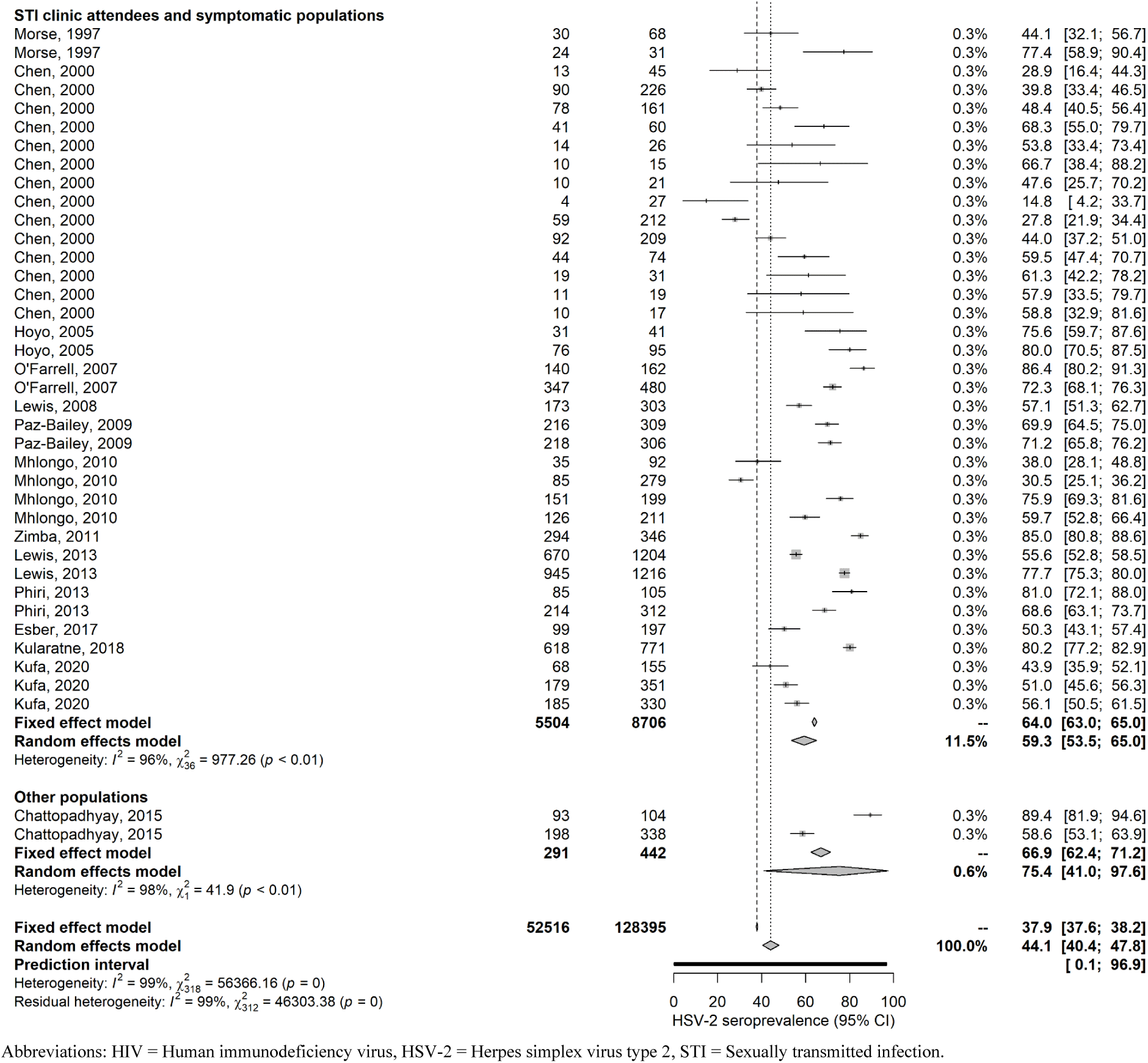
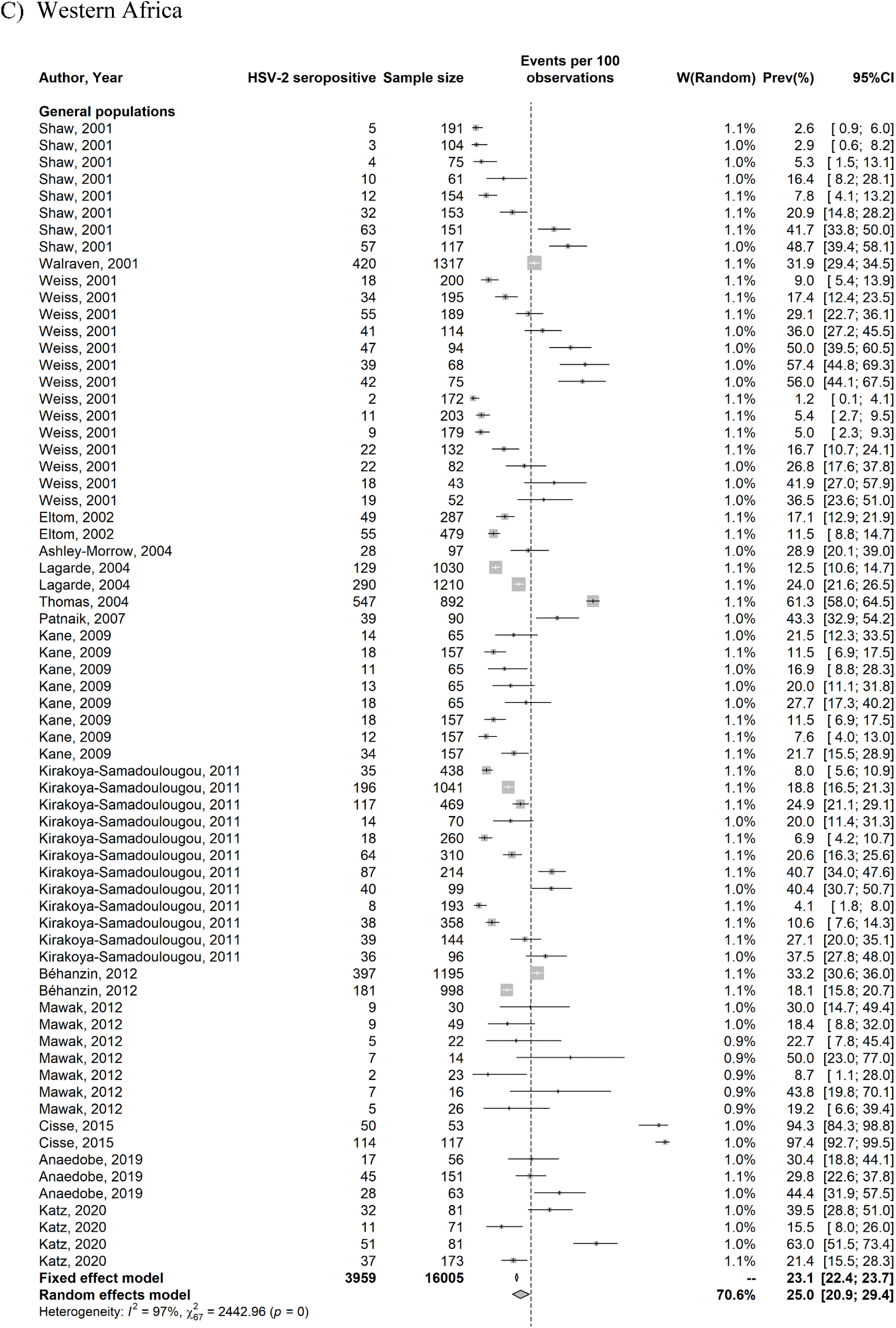
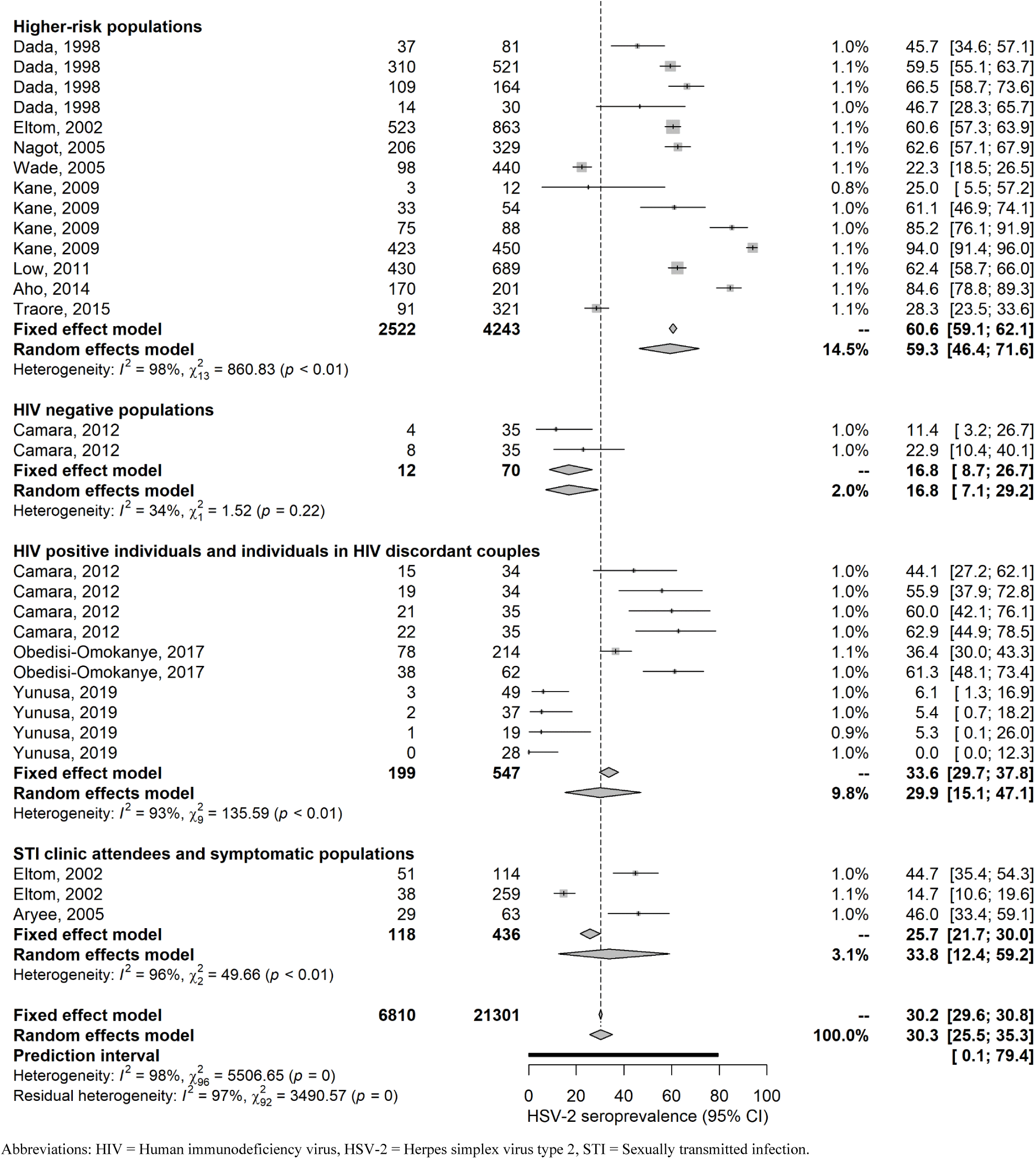
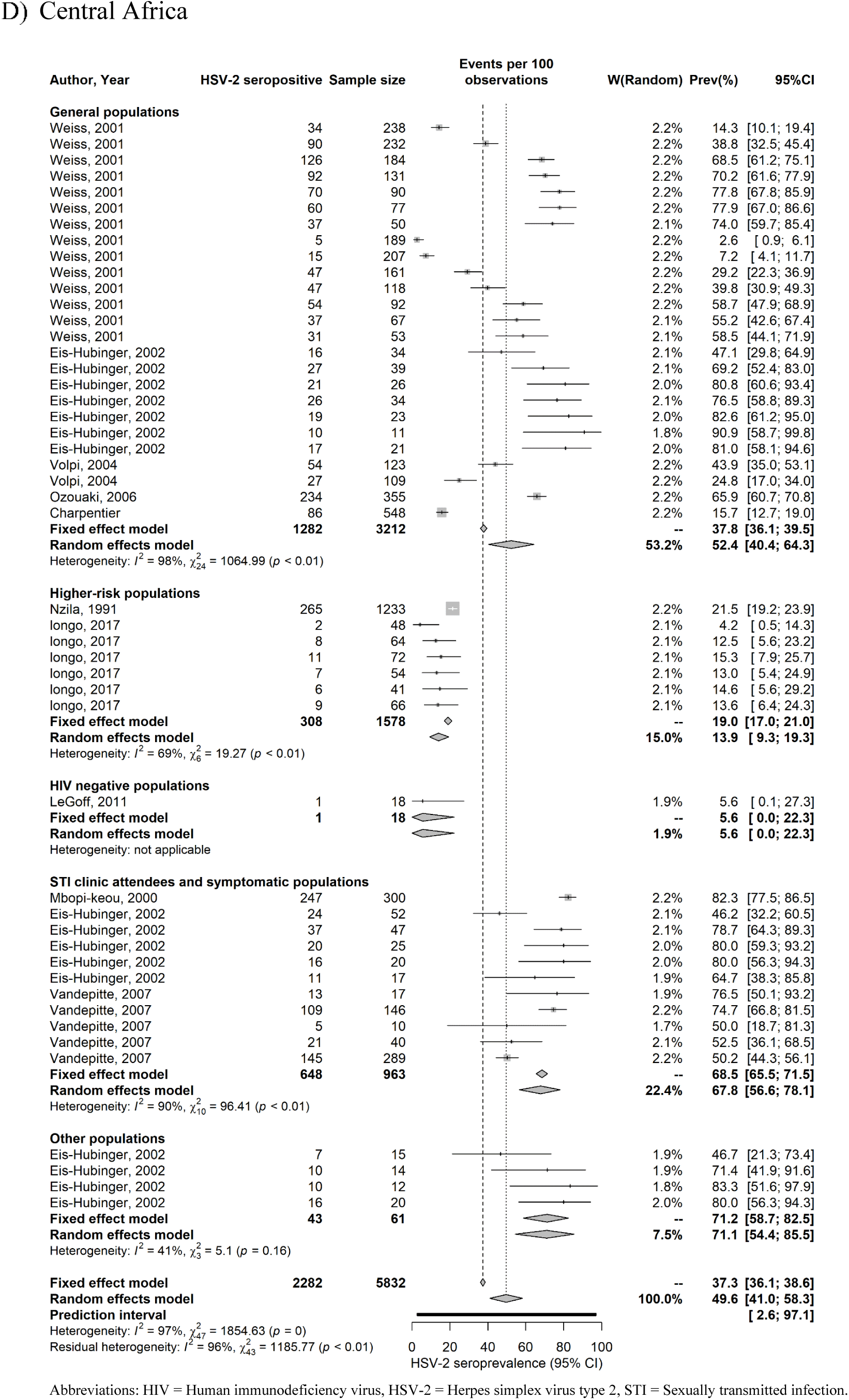
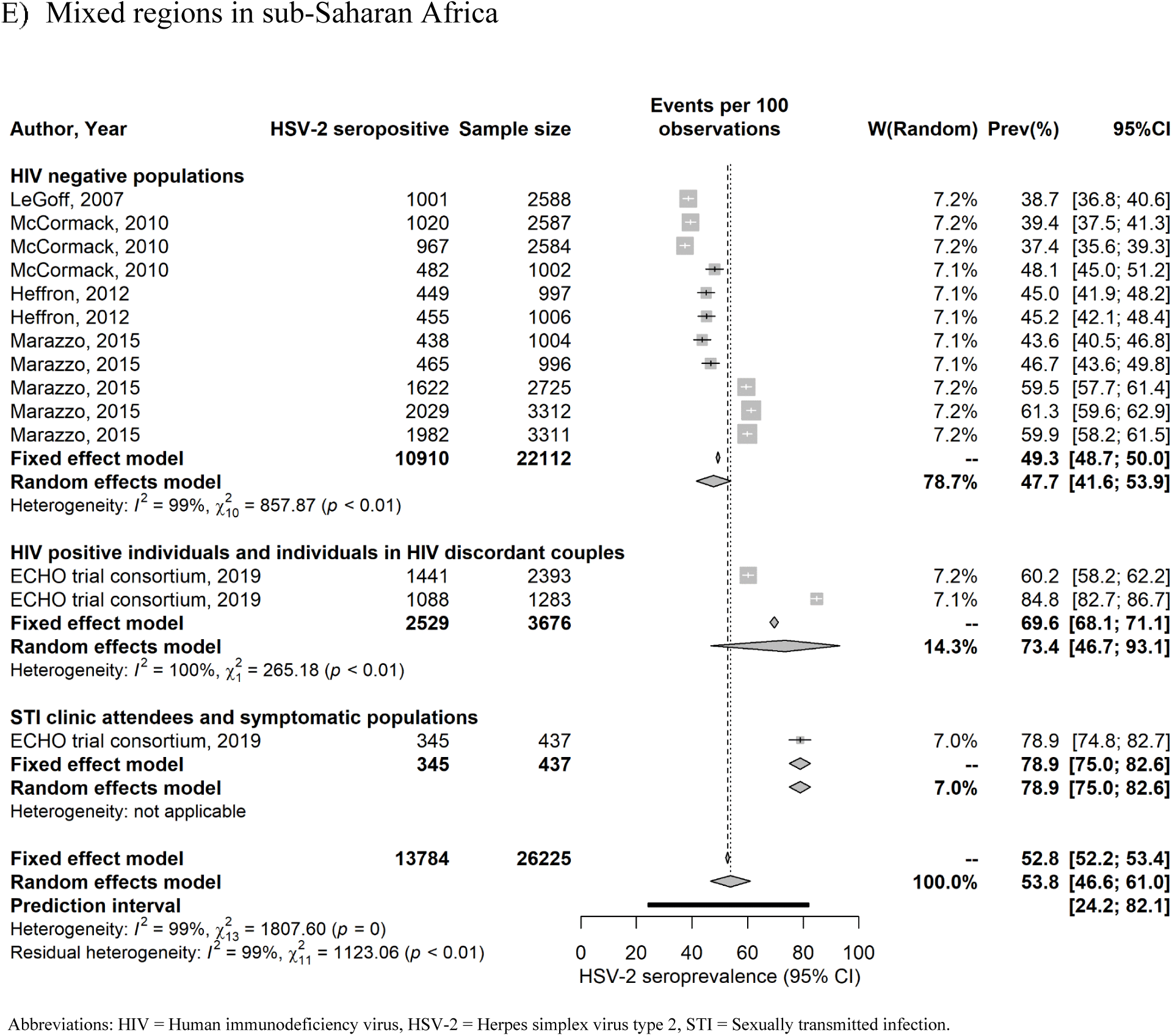
Forest plots presenting the outcomes of the pooled mean herpes simplex virus type 2 (HSV-2) seroprevalence among the different at risk populations across the sub-Saharan Africa subregions.

**Table S10.**
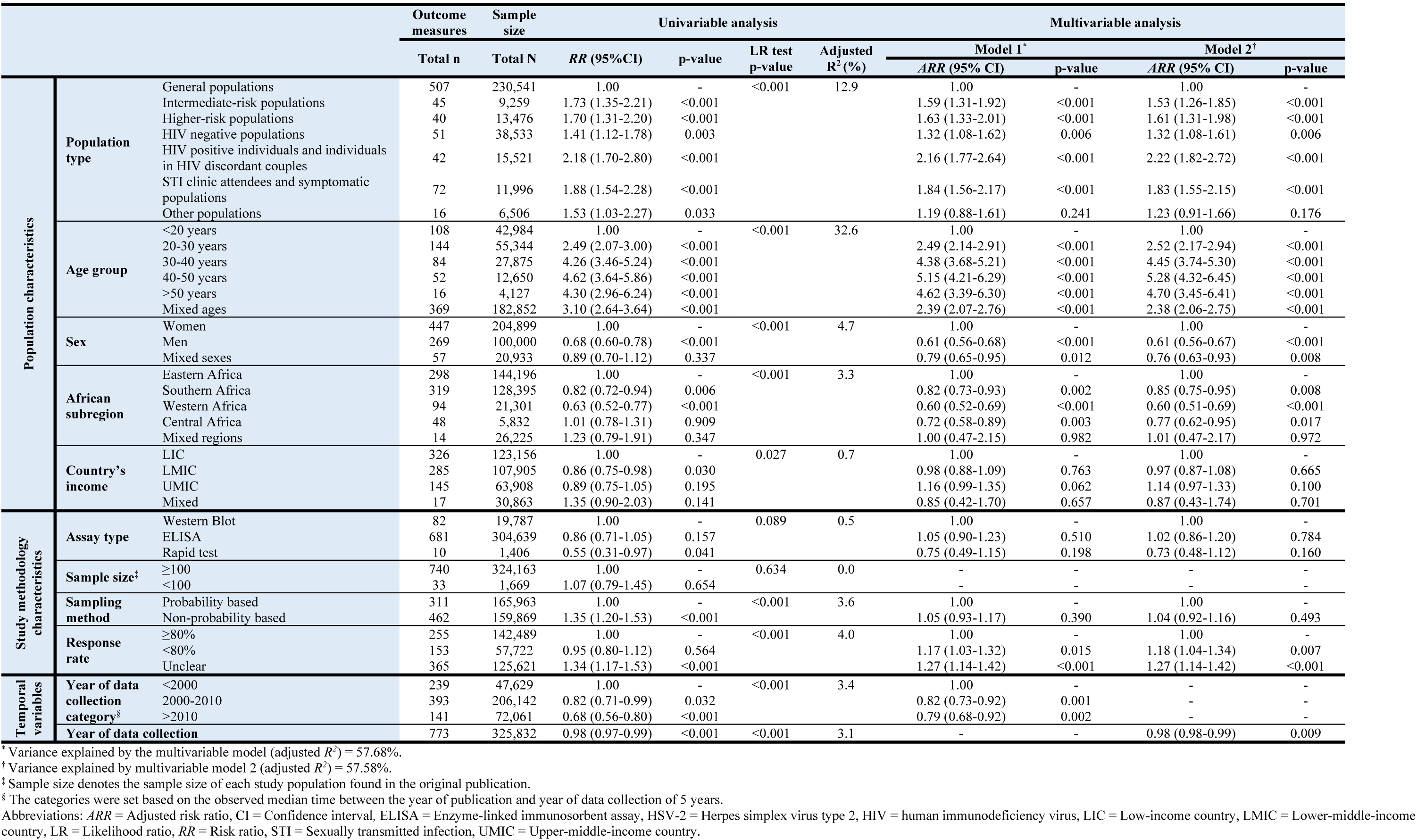
Univariable and multivariable meta-regression analyses for herpes simplex virus type 2 seroprevalence among the different at risk populations in sub-Saharan Africa using the year of data collection as a categorical variable or as a linear term (in replacement of year of publication).

**Table S11.**
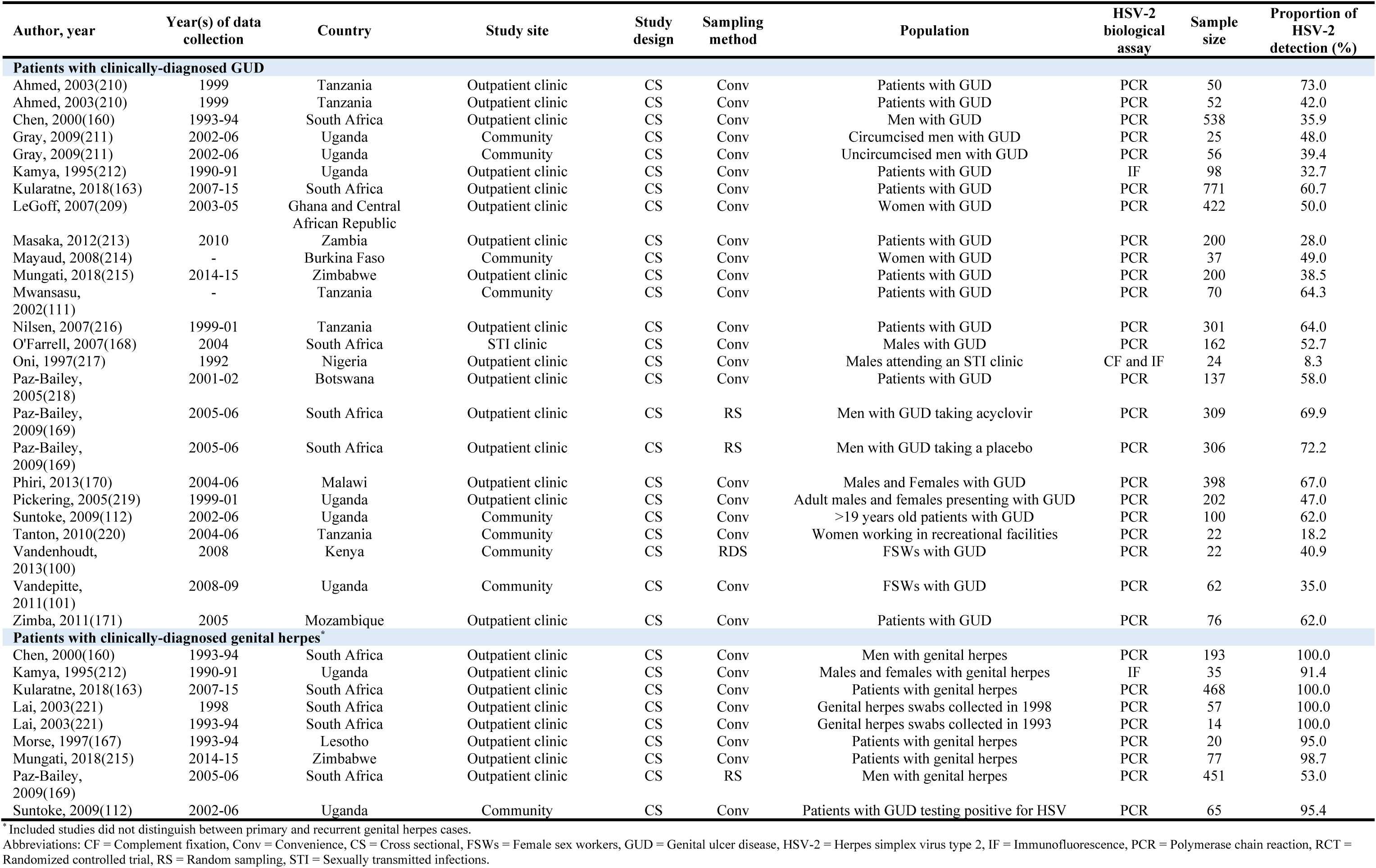
Studies reporting proportions of HSV-2 virus isolation in clinically-diagnosed genital ulcer disease and in clinically-diagnosed genital herpes in sub-Saharan Africa.

| Author, year | Year(s) of data collection | Country | Study site | Study design | Sampling method | Population | HSV-2 biological assay | Sample size | Proportion of HSV-2 detection (%) |
| --- | --- | --- | --- | --- | --- | --- | --- | --- | --- |
| **Patients with clinically-diagnosed GUD** |  |  |  |  |  |  |  |  |  |
| Ahmed, 2003(210) | 1999 | Tanzania | Outpatient clinic | CS | Conv | Patients with GUD | PCR | 50 | 73.0 |
| Ahmed, 2003(210) | 1999 | Tanzania | Outpatient clinic | CS | Conv | Patients with GUD | PCR | 52 | 42.0 |
| Chen, 2000(160) | 1993-94 | South Africa | Outpatient clinic | CS | Conv | Men with GUD | PCR | 538 | 35.9 |
| Gray, 2009(211) | 2002-06 | Uganda | Community | CS | Conv | Circumcised men with GUD | PCR | 25 | 48.0 |
| Gray, 2009(211) | 2002-06 | Uganda | Community | CS | Conv | Uncircumcised men with GUD | PCR | 56 | 39.4 |
| Kamya, 1995(212) | 1990-91 | Uganda | Outpatient clinic | CS | Conv | Patients with GUD | IF | 98 | 32.7 |
| Kularatne, 2018(163) | 2007-15 | South Africa | Outpatient clinic | CS | Conv | Patients with GUD | PCR | 771 | 60.7 |
| LeGoff, 2007(209) | 2003-05 | Ghana and Central African Republic | Outpatient clinic | CS | Conv | Women with GUD | PCR | 422 | 50.0 |
| Masaka, 2012(213) | 2010 | Zambia | Outpatient clinic | CS | Conv | Patients with GUD | PCR | 200 | 28.0 |
| Mayaud, 2008(214) | - | Burkina Faso | Community | CS | Conv | Women with GUD | PCR | 37 | 49.0 |
| Mungati, 2018(215) | 2014-15 | Zimbabwe | Outpatient clinic | CS | Conv | Patients with GUD | PCR | 200 | 38.5 |
| Mwansasu, 2002(111) | - | Tanzania | Community | CS | Conv | Patients with GUD | PCR | 70 | 64.3 |
| Nilsen, 2007(216) | 1999-01 | Tanzania | Outpatient clinic | CS | Conv | Patients with GUD | PCR | 301 | 64.0 |
| O'Farrell, 2007(168) | 2004 | South Africa | STI clinic | CS | Conv | Males with GUD | PCR | 162 | 52.7 |
| Oni, 1997(217) | 1992 | Nigeria | Outpatient clinic | CS | Conv | Males attending an STI clinic | CF and IF | 24 | 8.3 |
| Paz-Bailey, 2005(218) | 2001-02 | Botswana | Outpatient clinic | CS | Conv | Patients with GUD | PCR | 137 | 58.0 |
| Paz-Bailey, 2009(169) | 2005-06 | South Africa | Outpatient clinic | CS | RS | Men with GUD taking acyclovir | PCR | 309 | 69.9 |
| Paz-Bailey, 2009(169) | 2005-06 | South Africa | Outpatient clinic | CS | RS | Men with GUD taking a placebo | PCR | 306 | 72.2 |
| Phiri, 2013(170) | 2004-06 | Malawi | Outpatient clinic | CS | Conv | Males and Females with GUD | PCR | 398 | 67.0 |
| Pickering, 2005(219) | 1999-01 | Uganda | Outpatient clinic | CS | Conv | Adult males and females presenting with GUD | PCR | 202 | 47.0 |
| Suntoke, 2009(112) | 2002-06 | Uganda | Community | CS | Conv | >19 years old patients with GUD | PCR | 100 | 62.0 |
| Tanton, 2010(220) | 2004-06 | Tanzania | Community | CS | Conv | Women working in recreational facilities | PCR | 22 | 18.2 |
| Vandenhoudt, 2013(100) | 2008 | Kenya | Community | CS | RDS | FSWs with GUD | PCR | 22 | 40.9 |
| Vandepitte, 2011(101) | 2008-09 | Uganda | Community | CS | Conv | FSWs with GUD | PCR | 62 | 35.0 |
| Zimba, 2011(171) | 2005 | Mozambique | Outpatient clinic | CS | Conv | Patients with GUD | PCR | 76 | 62.0 |
| **Patients with clinically-diagnosed genital herpes\*** |  |  |  |  |  |  |  |  |  |
| Chen, 2000(160) | 1993-94 | South Africa | Outpatient clinic | CS | Conv | Men with genital herpes | PCR | 193 | 100.0 |
| Kamya, 1995(212) | 1990-91 | Uganda | Outpatient clinic | CS | Conv | Males and females with genital herpes | IF | 35 | 91.4 |
| Kularatne, 2018(163) | 2007-15 | South Africa | Outpatient clinic | CS | Conv | Patients with genital herpes | PCR | 468 | 100.0 |
| Lai, 2003(221) | 1998 | South Africa | Outpatient clinic | CS | Conv | Genital herpes swabs collected in 1998 | PCR | 57 | 100.0 |
| Lai, 2003(221) | 1993-94 | South Africa | Outpatient clinic | CS | Conv | Genital herpes swabs collected in 1993 | PCR | 14 | 100.0 |
| Morse, 1997(167) | 1993-94 | Lesotho | Outpatient clinic | CS | Conv | Patients with genital herpes | PCR | 20 | 95.0 |
| Mungati, 2018(215) | 2014-15 | Zimbabwe | Outpatient clinic | CS | Conv | Patients with genital herpes | PCR | 77 | 98.7 |
| Paz-Bailey, 2009(169) | 2005-06 | South Africa | Outpatient clinic | CS | RS | Men with genital herpes | PCR | 451 | 53.0 |
| Suntoke, 2009(112) | 2002-06 | Uganda | Community | CS | Conv | Patients with GUD testing positive for HSV | PCR | 65 | 95.4 |

\* Included studies did not distinguish between primary and recurrent genital herpes cases.
Abbreviations: CF = Complement fixation, Conv = Convenience, CS = Cross sectional, FSWs = Female sex workers, GUD = Genital ulcer disease, HSV-2 = Herpes simplex virus type 2, IF = Immunofluorescence, PCR = Polymerase chain reaction, RCT = Randomized controlled trial, RS = Random sampling, STI = Sexually transmitted infections.

**Figure S3.**
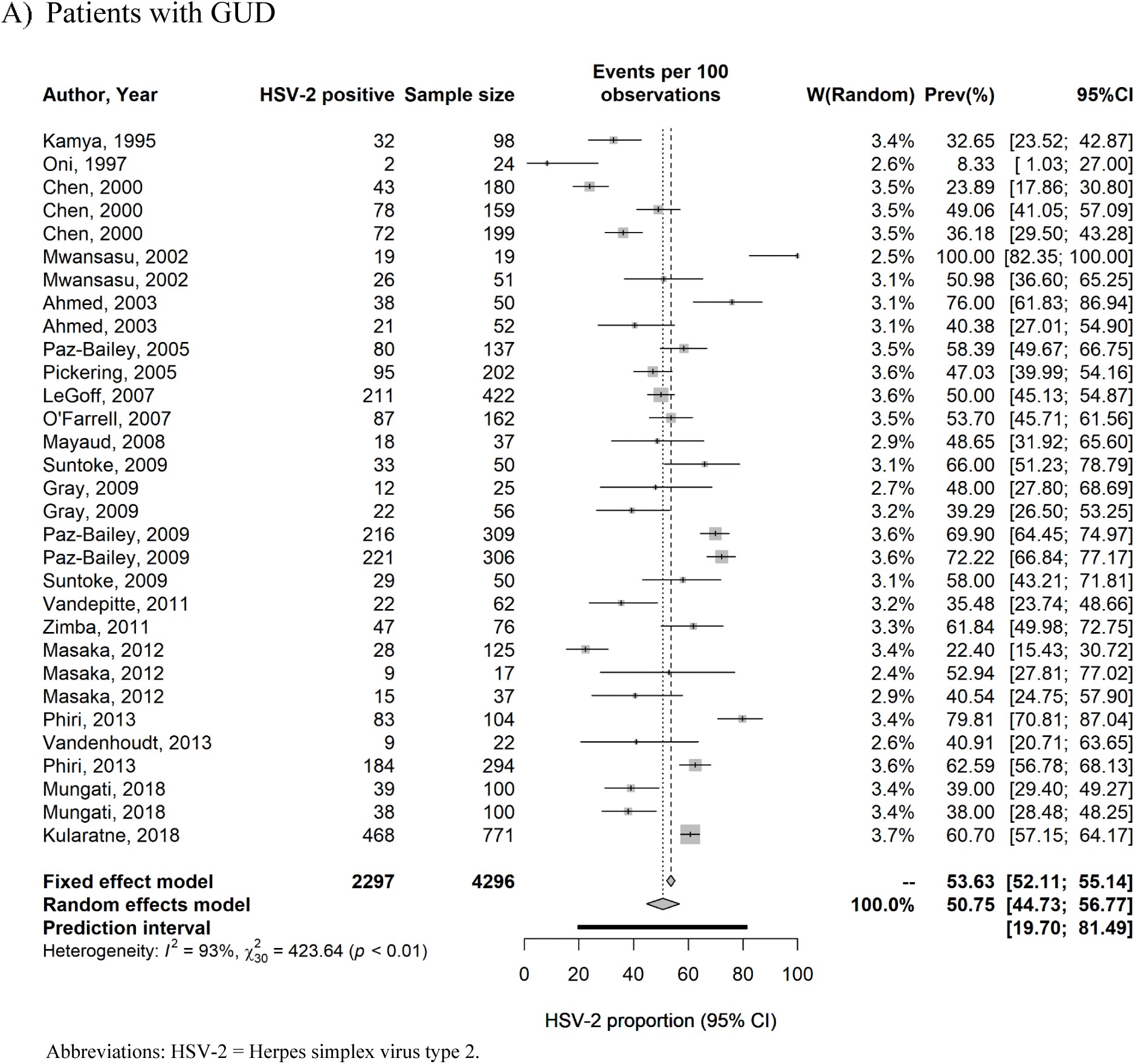
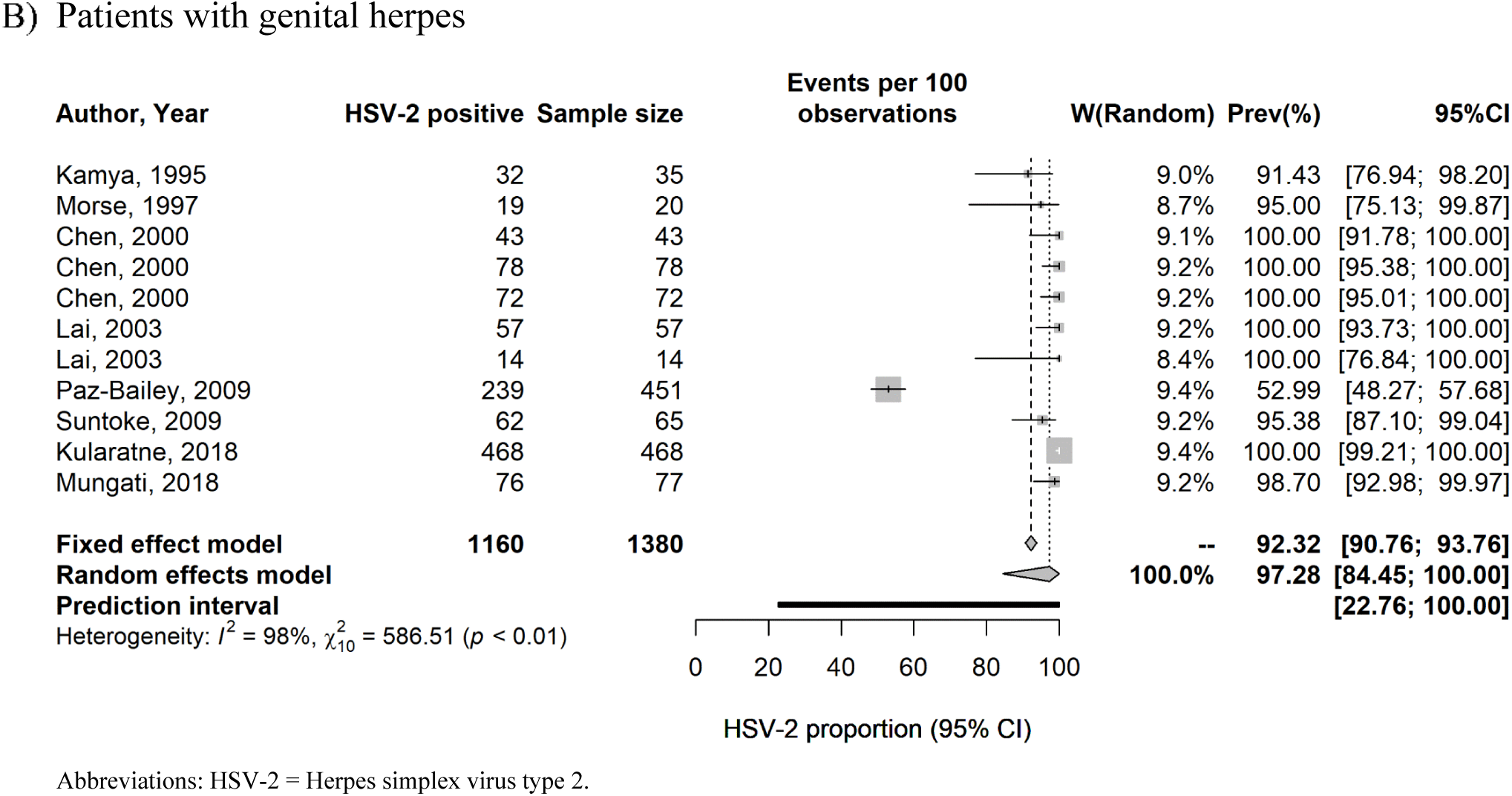
Forest plots presenting the outcomes of the pooled mean proportions of HSV-2 virus isolation in clinically-diagnosed genital ulcer disease (GUD) and in clinically-diagnosed genital herpes in sub-Saharan Africa.

**Table S12.** Summary of the precision assessment and risk of bias assessment for the studies reporting HSV-2 seroprevalence in sub-Saharan Africa.

| Quality assessment | HSV-2 seroprevalence measures |  |
| --- | --- | --- |
|  | Number of studies | % |
| **Precision of seroprevalence measures<sup>a</sup>** |  |  |
| Low precision | 34 | 10.6 |
| High precision | 288 | 89.4 |
| **Risk of bias quality domain<sup>b</sup>** |  |  |
| **Sampling method** |  |  |
| Low risk of bias | 88 | 27.3 |
| High risk of bias | 234 | 72.7 |
| **Response rate** |  |  |
| Low risk of bias | 85 | 26.4 |
| High risk of bias | 45 | 14.0 |
| Unclear risk of bias | 192 | 59.6 |
| **Summary of the risk of bias assessment** |  |  |
| **Low risk of bias** |  |  |
| In at least one quality domain | 107 | 33.2 |
| In both quality domains | 39 | 12.1 |
| **High risk of bias** |  |  |
| In at least one quality domain | 91 | 28.3 |
| In both quality domains | 23 | 7.1 |
| **Seroprevalence studies where risk of bias assessment was possible** | **322** | **100** |

<sup>a</sup>Precision was assessed based on the overall sample size (not each stratum subsample size) of the study as reported in the record/publication.
<sup>b</sup>Risk of bias was assessed based on the overall sample size (not each stratum subsample size) of the study as reported in the record/publication.
Abbreviations: HSV-2 = Herpes simplex virus type 2.

